# An approach to finding specific forms of dysbiosis that associate with different disorders

**DOI:** 10.1101/2024.04.23.24306162

**Authors:** Jonathan Williams, Inga Williams, Karl Morten, Julian Kenyon

## Abstract

**Background:** Many disorders display dysbiosis of the enteric microbiome, compared with healthy controls. Different disorders share a pattern of dysbiosis that may reflect ‘reverse causation’, due to non-specific effects of illness-in-general. Combining a range of disorders into an ‘aggregate non-healthy active control’ (ANHAC) group should highlight such non-specific dysbiosis. Differential dysbiosis between the ANHAC group and specific disorders may then reflect effects of treatment or bowel dysfunction, or may potentially be causal. Here, we illustrate this logic by testing if individual genera can differentiate an ANHAC group from two specific diagnostic groups.

**Methods:** We constructed an ANAHC group (n=17) that had 14 different disorders. We then used random forest analyses to test differential dysbiosis between the ANHAC group and two other disorders that have no known pathology, but: (i) symptoms of illness (Myalgic Encephalomyelitis / Chronic Fatigue Syndrome – ME/CFS – n = 38); or (ii) both illness and bowel dysfunction (ME/CFS comorbid with Irritable Bowel Syndrome – IBS – n=27).

**Results:** Many genera differentiated the ANHAC group from co-morbid IBS. However, only two genera - Roseburia and Dialister – discriminated the ANHAC group from ME/CFS.

**Conclusions:** Different disorders can associate with specific forms of dysbiosis, over-and-above non-specific effects of illness-in-general. Bowel dysfunction may contribute to dysbiosis in IBS via reverse causation. However, ME/CFS has symptoms of illness-in-general, but lacks known pathology or definitive treatment that could cause dysbiosis. Therefore, the specific dysbiosis in ME/CFS may be causal. [230 words]

**Contribution to the field:** Many disorders associate with enteric dysbiosis. The pattern of dysbiosis is largely consistent between unrelated disorders, which suggests that it mainly reflects non-specific secondary effects of illness-in-general (e.g. due to changes in activity levels, or diet). However, faecal microbiome transplantation (FMT) can be therapeutic in some disorders. This implies that unique features of dysbiosis may cause those specific disorders. Here, we propose a way to assess causal effects of dysbiosis, by testing if individual genera can discriminate individual disorders from an ‘aggregate non-healthy active control’ (ANHAC) group. Dysbiosis in the ANHAC group can control for non-specific effects of illness-in-general on the microbiome and so highlight potentially-causal forms of dysbiosis in specific disorders. This approach may provide insight into pathogenetic mechanisms of individual disorders and help to design specific forms of FMT to counteract them.

## Introduction

Many disorders associate with enteric dysbiosis. The pattern of dysbiosis is largely consistent between unrelated disorders.[1] So, this shared pattern may reflect reverse causation, due to non-specific secondary effects of illness-in-general (e.g. changes in activity, or diet). However, faecal microbiome transplantation (FMT) can be therapeutic in some disorders,[2–6] which indicates that unknown elements of dysbiosis may cause those specific disorders. Identifying such causal elements of dysbiosis could illuminate pathogenetic mechanisms and hence improve therapeutic strategies. The first step toward this identification is to control for non-specific secondary effects of illness-in-general on the microbiome.[1] Here, we propose a method to achieve this.

In principle, (a) illness-in-general may cause dysbiosis, (b) specific features of single disorders may cause dysbiosis, or (c) dysbiosis can cause single disorders. In order to highlight (b) and (c), we propose eliminating (a) by constructing an ‘aggregate non-healthy active control’ (ANHAC) group that includes a range of unrelated disorders (c.f.[7]). The profile of dysbiosis in this ANHAC group should represent the ‘shared signature’[1] of illness-in-general ((a) above – see detailed rationale in Methods). Hence, differential dysbiosis between the ANHAC group and single disorders may reflect specific associations of dysbiosis with those single disorders ((b) and (c), above).

Ideally, an ANHAC group should comprise patients with a wide range of disorders whose demographic and clinical (e.g. organ system and treatment) characteristics resemble those of the target disorder(s). Each disorder in the ANHAC group may possess a specific form of dysbiosis, but overall these specific associations should cancel. Hence, if the dysbiosis in the whole ANHAC group differs from that in a single target disorder, then the elements of dysbiosis that differentiate the two groups may be unique to the target disorder. Moreover, if the demographic and clinical features and treatments of the ANHAC group and target disorder match, then elements of dysbiosis that are unique to the target disorder may be causal.

One way to assess our proposed method is to compare the ANHAC group with single disorders that have no established pathology or definitive treatment. Irritable Bowel Syndrome (IBS) lacks known pathology or definitive treatment, but associates with bowel dysfunction. Hence differential dysbiosis between the ANHAC group and IBS may be secondary to bowel dysfunction. Going one step further, Myalgic Encephalomyelitis/Chronic Fatigue Syndrome (ME/CFS) also lacks known pathology or definitive treatment or major bowel dysfunction (comorbidity between IBS and ME/CFS is common[8, 9], but not ubiquitous). Therefore, because ‘average’ dysbiosis in an ANHAC group should reflect only illness-in-general, any differential dysbiosis between an ANHAC group and ME/CFS should not result from ‘reverse causation’ (see above), but may reflect microbiome features that contribute to causing ME/CFS.

In principle, a potential problem for our proposed method is heteroscedasticity. That is, the ANHAC group is diagnostically heterogeneous by design, so that the *mean* abundance of each genus reflects the effects of illness-in-general. However, the diagnostic heterogeneity may cause the *range* of observations of each individual genus to be wide. Such heteroscedasticity may make it impossible to detect differences between the *mean* levels of individual genera in the ANHAC group and individual disorders. One goal of the present study is to determine how far the problem of heteroscedasticity may be important in practice.

## Methods

### Ethics

We obtained microbiome data routinely for the primary purpose of guiding clinical management. Additionally, we obtained written consent from each participant to use their data for research. In line with UK legislation, the South Central Hampshire Research Ethics Committee deemed that our study did not, therefore, need ethical review.

### Patients

We studied 4 groups of patients – ANHAC, cancer, ME/CFS and IBS. We report only comparisons between the ANHAC group and ME/CFS or IBS, here (the comparison between the ANHAC and cancer groups is in the Supplementary information). Our IBS group comprised mainly people with co-morbid IBS and ME/CFS (such co-morbidity is common[3, 10–12] – but see[13]). This co-morbidity is advantageous in the present context, as it can highlight effects of bowel dysfunction.

We have reported data from some participants previously and posted the current data-set online in May 2023.[3, 14, 15] Here, we analysed only participants aged over 14. None had received FMT, but (i) all received dietary advice, (ii) those with established pathologies or treatments had ‘treatment-as-usual’ and (iii) those with ME/CFS or IBS had received β-glucan 1.3 and 1.6 for about 4 weeks prior to stool sampling. Here we coarsened patients’ ages to the nearest 5 years.

### Microbiome DNA sequencing

Participants collected stool samples and sent them for analysis via bacterial 16S ribosomal RNA gene sequencing of bacterial DNA (Atlas Biomed - see[16]). Specifically, identification of microbiota used NanoSeq Illumina paired sequencing to get 250bps after merging. The analysis generated Amplicon Sequence variants using QIIME2.[17]

### Rationale

Our design extends that of Gupta et al., who pooled stool metagenomes from 12 different disease or abnormal bodyweight conditions into “a single aggregate nonhealthy group”.[7] Gupta and colleagues reasoned that the comparing the profile of abundances of different genera in this aggregate nonhealthy group with the corresponding profile of healthy people would help to define the dysbiosis associated with illness-in-general. We extended Gupta’s logic to compare our aggregate non-healthy active control (ANHAC) group with groups that had single disorders – ME/CFS or IBS. We reasoned these comparisons could identify genera that associate specifically (and, possibly, causally) with individual disorders, after accounting for effects of illness-in-general.

Following the above reasoning, we tested if abundances of individual genera in an ANHAC group differ from those in single disorders. Specifically, we tested differential dysbiosis between an ANHAC group and two specific disorders: (1) IBS; and (2) ME/CFS. IBS lacks known pathology, so (1) can assess how far bowel dysfunction may cause dysbiosis; differential dysbiosis between ME/CFS and the ANHAC group may reflect causes of ME/CFS (see Introduction).

The present method requires the ANHAC group to be heterogeneous. However, if the microbiota of the ANHAC group are too heterogeneous, then comparisons with other, single-diagnosis groups may lack power to detect differences (see Introduction). We initially used binomial tests to compare (i) the proportion of zeros and (ii) the variances of each genus in the ANHAC group with those in each single-diagnosis group. However, (i) and (ii) inter-relate. Therefore we assessed heterogeneity more accurately using hierarchical mixed-membership beta regression, to test effects of diagnostic grouping on variance of abundances and on zero inflation while accounting for different levels of abundance of each genus between people (see Supplementary Information).

We used random forest (RF) analyses to test if the relative abundances of microbial genera can predict diagnostic categories.[18] RF may be optimal for this purpose.[19–22] However, RF tends to perform badly in high-dimensional analyses with a low fraction of relevant predictors and small sample sizes.[23] Therefore, we used preliminary RF analyses to select important variables for each classification, before constructing large RFs that used the selected variables to predict each diagnostic grouping. We weighted the RFs’ sampling procedures to eliminate effects of imbalance between group sizes, by down-sampling the larger group when growing decision trees.

We assessed the RFs’ abilities to predict each diagnostic grouping using the areas under the Receiver Operating Characteristic curves (AUROCs) along with their overall classification errors, which are available from the RF output.[24] We computed the 95% confidence limits for each AUROC.[25] The Supplementary Information gives the details of the RF analyses.

Finally, we assessed the dependence of clinical groupings on abundances of different microbiota qualitatively, by generating partial plots to show the forms of their (independent) effects on diagnostic groupings. RF analyses can, in principle, discriminate different groups on the basis of heteroscedasticity (the variances of individual genera in each group), rather than differences in location (the mean abundance of each genus in each group). In this case, the partial relation between group membership and the abundance of a genus should show a biphasic (U-shaped) form. In contrast, if the RF discriminates two groups on the basis of the mean abundance of a genus in each group, then the partial relationship should show a monotonic form. We examined the partial dependence of group membership on each genus that was important in the RF analyses.

## Results

### Sample

The main study included 82 participants (see Supplementary Information for cancer group). Three-quarters (61/82) were female and the median age was 58 (IQR 35-67). The age and sex distributions of these diagnostic groups were similar (age: KW χ^2^ = 0.8, 2df, p=0.67; sex χ^2^ = 2.7, 2df, p=0.25).

The ANHAC group comprised 17 people with miscellaneous disorders (1 each of morbid obesity, acne rosacea, polymyalgia rheumatica, alopecia areata, anxiety and insomnia, autism, herpes genitalis, seborrhoeic dermatitis, eczema, Alzheimer-type dementia, and Parkinson’s Disease; 2 each with Crohn’s disease, ulcerative colitis, and motor neurone disease); 38 people had ME/CFS; 27 people had IBS (23 co-morbid with ME/CFS).

### Compositions of microbiota

Data were available for 193 genera, but only 94 genera had fewer than 80% non-zero abundances (see Supplementary Table S1). There was no evidence of heteroscedasticity between different diagnostic groups (see Supplementary Information).

### Differential dysbiosis between the ANHAC group and single-diagnosis groups

The proportions of individual genera discriminated the ANHAC group from the ME/CFS and IBS groups (see Table 1).

**Table 1:** accuracy of the discrimination between clinical groupings by the random forest analyses

| Measure | Comparison: | ANHAC vs IBS | ANHAC vs ME/CFS |
| --- | --- | --- | --- |
| AUROC |  | 71.2 (55.4 – 87.0) | 70.9 (56.2 – 85.6) |
| Error rate |  | 31.8 | 34.5 |

Values are rounded percentages, with 95% confidence limits in parentheses. Key: AUROC = Area Under the Receiver Operating Characteristic curve. The error rate is the overall proportion of misclassification of cases in each analysis. Larger AUROCs and smaller error rates indicate better discrimination. Note that analyses that used the true ages, prior to coarsening them to meet the requirements of publication, yielded slightly higher AUROCs (71.8 for both IBS and ME/CFS) and lower error rate for ME/CFS (27.3).

Thirty individual genera discriminated the ANHAC group from the IBS group (see Supplementary Information). In contrast, only two specific genera – Roseburia and Dialister – discriminated the ANHAC group from the ME/CFS group (see Figures 1A-1B). The grouping of ‘Unclassified Bacteria’ also contributed to discriminating the ANHAC group from both the IBS and CFS groups. The Supplementary Information shows the full details of the dependence of diagnostic categories on individual genera that contribute to differential dsybiosis.

**Figures 1A-1B.**
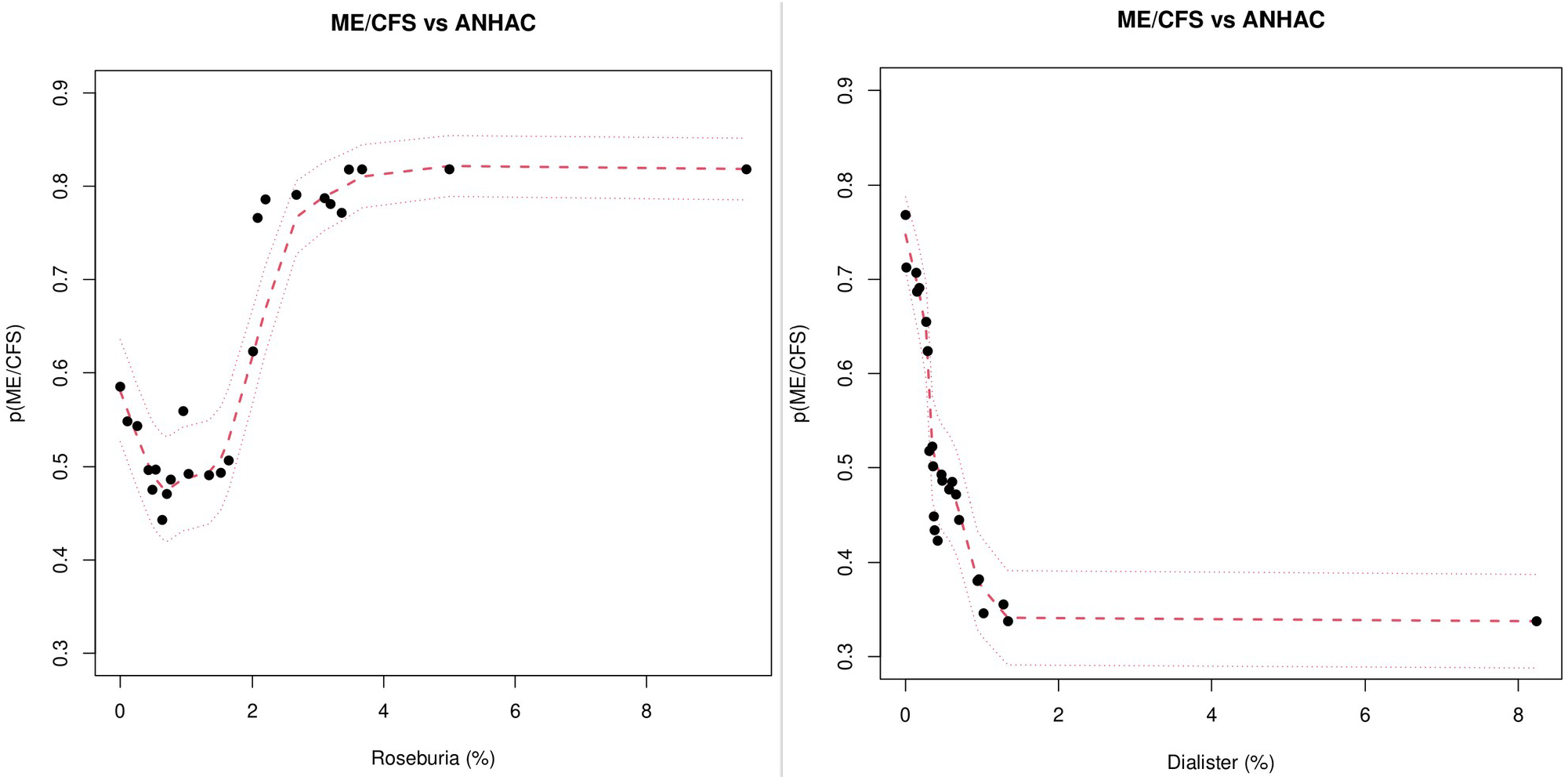
partial plots of the dependence of ME/CFS on Roseburia and Dialister.

Each panel shows the probability of ME/CFS (compared with theANHAC group; y-axis) over the range of abundances (%) of an individual genus (x-axis), adjusted for all other predictors in the random forest (RF) model. Both plots show monotonic dependence of ME/CFS on the abundance of the genera, which indicates that heteroscedasticity did not exert undue influence in the RF model. See Supplementary files for partial plots of all genera selected for the random forest analyses.

## Discussion

The present study found that proportions of microbiota can discriminate a heterogeneous aggregate non-healthy active-control (ANHAC) group from other, more homogeneous patient groups. Overall dysbiosis in the ANHAC group may reflect non-specific secondary effects of illness-in-general. Hence, differential dysbiosis between the ANHAC group and other, more homogeneous diagnostic groups may reflect specific effects of pathologies or their treatments in the homogenous groups. Alternatively, in groups that lack established pathology or definitive treatment (e.g. ME/CFS) – differential dysbiosis from the ANHAC group may indicate causal elements of the microbiome. Overall, our results provide preliminary proof of the principle that this approach can delineate specific dysbiosis in individual disorders, which may help to tailor appropriate treatments.

The present study did not include healthy controls. Many single-disorder groups differ from healthy controls[1, 7] and there is “a common signal for gut dysbiosis …. shared across unrelated diseases”. [1] Hence, it is likely that (a) the microbiomes of healthy controls will differ from any patient group (e.g. [1, 26–28]); and (b) our ANHAC group showed dysbiosis, overall. Consistent with (b), the random forest could not differentiate the ANHAC group from a grouping of cancer diagnoses (see Supplementary Information and compare[1, 29–32]). However, the absence of healthy controls limits characterization of our ANHAC group and the nature of its dysbiosis. This limitation (which we address further in the Supplementary Information) does not prevent the proof of principle of our approach of comparing an ANHAC group with single-disorder groups. Further studies should compare ANHAC groups with healthy controls to assess the nature of dysbiosis that reflects illness-in-general.[7]

The random forest analyses differentiated the ANHAC grouping from both the ME/CFS and IBS groups. Previous studies have shown that the microbiomes of healthy controls differ from both IBS[1, 33, 34] and from ME/CFS.[14, 35–38] So, the differences that we observed imply that ME/CFS and IBS have particular forms of dysbiosis. Almost all of the IBS group had co-morbid IBS and ME/CFS. Hence, the genera that discriminate the ANHAC and IBS groups may represent the combined effects of ME/CFS and gastrointestinal dysfunction. Consistent with this, low levels of Dialister and high levels of Roseburia or Unclassified bacteria contributed to discriminating both ME/CFS and IBS from the ANHAC grouping (see Supplementary file of partial plots). These convergent findings strengthen each other. The remaining genera that discriminate IBS from the ANHAC grouping may reflect causes or effects of the gastrointestinal dysfunction in IBS.

The ME/CFS group had higher proportions of Roseburia than the ANHAC group. The logic of comparing single disorders with an ANHAC group is that this can control for ‘reverse causation’ due to illness-in-general (see Introduction). Since ME/CFS lacks established pathology,[1, 39] our result implies that Roseburia may cause ME/CFS. At first glance, this conflicts with reports that *low* abundance of Roseburia generally associates with illness-in-general[1] and fatigue.[40] However, people with ME/CFS complain of cognitive difficulties (“brain fog”[41, 42]) and our observation of more Roseburia in ME/CFS fits with findings of recent Mendelian Randomisation (MR) studies that higher Roseburia can *cause* worse cognition.[43, 44] Additionally, our observation of low Dialister in ME/CFS fits with findings that levels of Dialister were almost significantly lower in ME/CFS[1] and that low levels of Dialister can *cause* worse cognition.[43, 44] It is unlikely that 2 out of 3 taxa that predicted ME/CFS in our small study would concur with findings of causal MR analyses of a large sample (¼ million) by chance alone (p=0.006 – see Supplementary Information). Hence, the congruity of these results reinforces our logic that excluding reverse causation, by comparing single disorders with an ANHAC group, may help to identify causal elements of dysbiosis.

The ME/CFS group had a higher proportion of “Unclassified Bacteria” than the ANHAC group. Despite advances in understanding the microbiome, many of its bacteria remain unclassified. Again, the logic that comparing single disorders with an ANHAC group can exclude reverse causation suggests that chronic infection with unknown bacteria could cause ME/CFS. An alternative possibility is that Roseburia may interact with an unknown taxon (see Supplementary Information). If confirmed, these possibilities would have important clinical implications.

No other studies have compared individual disorders with an ANHAC group. Therefore, we used publicly-available data (see Supplementary Information) to do this for two of the specific disorders that we studied – cancer and IBS. In brief, we used publicly-available data from a recent large study[1] to create a new ANHAC group that represents the microbiome signature of illness-in-general. We reasoned that deviations from this signature in specific disorders may reflect specific forms of dysbiosis that associate uniquely with those disorders. The genera that deviated from the signature of illness-in-general in the regressions for cancer and IBS in those independent data correspond with the genera that random forest selected to discriminate these disorders in our study. This correspondence reinforces the reliability and validity of the ANHAC approach to defining associations between specific disorders and specific forms of dysbiosis. We describe the methods, results and limitations of these analyses in more detail the Supplementary Information.

Heteroscedasticity was not a major limitation for the ANHAC comparisons. Not only did these comparisons yield results that resemble other recent findings (see above), but the partial plots of group membership on the abundances of individual genera were monotonic (which indicates that heteroscedasticity may be unimportant – see Methods). However, the ANHAC group in our study was small, which may limit the influence of heteroscedasticity. Further studies should construct larger ANHAC groups and examine the influence of heteroscedasticity more fully.

We used random forest (RF) analyses to compare individual diagnoses with our ANHAC group. Using RF is not essential to the ANHAC approach that we present here. In principle, this approach can use any form of discriminant analysis, or machine learning model. However, RF analyses are generally both robust and sensitive when analysing microbiome data.[20, 45] Moreover in the present context, RF models generated partial plots that could help to assess the influence of heteroscedasticity in the ANHAC approach (see above).

Our study focused on excluding reverse causation in associations between enteric dysbiosis and disorders that lack established pathology. However, there is increasing recognition that reverse causation can complicate many kinds of clinico-pathological associations that underpin our understanding of diseases (e.g. [46–53]). Methods in current use to exclude reverse causation may rely on unproven assumptions,[54] large samples,[55] long-term follow-up (e.g.[46, 56]) and/or complex statistical methods.[50, 56] In contrast, our method of constructing an ANHAC grouping uses clinical reasoning to help to exclude reverse causation due to illness-in-general. In the case of ME/CFS (where reverse causation due to definitive treatment is unlikely), our small study yielded findings comparable with those of complex Mendelian Randomisation analyses of a large sample. [43, 44] Such large samples are likely to include people with a wide range of illnesses, as well as a majority of ‘healthy controls’ – and so may already incorporate elements of our ANHAC approach. Potentially, (a) our ANHAC method may be applicable with other types of data that may carry a shared signature of illness-in-general – e.g. metabolomic or immunological measures – and (b) combining our ANHAC method with other strategies to exclude reverse causation may help to clarify causal mechanisms more efficiently in cross-sectional samples.

In summary, we present an approach to control for reverse causation of dysbiosis due to illness-in-general, and so help to define forms of dysbiosis that may contribute to causing specific disorders. We used the unique features of ME/CFS to illustrate our method, but did not expect our method to illuminate ME/CFS because our sample is so small. Nevertheless, correspondence between our findings and those of Mendelian Randomisation studies of microbiota that affect cognition[43, 44] supports the rationale of our approach. Ultimately, such causal knowledge may help to tailor FMT to treat ME/CFS.

## Supporting information

R code for random forest analysis

R code for analysis of Cao's dataset

R code for analysis of Gacesa's dataset

R code for heteroscedasticity analysis

Examples of decision trees of interaction between genera

AUROCs and partial plots

Dove_Atlas data

## Data Availability

All data produced in the present study are available upon reasonable request to the authors, or are already available online (as indicated in the text).

https://figshare.com/articles/dataset/Raw_data_for_microbiome_abondances_for_patients_1_Cancer_2_Chronic_Fatigue_syndrome_CFS_Irritable_bowel_Syndrome_IBS_4_Miscellaneous_6_CFS_/22762448

## Acknowledgements

IW received support from the Action for ME Foundation.

