## Supplementary information for "An approach to finding specific forms of dysbiosis that associate with different disorders"

**(1) ANHAC vs cancer; (2) heteroscedasticity; (3) comparison with independent studies**

### Introduction

ANHAC vs cancer: The logic of constructing an aggregate non-healthy active control (ANHAC) group to represent illness-in-general implies that we should not be able to discriminate different ANHAC groups. We tested this by comparing our ANHAC group with a grouping of patients who had cancers in several sites, representing a second ANHAC group. Cancer patients have dysbiosis.(1–5) Therefore, if our ANHAC and cancer groupings do not differ, then this would indicate that the ANHAC grouping is representative of illness in general or of reverse causation due to treatment.

Heteroscedasticity: The ANHAC group is heterogeneous by design. Hence, large variance of each genus (heteroscedasticity) in the ANHAC group could obscure differences between this group and single-disorder groups.

The reliability of our proposed method – comparing individual diagnostic categories with an ANHAC group – is uncertain, because no comparable reports exist. Therefore, in order to test our method's reliability, we used it to re-analyse secondary data from a large study.(4) We present these analyses in a separate section at the end of the Supplementary Information.

### Methods

All analyses used the open-source statistical programming language R. We provide the data and code to undertake all analyses (Supplementary files 1 & 2).

The abundances of different microbiota in each person's microbiome in our study are compositional data that sum to 100%. Two-thirds (66.6%) of abundances were zero. Troll and colleagues have shown that the machine-learning method “random forest” may be optimal for analysing raw compositional data that include zeros.(6) Here, therefore, we used random forest analyses.

The random forest (RF) analyses used the R package randomforestSRC.(7) We used a 2-stage procedure for each analysis. The first stage used each variable’s minimal depth to select those variables that may be relevant for discriminating the clinical groupings;(8) the second stage performed the discrimination. The variable selection stage grew 20000 classification trees and the discrimination stage grew 100000 trees. We did not formally ‘tune’ the random forest hyperparameters, but used small values to encourage robustness. Hence, the trees in both stages used 10 randomly-selected splits and tried 2 variables at each split to create a minimal node of size 1 at a maximal split depth of 5. The analyses down-sampled cases in the larger group, to reduce bias due to imbalance between group sizes.

The RF analyses provided the area under the receiver operating characteristic curve (AUROC) and overall error rate for classification. We also computed the 95% confidence limits of the AUROC, using R’s ROCR package.(9)

Random forest can naturally incorporate high-dimensional non-linear interactions. These are evident in the structure of decision trees (e.g. see Supplementary file ). The RF analyses also allowed assessment of whether interactions between genera may determine diagnostic categories. We assessed interactions using variable importance.(10)

**Results**

*Genera*

Table S1 shows the number of non-zero observations and the mean and variance of each genus in each diagnostic group. The mmglms found no differences between diagnostic groups.

| Diagnostic group: | ANHAC | | | IBS | | | ME/CFS | | | Cancer | | |
|---|---|---|---|---|---|---|---|---|---|---|---|---|
| Genus | N | Mean | Var | N | Mean | Var | N | Mean | Var | N | Mean | Var |
| Actinomyces | 6 | 0.00 | 0.00 | 8 | 0.00 | 0.00 | 9 | 0.01 | 0.00 | 7 | 0.01 | 0.00 |
| Adlercreutzia | 11 | 0.03 | 0.00 | 15 | 0.06 | 0.02 | 28 | 0.05 | 0.01 | 14 | 0.03 | 0.00 |
| Agathobacter | 17 | 1.22 | 3.42 | 21 | 0.65 | 0.67 | 36 | 1.42 | 2.83 | 27 | 1.01 | 1.29 |
| Akkermansia | 9 | 2.12 | 14.2 | 18 | 2.53 | 19.5 | 21 | 1.69 | 8.63 | 14 | 1.60 | 8.39 |
| Alistipes | 17 | 3.50 | 7.90 | 25 | 3.43 | 7.38 | 38 | 4.92 | 21.6 | 25 | 2.89 | 7.82 |
| Anaerostipes | 6 | 0.59 | 0.39 | 27 | 0.61 | 0.55 | 38 | 0.47 | 0.33 | 27 | 0.58 | 0.44 |
| Anaerotruncus | 3 | 0.01 | 0.00 | 12 | 0.01 | 0.00 | 10 | 0.01 | 0.00 | 8 | 0.02 | 0.00 |
| Bacteroides | 7 | 26.3 | 217.1 | 27 | 27.9 | 176.1 | 38 | 24.2 | 244.1 | 27 | 29.3 | 358.1 |
| Bacteroides.pectinophilus.group | 3 | 0.04 | 0.02 | 6 | 0.04 | 0.02 | 12 | 0.03 | 0.02 | 3 | 0.00 | 0.00 |
| Barnesiella | 11 | 1.42 | 3.98 | 17 | 0.89 | 1.10 | 30 | 1.05 | 0.98 | 16 | 1.09 | 1.59 |
| Bifidobacterium | 16 | 0.72 | 0.82 | 23 | 0.90 | 2.38 | 30 | 0.80 | 2.60 | 20 | 0.75 | 2.75 |
| Bilophila | 15 | 0.39 | 0.07 | 22 | 0.30 | 0.12 | 32 | 0.27 | 0.06 | 20 | 0.39 | 0.26 |
| Blautia | 17 | 1.30 | 0.90 | 27 | 1.52 | 1.98 | 38 | 1.63 | 2.78 | 27 | 1.45 | 2.45 |
| Butyricicoccus | 17 | 0.39 | 0.09 | 27 | 0.45 | 0.17 | 37 | 0.55 | 0.16 | 27 | 0.46 | 0.18 |
| Butyricimonas | 11 | 0.28 | 0.19 | 14 | 0.16 | 0.06 | 23 | 0.21 | 0.07 | 20 | 0.32 | 0.12 |
| Candidatus.Soleaferrea | 8 | 0.02 | 0.00 | 18 | 0.03 | 0.00 | 16 | 0.01 | 0.00 | 11 | 0.02 | 0.01 |
| Chloroplast.group.2 | 6 | 0.02 | 0.00 | 15 | 0.05 | 0.01 | 18 | 0.05 | 0.01 | 17 | 0.13 | 0.11 |
| Clostridium.innocuum.group | 6 | 0.01 | 0.00 | 7 | 0.01 | 0.00 | 3 | 0.00 | 0.00 | 6 | 0.01 | 0.00 |
| Clostridium.sensu.stricto.1 | 15 | 0.38 | 0.36 | 20 | 0.46 | 0.84 | 31 | 0.38 | 0.85 | 21 | 0.75 | 5.27 |
| Collinsella | 14 | 0.50 | 0.16 | 20 | 0.47 | 0.42 | 27 | 0.57 | 2.69 | 22 | 0.53 | 0.77 |
| Coprobacter | 12 | 0.14 | 0.09 | 16 | 0.19 | 0.14 | 20 | 0.08 | 0.02 | 19 | 0.35 | 0.21 |
| Coprococcus.3 | 15 | 0.21 | 0.07 | 18 | 0.14 | 0.05 | 36 | 0.18 | 0.03 | 22 | 0.16 | 0.03 |
| Desulfovibrio | 7 | 0.32 | 0.37 | 10 | 0.30 | 0.47 | 19 | 0.23 | 0.30 | 15 | 0.36 | 0.41 |
| Dialister | 14 | 1.19 | 4.79 | 11 | 0.38 | 1.08 | 18 | 0.21 | 0.10 | 14 | 0.58 | 0.63 |
| Dorea | 17 | 0.11 | 0.02 | 23 | 0.06 | 0.00 | 34 | 0.11 | 0.03 | 24 | 0.06 | 0.00 |
| DTU089 | 8 | 0.02 | 0.00 | 13 | 0.02 | 0.00 | 11 | 0.01 | 0.00 | 11 | 0.01 | 0.00 |
| Eggerthella | 4 | 0.01 | 0.00 | 11 | 0.01 | 0.00 | 10 | 0.02 | 0.01 | 7 | 0.02 | 0.00 |
| Eisenbergiella | 5 | 0.01 | 0.00 | 8 | 0.04 | 0.02 | 8 | 0.06 | 0.09 | 6 | 0.02 | 0.01 |
| Erysipelatoclostridium | 8 | 0.01 | 0.00 | 12 | 0.03 | 0.00 | 18 | 0.02 | 0.00 | 11 | 0.02 | 0.00 |
| Erysipelotrichaceae.UCG.003 | 17 | 0.16 | 0.02 | 20 | 0.13 | 0.02 | 33 | 0.15 | 0.03 | 20 | 0.12 | 0.03 |
| Escherichia.Shigella | 14 | 0.63 | 2.31 | 22 | 1.38 | 11.6 | 22 | 1.35 | 19.0 | 18 | 4.86 | 87.6 |
| Eubacterium.eligens.group | 15 | 0.83 | 1.37 | 23 | 0.59 | 0.48 | 32 | 0.67 | 0.63 | 20 | 0.58 | 0.55 |
| Eubacterium.hallii.group | 17 | 0.44 | 0.15 | 24 | 0.39 | 0.27 | 38 | 0.44 | 0.36 | 26 | 0.55 | 0.80 |
| Eubacterium.ruminantium.group | 2 | 0.02 | 0.01 | 8 | 0.14 | 0.12 | 10 | 0.45 | 1.59 | 3 | 0.02 | 0.01 |
| Faecalibacterium | 17 | 8.53 | 27.6 | 27 | 7.92 | 24.3 | 38 | 8.46 | 15.2 | 26 | 7.91 | 25.0 |
| Flavonifractor | 17 | 0.27 | 0.08 | 27 | 0.33 | 0.07 | 36 | 0.27 | 0.14 | 25 | 0.30 | 0.13 |
| Gordonibacter | 6 | 0.01 | 0.00 | 7 | 0.02 | 0.00 | 12 | 0.01 | 0.00 | 9 | 0.01 | 0.00 |
| Haemophilus | 9 | 0.12 | 0.04 | 19 | 0.19 | 0.13 | 23 | 0.13 | 0.07 | 18 | 0.37 | 1.56 |
| Holdemanella | 6 | 0.32 | 0.45 | 8 | 0.09 | 0.04 | 15 | 0.21 | 0.20 | 4 | 0.02 | 0.01 |
| Holdemania | 8 | 0.03 | 0.00 | 20 | 0.03 | 0.00 | 28 | 0.03 | 0.00 | 19 | 0.02 | 0.00 |
| Hungatella | 5 | 0.01 | 0.00 | 11 | 0.03 | 0.01 | 13 | 0.10 | 0.12 | 4 | 0.01 | 0.00 |
| Intestinibacter | 12 | 0.11 | 0.02 | 18 | 0.09 | 0.01 | 31 | 0.19 | 0.34 | 18 | 0.44 | 3.12 |

| | | | | | | | | | | | | |
|---|---|---|---|---|---|---|---|---|---|---|---|---|
| Intestinimonas | 0 | 0.03 | 0.00 | 16 | 0.06 | 0.01 | 19 | 0.03 | 0.00 | 12 | 0.03 | 0.00 |
| Lachnoclostridium | 16 | 0.71 | 0.24 | 26 | 0.66 | 0.47 | 38 | 0.85 | 2.08 | 26 | 0.60 | 0.30 |
| Lachnospira | 17 | 1.42 | 2.72 | 25 | 1.26 | 3.75 | 35 | 1.56 | 3.49 | 26 | 1.39 | 1.96 |
| Lachnospiraceae.UCG.001 | 12 | 0.39 | 0.53 | 19 | 0.36 | 0.23 | 29 | 0.64 | 0.58 | 20 | 0.34 | 0.25 |
| Lactobacillus | 5 | 0.01 | 0.00 | 9 | 0.01 | 0.00 | 7 | 0.10 | 0.34 | 6 | 0.11 | 0.23 |
| Methanobrevibacter | 3 | 0.03 | 0.01 | 4 | 0.19 | 0.43 | 13 | 0.16 | 0.24 | 8 | 0.03 | 0.00 |
| Mitsuokella | 10 | 0.06 | 0.01 | 16 | 0.12 | 0.06 | 32 | 0.17 | 0.17 | 17 | 0.04 | 0.00 |
| Mogibacterium | 0 | 0.00 | 0.00 | 10 | 0.05 | 0.03 | 13 | 0.02 | 0.00 | 8 | 0.01 | 0.00 |
| Muribaculum | 7 | 0.33 | 0.59 | 8 | 0.12 | 0.06 | 13 | 0.13 | 0.07 | 8 | 0.15 | 0.10 |
| Odoribacter | 16 | 0.39 | 0.06 | 23 | 0.40 | 0.11 | 36 | 0.54 | 0.14 | 24 | 0.45 | 0.14 |
| Olsenella | 9 | 0.06 | 0.01 | 6 | 0.03 | 0.00 | 9 | 0.02 | 0.00 | 7 | 0.01 | 0.00 |
| Oscillibacter | 17 | 0.84 | 0.18 | 26 | 0.81 | 0.42 | 38 | 1.07 | 0.48 | 26 | 0.83 | 0.35 |
| Oxalobacter | 6 | 0.04 | 0.00 | 9 | 0.02 | 0.00 | 15 | 0.04 | 0.01 | 7 | 0.01 | 0.00 |
| Papillibacter | 15 | 0.94 | 1.06 | 24 | 1.04 | 1.31 | 35 | 0.77 | 0.71 | 22 | 0.61 | 0.76 |
| Parabacteroides | 17 | 2.54 | 8.40 | 26 | 3.19 | 6.47 | 34 | 1.82 | 2.80 | 24 | 2.18 | 4.14 |
| Parasutterella | 4 | 0.54 | 1.01 | 14 | 0.42 | 0.70 | 31 | 0.75 | 0.86 | 14 | 0.41 | 0.76 |
| Phascolarctobacterium | 7 | 1.01 | 4.06 | 13 | 0.85 | 1.43 | 26 | 1.57 | 4.23 | 9 | 1.35 | 15.2 |
| Phocea | 6 | 0.01 | 0.00 | 9 | 0.02 | 0.00 | 18 | 0.04 | 0.04 | 5 | 0.04 | 0.03 |
| Prevotella.group.9 | 7 | 5.44 | 148.1 | 7 | 3.44 | 113.1 | 17 | 3.40 | 67.4 | 9 | 5.81 | 190.1 |
| Romboutsia | 16 | 0.19 | 0.04 | 18 | 0.16 | 0.08 | 30 | 0.20 | 0.09 | 18 | 0.32 | 1.32 |
| Roseburia | 17 | 1.10 | 0.66 | 25 | 1.67 | 3.46 | 36 | 2.21 | 3.75 | 27 | 1.44 | 2.22 |
| Rothia | 3 | 0.00 | 0.00 | 7 | 0.00 | 0.00 | 7 | 0.00 | 0.00 | 11 | 0.02 | 0.00 |
| Ruminiclostridium | 17 | 0.34 | 0.18 | 27 | 0.45 | 0.93 | 35 | 0.25 | 0.05 | 25 | 0.40 | 0.63 |
| Ruminiclostridium.group.1 | 5 | 0.05 | 0.03 | 7 | 0.02 | 0.00 | 16 | 0.03 | 0.00 | 5 | 0.01 | 0.00 |
| Ruminiclostridium.group.6 | 12 | 0.31 | 0.27 | 17 | 0.48 | 1.55 | 30 | 0.60 | 0.93 | 16 | 0.50 | 1.07 |
| Ruminiclostridium.group.9 | 17 | 0.73 | 2.83 | 26 | 0.51 | 0.51 | 36 | 0.71 | 1.39 | 24 | 0.20 | 0.05 |
| Ruminococcaceae.UCG.002 | 15 | 3.04 | 7.56 | 24 | 1.35 | 3.66 | 35 | 2.13 | 2.81 | 20 | 1.40 | 2.99 |
| Ruminococcus.group.1 | 14 | 0.57 | 1.23 | 22 | 1.09 | 1.85 | 31 | 0.67 | 0.97 | 17 | 0.19 | 0.13 |
| Ruminococcus.group.2 | 10 | 0.58 | 0.88 | 20 | 0.80 | 1.04 | 29 | 0.71 | 0.55 | 16 | 0.46 | 0.82 |
| Ruminococcus.Lachnospiraceae | 17 | 0.79 | 0.48 | 27 | 0.74 | 0.43 | 38 | 0.72 | 0.53 | 27 | 0.87 | 0.85 |
| Senegalimassilia | 4 | 0.02 | 0.00 | 6 | 0.02 | 0.00 | 13 | 0.03 | 0.00 | 9 | 0.02 | 0.00 |
| Slackia | 3 | 0.02 | 0.00 | 8 | 0.01 | 0.00 | 14 | 0.03 | 0.00 | 8 | 0.02 | 0.00 |
| Streptococcus | 15 | 0.09 | 0.01 | 25 | 0.34 | 0.50 | 33 | 0.76 | 13.4 | 24 | 0.29 | 0.36 |
| Sutterella | 11 | 1.63 | 3.24 | 17 | 1.33 | 4.68 | 23 | 1.01 | 2.26 | 15 | 1.24 | 2.91 |
| Terrisporobacter | 8 | 0.07 | 0.01 | 12 | 0.05 | 0.02 | 21 | 0.11 | 0.06 | 12 | 0.06 | 0.01 |
| Turicibacter | 7 | 0.04 | 0.00 | 14 | 0.12 | 0.10 | 21 | 0.09 | 0.04 | 16 | 0.12 | 0.06 |
| Tyzzerella | 14 | 0.15 | 0.04 | 24 | 0.13 | 0.01 | 35 | 0.17 | 0.04 | 25 | 0.20 | 0.07 |
| UBA1819 | 17 | 0.55 | 1.68 | 21 | 0.28 | 0.18 | 33 | 0.19 | 0.13 | 22 | 0.19 | 0.23 |
| Unknown.Bacteria | 15 | 1.54 | 5.74 | 26 | 1.96 | 4.70 | 37 | 2.61 | 6.28 | 23 | 1.97 | 9.08 |
| Unknown.Bacteroidales | 7 | 0.85 | 4.02 | 11 | 0.52 | 1.62 | 18 | 0.62 | 2.16 | 15 | 0.30 | 0.29 |
| Unknown.Christensenellaceae | 8 | 0.02 | 0.00 | 12 | 0.03 | 0.00 | 27 | 0.03 | 0.00 | 9 | 0.01 | 0.00 |
| Unknown.Clostridiales | 14 | 1.70 | 4.87 | 23 | 1.85 | 3.86 | 35 | 1.52 | 2.81 | 24 | 0.88 | 1.23 |
| Unknown.Coriobacteriales | 12 | 0.05 | 0.00 | 14 | 0.06 | 0.03 | 23 | 0.11 | 0.20 | 9 | 0.03 | 0.00 |
| Unknown.Enterobacteriaceae | 3 | 0.42 | 2.86 | 6 | 1.69 | 36.0 | 13 | 0.12 | 0.13 | 14 | 1.40 | 37.3 |
| Unknown.Erysipelotrichaceae | 8 | 0.01 | 0.00 | 13 | 0.11 | 0.06 | 13 | 0.05 | 0.04 | 10 | 0.05 | 0.02 |
| Unknown.Firmicutes | 17 | 3.39 | 14.8 | 26 | 3.37 | 22.2 | 37 | 3.75 | 17.4 | 25 | 1.46 | 4.43 |
| Unknown.Lachnospiraceae | 17 | 6.93 | 15.3 | 27 | 6.89 | 12.0 | 38 | 8.52 | 13.9 | 27 | 5.89 | 11.9 |
| Unknown.Prevotellaceae | 3 | 2.43 | 68.6 | 6 | 1.21 | 7.94 | 8 | 0.76 | 7.65 | 5 | 2.10 | 34.8 |
| Unknown.Ruminococcaceae | 17 | 2.92 | 9.17 | 25 | 2.79 | 9.40 | 38 | 3.35 | 7.02 | 26 | 2.09 | 4.86 |
| Veillonella | 15 | 0.10 | 0.03 | 22 | 0.64 | 7.86 | 30 | 0.26 | 0.74 | 23 | 0.36 | 0.48 |
| Victivallis | 9 | 0.05 | 0.00 | 5 | 0.02 | 0.01 | 18 | 0.06 | 0.01 | 7 | 0.04 | 0.02 |

Legend: the genus-wise mean proportions (%) in each person’s microbiome; variances include zeros

*ANHAC vs cancer groupings*

Twenty-seven participants had cancer (12 breast, 6 colorectal, 4 prostate, 2 melanoma, 3 others). The random forest analyses could not discriminate the ANHAC and cancer groupings (AUROC = 61.0%, 95% CI = 44.1 – 77.9; overall error rate = 38.6%).

*Heteroscedasticity*

Non-parametric tests: The proportion of zero observations was lower in 69/95 individual genera in the ANHAC group than in the grouping of cancer patients (binomial p=0.001) or in 61/95 genera in people with IBS (binomial p=0.007), but did not differ from people with ME/CFS (49/95 genera, binomial p=0.84). The variances of different genera did not differ, overall, between the ANHAC group and the single-disorder groupings (all paired WMW p>0.25) (not shown).

*Random forest models*

The file "decision tree examples 040424.pdf" shows examples of decision trees for the discrimination between the ANHAC and ME/CFS groups.

**Discussion**

The present study found that random forest analyses could discriminate single disorders from an aggregate non-healthy control (ANHAC) group. The main report discusses the potential importance of these discriminations. Here, their importance is that they demonstrate that heteroscedasticity of the ANHAC group may not obscure all differences from single-disorder groups.

**1) ANHAC vs cancer**

Random forest analyses could not discriminate the ANHAC and cancer groupings. This negative result is consistent with the view that the cancer grouping itself represents an ANHAC group, since the cancer grouping included patients who had different kinds of cancer in different sites and may have received a range of different treatments. However, an alternative explanation is that the cancer group was, overall, older than the ANHAC group. Hence, if the relative abundances of different genera depend on age, this dependence could potentially obscure further associations between those genera and the cancer grouping. We address the question whether mismatches between the ANHAC group and single disorders may seriously bias the findings of our method below. For now, we note that (a) in principle, random forest analyses can account for such mismatches, because they can natively incorporate interactions between the confounding mismatched factors and potentially-predictive genera; (b) in practice, neither age nor sex contributed importantly to discriminating IBS or ME/CFS from our ANHAC grouping.

**2) Heteroscedasticity**

The proportion of observations of zero abundance of individual genera was lower in the ANHAC group than in the cancer and IBS groups. In one sense, this indicates that the ANHAC group had a more varied  microbiome, which is consistent with the *a priori* reasoning that it should be more heteroscedastic. On the other hand, the variances of individual genera did not differ between the ANHAC grouping and single disorders, so that mathematical heteroscedasticity was absent. In short, the ANHAC group showed greater biological diversity, but less mathematical heteroscedasticity.

Defining heteroscedasticity in relation to dysbiosis is difficult. Many studies have defined dysbiosis by comparing diversity indices of microbiota within individuals.(11–13) This approach assumes that each person is fundamentally similar, and that patterns of microbiota within each person can constitute potentially-causal dysbiosis. Conversely, it is equally possible that each microbe is fundamentally similar but that people differ, so that varying abundances of each microbe across different people reflect possibly-pathogenic causal effects of each person's enteric environment. Potentially, mixed-membership generalised linear models may be able to assess heteroscedasticity while incorporating both of these perspectives.

The principal limitation of the present study are its small size – especially the small size of the ANHAC group. However, we analysed publicly-available data based on a much larger sample size, in order to assess the reliability of the present results (see below). A second limitation is the fact that two-thirds (66.6%) of abundance data were zero. These may be (i) true zeros, (ii) below the limits of detection (BLD), or (iii) result from rounding. If many BLD or rounding zero values were present, then this could limit the sensitivity of the random forest analyses – which implies that we may have under-estimated our method's ability discriminate the ANHAC group from individual disorders. A third limitation is that available abundances of different microbiota in each person's microbiome in our study are compositional data that sum to 100%. Analyses of compositional data may benefit from special statistical methods. However, the random forest analyses that we used are non-parametric and so may be relatively insensitive to compositional data.

#### 3) Testing the reliability of our method by comparing it with previous studies' findings

#### Introduction

No previous studies have compared an aggregate non-healthy active control (ANHAC) group with individual disorders. Therefore, in order to test the reliability of our method, we analysed publicly-available data from two previous reports of large sample. First, Gacesa and colleagues(4) presented effect sizes of different genera on medical conditions or symptoms in 8208 people (see Supplementary Table S3b in(4)). Second, Cao and colleagues used Mendelian Randomisation analyses of a very large sample (¼ million) to assess causal effects of individual genera on cognitive function.

#### Method – Gacesa comparison

We compared the cancer (any, non-basal-cell) and irritable bowel syndrome (Rome-3 criteria – any type) groups from Gacesa's study with those from our study. We did not include ME/CFS, because (a) the comparison requires many genera and (b) both named genera that discriminated our ME/CFS group from our ANHAC grouping also contributed to discriminating our IBS and ANHAC groups.

Gacesa's Supplementary Table S3b includes relations between 50 genera and 252 phenotypes, in 8208 people aged 8-84. The phenotypes included 81 medical disorders with at least 20 cases in the total sample of 8028 people. The relations are effect sizes for partial associations of each genus with each phenotype, adjusted for all other genera and phenotypes and for basic demographic data.

In order to compare Gacesa's data with our findings, we used Gacesa's data to construct a new ANHAC group. This new ANHAC group that comprised 28 medical disorders or diagnostic groupings with a total group size probably in excess of 2500 (see Table S2 – the precise size cannot be calculated from the available data). We then computed the mean effect size for each genus, across these 28 disorders. These mean effect sizes represent the shared microbiome signature of illness-in-general and deviations from this signature in specific disorders may reflect specific forms of dysbiosis that associate uniquely with those disorders.

We assessed the similarities and differences between the ANHAC group, the Healthy Controls and the clinical groups (IBS and cancer) in Gacesa's data using robust linear regression with a 50% breakdown point.(14) Robust regression is resistant to leverage by outliers and so can separate overall tendencies from individual outlier 'effects' (which is important for the analysis – see below).

We first tested if the genus-wise mean effect sizes for the ANHAC group demonstrate a "shared microbiome signature of disease".(4) If so, then the ANHAC group's effect sizes should relate *inversely* to those of the Healthy Control group ("MED.DISEASES.None.No.Diseases") (cf Fig. 4a in(4)).

Subsequently, we tested how far the profiles of the genus-wise mean effect sizes for IBS and cancer resemble the ANHAC group in Gacesa's data. If IBS and cancer share the dysbiosis due to illness-in-general, then their genus-wise effect sizes should relate *directly* to those of the ANHAC group.

Finally, we tested if *deviations* from the "shared microbiome signature of disease" in Gacesa's data correspond with the genera that discriminated IBS and cancer in our study. (We could not do this for ME/CFS, because only 2 known genera discriminated CFS and the test needs at least 3 genera). To do this, using Gacesa's data, we extracted the residuals from the robust regressions (see above), then estimated simple variance ratios to test if, overall, the variance of these residuals was greater in the genera that discriminated the clinical groups in our own data. This variance represents departures from the shared microbiome signature of disease in Gacesa's data that contributes to discriminating the ANAHC group from individual disorders in the random forest analyses of our own data-set.

**Method – Cao comparison**

Cao's Supplementary Table S2 includes MR-derived estimates of causal effects of 119 genera on cognition in a sample of 257841 people. Cao reported that 7 genera had significant causal effects on cognition – two of which corresponded with genera that predicted ME/CFS in our study. We calculated the probability of this degree of concordance directly using a permutation procedure.

**Results – Gacesa comparison**

Table S1: The disorders from Gacesa's data that we analysed:-

| **Diagnostic group / disorder** | N |
|---|---|
| Healthy Controls – MED.DISEASES.None.No.Diseases | 1876 |
| MED.DISEASES.Gastrointestinal.Rome3_IBS.Any | 650 |
| MED.DISEASES.Cancer.AnyNonBasal | 300 |
| **Aggregate Non-Healthy Active Control (ANHAC) grouping** | |
| MED.DISEASES.Blood.Anemia | 1189 |
| MED.DISEASES.Cardiovascular.Atherosclerosis | 36 |
| MED.DISEASES.Cardiovascular.Hypertension | 1711 |
| MED.DISEASES.Endocrine.Autoimmune.DiabetesT1 | 25 |
| MED.DISEASES.Endocrine.DiabetesT2 | 181 |
| MED.DISEASES.Gastrointestinal.Autoimmune.Celiac | 50 |
| MED.DISEASES.Gastrointestinal.Autoimmune.IBD.CD | 28 |
| MED.DISEASES.Gastrointestinal.Autoimmune.IBD.UC | 71 |
| MED.DISEASES.Gastrointestinal.Stomach.Ulcer | 303 |
| MED.DISEASES.Hepatologic.Gallstones | 353 |
| MED.DISEASES.Hepatologic.Hepatitis | 26 |
| MED.DISEASES.Mental.Any | 1145 |
| MED.DISEASES.Neurological.Autoimmune.Multiple.Sclerosis | 59 |
| MED.DISEASES.Neurological.Epilepsy | 83 |
| MED.DISEASES.Neurological.Migraine | 1412 |
| MED.DISEASES.Neurological.Stroke | 70 |
| MED.DISEASES.Other.Autoimmune.Rheumatoid.Artritis | 163 |
| MED.DISEASES.Other.Fractures.Hip | 67 |
| MED.DISEASES.Other.Fractures.Other | 171 |
| MED.DISEASES.Other.Kidney.Stones | 101 |
| MED.DISEASES.Other.Osteoarthritis | 1020 |
| MED.DISEASES.Other.Osteoporosis | 156 |
| MED.DISEASES.Pulmonary.Autoimmune.Asthma | 382 |

| | |
|---|---|
| MED.DISEASES.Pulmonary.COPD | 263 |
| MED.DISEASES.Pulmonary.Pulmonary.Embolism | 44 |
| MED.DISEASES.Skin.Autoimmune.Atopic.dermatitis | 1210 |
| MED.DISEASES.Skin.Autoimmune.Psoriasis | 218 |
| MED.DISEASES.Skin.Autoimmune.Severe.acne | 211 |

Legend: The table shows the name of each diagnostic group or disorder in Gacesa's Table S3b and number of people with that disorder. Note that co-morbidity is possible, so that the total N does not sum to the total number of people in |Gacesa's report (8208).

<u>Table S2:</u> The genera that we analysed and their effect sizes on different diagnostic groupings:-

| | ANHAC | IBS | ME/CFS | Cancer |
|---|---|---|---|---|
| Actinomyces | 0.01 | 0.09 | -0.17 | 0.01 |
| Adlercreutzia | 0.15 | -0.02 | -0.32 | -0.04 |
| Akkermansia | -0.28 | -0.37 | 0.05 | -0.04 |
| Alistipes | 0.00 | -0.08 | 0.01 | 0.06 |
| Anaerostipes | 0.02 | 0.14 | -0.20 | 0.18 |
| Anaerotruncus | 0.29 | 0.56 | 0.66 | 0.31 |
| Bacteroides | 0.09 | 0.25 | 0.20 | 0.11 |
| Barnesiella | -0.20 | -0.31 | -1.61 | -0.39 |
| Bifidobacterium | -0.25 | -0.55 | -1.21 | 0.02 |
| Bilophila | -0.16 | -0.15 | -0.88 | -0.36 |
| Blautia | 0.06 | 0.00 | 0.71 | -0.02 |
| Collinsella | -0.16 | -0.30 | -0.38 | -0.09 |
| Coprobacter | 0.10 | -0.22 | -0.29 | 0.17 |
| Desulfovibrio | -0.32 | -0.77 | -0.50 | -0.72 |
| Dialister | 0.02 | 0.25 | -1.51 | -0.38 |
| Dorea | -0.23 | -0.12 | -0.73 | -0.21 |
| Eggerthella | 0.34 | 0.66 | 0.96 | 0.28 |
| Erysipelotrichaceae_NOS | -0.02 | -0.34 | -0.20 | 0.42 |
| Escherichia | 0.14 | 0.01 | 0.96 | 0.33 |
| Eubacterium | -0.11 | -0.15 | -0.07 | 0.02 |
| Faecalibacterium | -0.13 | -0.27 | -0.03 | -0.07 |
| Flavonifractor | 0.49 | 0.59 | 1.34 | 0.76 |
| Gordonibacter | 0.05 | 0.34 | 0.48 | 0.18 |
| Haemophilus | 0.05 | 0.03 | 0.16 | -0.50 |
| Holdemania | 0.29 | 0.80 | 0.62 | 0.48 |
| Lactobacillus | -0.04 | -0.23 | -0.47 | -0.28 |
| Methanobrevibacter | -0.08 | -0.10 | 0.19 | -0.22 |
| Mitsuokella | -0.22 | -0.43 | -0.67 | -0.08 |
| Odoribacter | -0.02 | -0.15 | -0.21 | -0.14 |
| Oscillibacter | 0.11 | 0.11 | 0.15 | 0.17 |
| Oxalobacter | -0.27 | -0.49 | 0.22 | -0.31 |
| Parabacteroides | -0.02 | -0.12 | 0.28 | -0.08 |
| Parasutterella | 0.01 | -0.24 | -1.14 | 0.39 |
| Phascolarctobacterium | -0.20 | -0.72 | 1.46 | -0.02 |
| Prevotella | -0.29 | -0.45 | 1.34 | -0.25 |
| Roseburia | -0.07 | -0.08 | 0.01 | -0.12 |
| Rothia | -0.08 | -0.05 | -0.40 | -0.51 |
| Ruminococcaceae_NOS | 0.31 | 0.35 | 0.01 | 0.77 |

| | | | | |
|---|---|---|---|---|
| Ruminococcus | -0.24 | -0.51 | 0.14 | -0.04 |
| Streptococcus | 0.12 | 0.05 | 0.21 | -0.17 |
| Sutterella | -0.29 | -0.41 | -0.72 | -0.12 |
| Bacteroidales_NOS | -0.15 | -0.39 | -0.60 | -0.05 |
| Clostridiales_NOS | 0.02 | -0.02 | -0.03 | 0.01 |
| Lachnospiraceae_NOS | 0.05 | 0.12 | 0.20 | -0.04 |
| Veillonella | 0.13 | 0.43 | 0.38 | -0.14 |

Legend: the name of each genus and its effect size in each diagnostic group, from table S3b of Gacesa's report. Values for the ANHAC group are means of effect sizes for the 28 disorders that we chose to represent this group; values for the IBS, ME/CFS and cancer groups are those provided by Gacesa. We substitute 'NOS' (not otherwise specified) for 'unclassified' in Gacesa's table. Note that the largest single effect size for Dialister indexes its association with ME/CFS; this corresponds with our result, but is not quite significant here (p~0.06 – see Gacesa's Supplementary Table S3b).

The ANHAC group showed a "shared microbiome signature of disease"(4) (Fig S1). Specifically, the genus-wise mean effect sizes of the ANHAC group related strongly and *inversely* to those of the same genera in the Healthy Controls (b=-0.80 ± 0.10, t = -8.26, p<0.001; adjusted $R^2$ = 61.9%).

Figure S1: The relation between genus-wise effect sizes of Healthy Controls and the ANHAC group

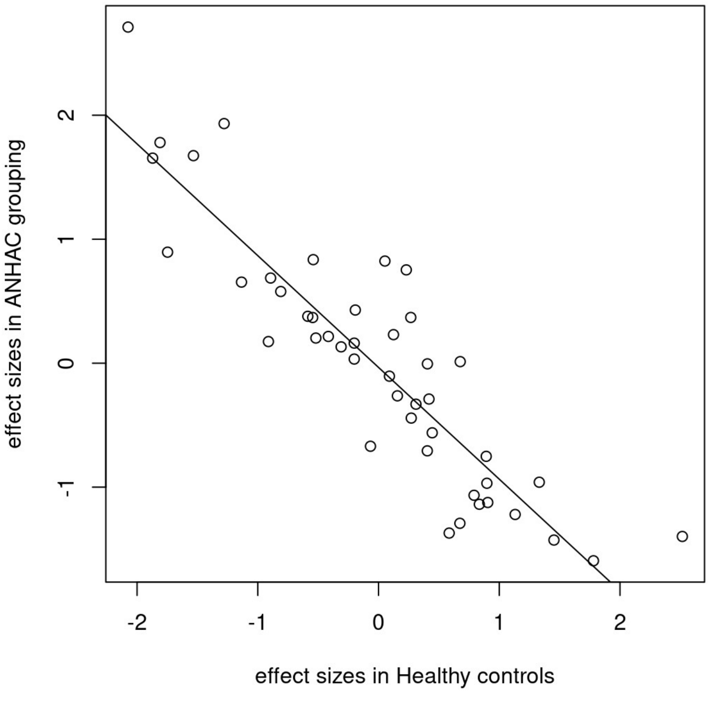


Legend: the figure shows the mean effect sizes for the ANHAC group (y-axis) and the corresponding values for the Healthy Controls (y-axis), together with the robust regression line. The inverse correlation here parallels the results in Figure 4a in Gacesa's main report.(4)

Gacesa's IBS group showed the "shared microbiome signature of disease" (Fig. S2). Specifically, the genus-wise effect sizes of the IBS group related strongly and *directly* to those of the ANHAC group ($b=0.87 \pm 0.09$, $t = 9.94$, $p<0.001$; adjusted $R^2 = 76.2\%$) and there was no additional relationship to the Healthy Controls ($\chi^2 = 0.01$, 1df, $p=0.92$).

Figure S2: the relation between genus-wise effect sizes in the ANHAC and IBS groups in(4)

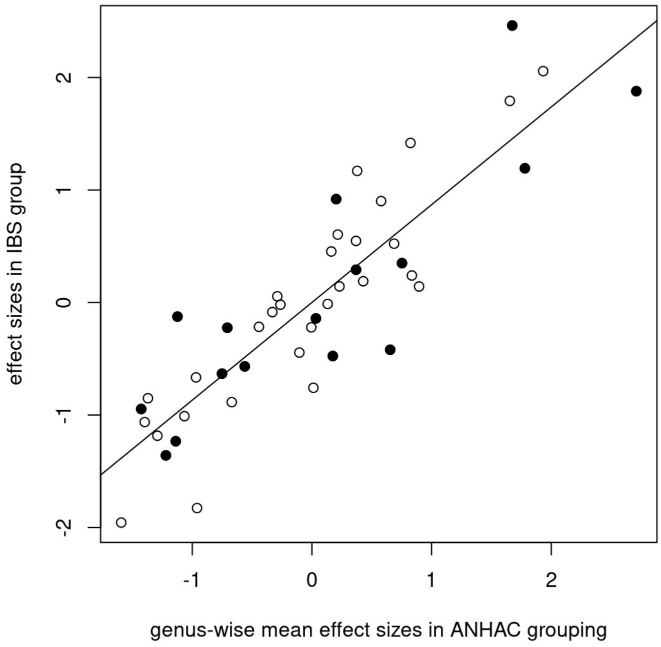


Legend: the effect size for each genus for the IBS group (y-axis) over the mean effect sizes in the ANHAC grouping (x-axis). Bullets are genera that discriminated the ANHAC and IBS groups in our study; open circles are genera that did not discriminate these groups.

Gacesa's cancer grouping showed the "shared microbiome signature of disease" (Fig. S3). Specifically, the genus-wise effect sizes of the cancer grouping related strongly and directly to those of the ANHAC group ($b = 0.66 \pm 0.12$, $t = 5.29$, $p<0.001$; adjusted $R^2 = 43.7\%$) and there was no additional relationship to the Healthy Controls ($\chi^2 = 1.16$, 1df, $p=0.28$).

Figure S3: the relation between genus-wise effect sizes in cancer and ANHAC groups in(4)

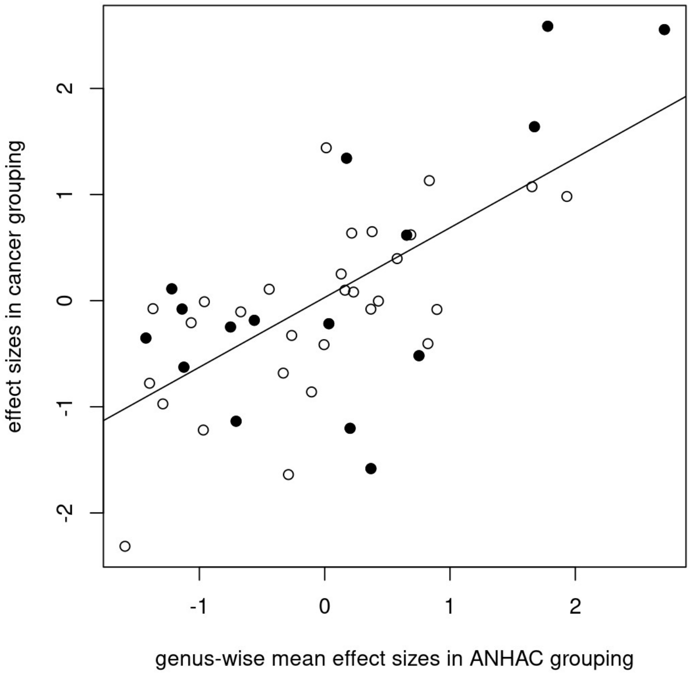


Legend: the effect size for each genus for the cancer grouping (y-axis) over the mean effect sizes in the ANHAC grouping (x-axis). Bullets are genera that random forest selected to discriminate the ANHAC and cancer groups in our study; open circles are unselected genera.

Genera that showed greater departures from the "shared microbiome signature of disease" in Gacesa's data corresponded with those that discriminated the clinical groupings in our data (see Fig. S4). Specifically, the variance of the genus-wise residuals from the robust regressions of Gacesa's data (see above) was larger for genera that discriminated the clinical groupings in our data than the variance of the residuals from genera that did not discriminate the clinical groupings (F = 3.13, 19/24df, p=0.005). These residuals represent variation that is not part of the "shared microbiome signature of disease", but does contribute to discriminate clinical groupings – so that it represents effects of genera that may be specific to individual disorders.

Figure S4: the distributions of residuals from robust analyses of Gacesa's data (see figures 2-3, above) according to their selection to discriminate the IBS and cancer groups in our original study

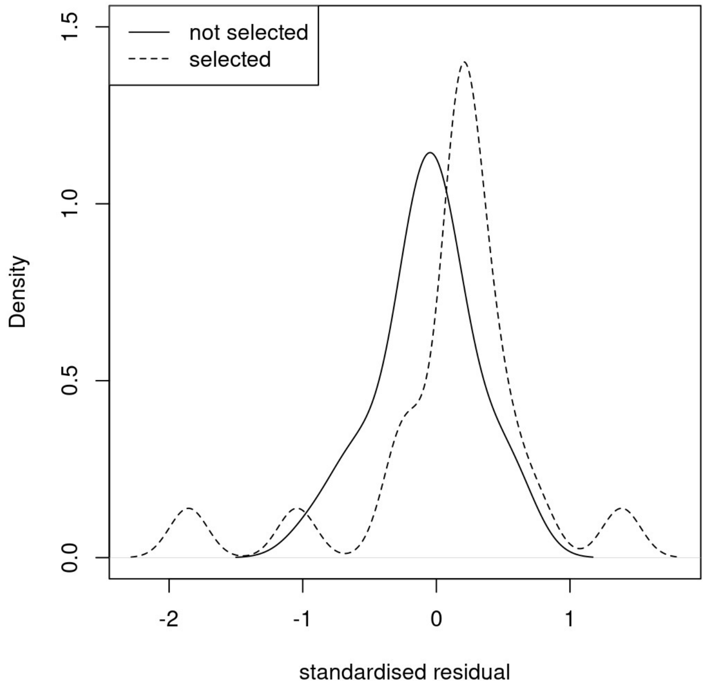


Legend: The distribution of residuals from the robust regression of effect sizes of individual genera on single disorders on mean effect sizes of genera in the ANHAC group in *Gacesa's* data, according to their selection (by random forest) to discriminate IBS or cancer from the ANHAC grouping in *our* original data. The variance of the genera that the random forest selected was greater than that of the remaining genera, indicating that effects that are not part of the shared signature of illness-in-general may contribute to individual disorders.

**Results – Cao comparison**

The probability that by chance alone Cao's Mendelian Randomisation study would detect 2 of the 3 genera (Dialister and Roseburia) that predicted ME/CFS in our random forest analyses was p=0.0045.

### Discussion

*General conclusion*

Genera that could discriminate the ANHAC group and individual disorders in our small study corresponded with those that showed greater departures from the "shared microbiome signature of disease" in Gacesa's large independent data-set.(4) Additionally, there was good correspondence between our findings and those of Cao's Mendelian Randomisation study to determine genera that have causal effects on cognition. These convergent effects indicate that our method may reliably detect forms of dysbiosis that are not secondary to illness-in-general, but relate to specific features of individual disorders.

*Gacesa*

The effect sizes of individual genera in the ANHAC group that we created using Gacesa's data related *inversely* to the effect sizes of the genera in their Healthy Controls. This inverse relationship is consistent with Gacesa's conclusion that there is a "shared microbiome signature of disease".(4) The most likely explanation of such a shared signature is reverse causation, due to non-specific effects of illness-in-general on the microbiome – effects that are absent in the Healthy Controls.

The effect sizes of individual genera in the ANHAC group that we created using Gacesa's data related *directly* to the effect sizes of the genera in their IBS and cancer groupings. These direct relationships further support Gacesa's conclusion that there is a "shared microbiome signature of disease".(4) Since the IBS and cancer groupings share elements of illness-in-general, their microbiome signatures have many elements in common with that of the ANHAC group.

The residuals from the robust regression represent departures from the overall shared signature of illness-in-general in Gacesa's data that might reflect effects of specific genera on individual disorders. Consistent with this, genera that show greater residuals in the robust regression of Gacesa's data corresponded with genera that random forest analyses selected to discriminate the ANHAC group from IBS and cancer in our small study. This correspondence indicates that our study's method may be reliable and its logic may be valid.

*Correspondence with Cao's Mendelian Randomisation results*

There was good correspondence between our findings that Dialister and Roseburia could predict ME/CFS and those of Cao, Wang and their colleagues that these genera may *cause* cognitive impairment.(15,16) In fact, the simple permutation analysis that we used to assess this correspondence under-estimated both its statistical significance and surprise factor, because the analysis did not take into account the direction of each genus's effect. In contrast to our results, He and collegaues(17) found that Paraprevotella and Ruminococcaceae.UCG.014 showed possible causal associations with ME/CFS in ½ million people. However, these associations were tiny and, *contra* He's finding but in line with ours, Gacesa found that low Dialister associates almost significantly with ME/CFS (p=0.069).(4) The reason why our findings align better with those of Cao and Wang than with those of He's report is uncertain. In contrast with the simplicity of our approach, the complexity of MR methods is such that even when analysing the same large sample, differences in the analytical method for computing MR can yield different results. Cao and Wang's studies analysed the same sample, and found a total of 11 genera that may cause ME/CFS, but only 5/11 genera were in common between the two studies. One of these five genera – Paraprevotella – also showed a tiny causal effect on ME/CFS in He's study(17) – which fits the notion that genera that can cause cognitive impairment may also contribute to ME/CFS.

*General considerations*

We constructed ANHAC groups (using Gacesa's data and our own) that had a wide range of pathologies, symptoms and treatments. Such groups may be suitable for representing broad secondary effects of illness-in-general on the microbiome. However, it may be possible to test causal roles of specific forms of dysbiosis for specific features of individual (target) disorders, by constructing ANHAC groups that match all features the target disorders, except the specific feature(s) of interest. For example, to test if specific forms of dysbiosis contribute to diarrhoea in IBS, it may be optimal to construct an ANHAC group that includes only disorders with different pathologies that cause diarrhoea or constipation (e.g. Crohn's disease, Ulcerative Colitis, hyperthyroidism, carcinoid tumours of the gastrointestinal tract, Hirschsprung's disease, etc.).

Our sample was too small to allow us to construct an ANHAC group whose features matched a single target disorder. Although random forest can potentially account for mismatch (see above), this may be more difficult with other forms of analysis. Therefore, our approach ideally needs a way to reject dysbiosis-disorder associations that result from inadequate matching. To this end, we experimented with permuting links between microbiome measures and diagnostic group (ANHAC or the single disorder), while maintaining constant links between clinical features and each individual (see details in Methods). This permutation approach should detect only specific forms of dysbiosis that discriminate the ANAHC group and target disorder(s) over-and-above any clinical contrasts between the groups. However, in practice, the permutation analyses were negatively biased in discriminating the ANHAC group from single disorders. This may reflect the disruption of associations between the demographic factors and microbial genera. Further studies should devise methods to eliminate this negative bias.

**List of Supplementary Files**

1) The data that we used in our original study (“Dove_Atlas data 040424.csv”)

2) The R code for the random forest analyses in our study (“random forest analyses 070424.txt”)

3) The R code for the analyses of heteroscedasticity in our study (“heteroscedasticity analyses 040424.txt”)

4) Gacesa’s Supplementary Table S3b is readily available at:-

https://static-content.springer.com/esm/art%3A10.1038%2Fs41586-022-04567-7/MediaObjects/41586_2022_4567_MOESM4_ESM.xlsx

We downloaded this an converted it to a comma-separated values (csv) file - "Gacesa_S3b.csv'

5) The R code that we used to analyse the data from Gacesa’s study and relate it to our own (“Gacesa_prg1_030424.txt”)

6) The R code that we used to analyse the data from Cao’s study and relate it to our own (“Cao_prg1_200424.txt”)

7) The Receiver Operating Characteristic curves and partial plots from the Random Forest analyses (“AUROCs and partial plots Mon Apr  1.pdf”)

8) Example decision trees of interactions between genera (“decision tree examples 040424.pdf”)

8. Ishwaran H, Chen X, Minn A, Lu M, Lauer M, Kogalur UB. randomForestSRC: minimal depth vignette. [Internet]. Available from: http://randomforestsrc.org/articles/minidep.html
