## Supplementary material for "An approach to finding specific forms of dysbiosis that associate with different disorders": R code for random forest analysis

```
######################################################################
#= START OF R CODE TO ANALYSE DATA ===================================#
#= USING RANDOM FOREST, FOR MAIN REPORT ==============================#
#====================================================================#
#====================================================================#
#==NOTE THAT THE MAIN OUTPUT FILE FROM ===============================#
#==THIS CODE MAY CRASH 'GEDIT' TEXT EDITOR ===========================#
#==BUT IS OPENABLE USING LIBREOFFICE =================================#
#====================================================================#
#====================================================================#
######################################################################

# read in data
mydir='/home/jonathan/Desktop/Atlas/'
bdat=read.csv(paste0(mydir,'Dove_Atlas data 200424.csv'))
# check bdat structure
dim(bdat); names(bdat)[1:7]; table(bdat$dgn)
str(bdat); bdat[1:5,1:15]
# id is patient number; age is years; sex is 0=F, 1=M
# dgn is 1=cancer, 2=IBS, 4=ANHAC, 6=CFS
# ibs is presence of IBS; cfs is presence of ME/CFS
# all patients have ill=1 (yes)
#  id age sex dgn ibs cfs ill Actinomyces Adlercreutzia Agathobacter Akkermansia
Alistipes
#1  1  77   0   2   1   0   1        0.02          0.00         0.95        0.02
3.36
#2  2  81   0   1   0   0   1        0.05          0.04         1.01        2.03
7.51
#3  3  75   1   4   0   0   1        0.00          0.00         0.01        0.58
3.36
#4  4  85   0   4   0   0   1        0.01          0.06         0.88        7.87
4.25
#5  5  50   0   2   1   1   1        0.00          0.07         0.04        9.61
3.53

# change all ages to quinquennia for purposes of publication (true ages
available on reasonable request)
# bdat$age=round(bdat$age/5)

# raretax is the sum of proportions of genera with more than 80% zero
observations
all(apply(bdat[,7:100],1,sum)==100) # TRUE
# pathgen is the sum of proportions of genera that Atlas defines as 'pathogenic'
and is unused here

# function to find random random-number seeds
jrnd=function(x)
return(round(as.numeric(substr(as.character(((x+10)^.9)*1e12),6,12))))
# function to count number of zeros in a vector
nzs=function(x) return(length(x[x==0]))
# function to return shortened version of date()
mydate=function() return(substr(date(),1,10))


# basic demographic results
table(bdat$sex,bdat$dgn); tapply(bdat$age,bdat$dgn,quantile)

# supplementary table S1
if (!exists('ngens')) ngens=names(bdat)[7:100] # omit pathgen
genns=names(bdat)%in%ngens; names(bdat)[genns] # also make T/F marker of genera
to be used
gres=array(NA,c(length(ngens),12),dimnames=list(ngens,rep(c('n','mean','var'),4)
))
gres[,1]=length(bdat$dgn[bdat$dgn==4])-apply(bdat[bdat$dgn==4,ngens],2,nzs)
```

```
gres[,2]=round(apply(bdat[bdat$dgn==4,ngens],2,mean),2)
gres[,3]=round(apply(bdat[bdat$dgn==4,ngens],2,var),2)
gres[,4]=length(bdat$dgn[bdat$dgn==2])-apply(bdat[bdat$dgn==2,ngens],2,nzs)
gres[,5]=round(apply(bdat[bdat$dgn==2,ngens],2,mean),2)
gres[,6]=round(apply(bdat[bdat$dgn==2,ngens],2,var),2)
gres[,7]=length(bdat$dgn[bdat$dgn==6])-apply(bdat[bdat$dgn==6,ngens],2,nzs)
gres[,8]=round(apply(bdat[bdat$dgn==6,ngens],2,mean),2)
gres[,9]=round(apply(bdat[bdat$dgn==6,ngens],2,var),2)
gres[,10]=length(bdat$dgn[bdat$dgn==1])-apply(bdat[bdat$dgn==1,ngens],2,nzs)
gres[,11]=round(apply(bdat[bdat$dgn==1,ngens],2,mean),2)
gres[,12]=round(apply(bdat[bdat$dgn==1,ngens],2,var),2)
gres # compare supplementary table S1

# set up various names and lists
yl='importance'; xl='measure'
errvars=list(NULL); varsel=list(NULL); usevars=list(NULL); N=1
# aucs = list to hold areas under the Receiver Operating Characteristic curve
aucs=array(NA,c(N,4),dimnames=list(NULL,c('AUC','se','L99%','U99%')))
prcs=array(NA,c(N,2),dimnames=list(NULL,c('integral','davisgoad')))
aprcs=array(NA,c(N,2),dimnames=list(NULL,c('C','D')))
jprcs=list(NULL); v1vars=list(NULL); vimps=list(NULL)
alluc=array(NA,c(N,2),dimnames=list(NULL,c('C','D')))
ptab=array(NA,c(N,4,2),dimnames=list(NULL,c('TN','FN','FP','TP'),c('C','D')))
errate=array(NA,c(N,3),dimnames=list(NULL,c('all','C','D')))

# load libraries
library(randomForestSRC); library(precrec); library(ROCR); library(cvAUC);
library(PRROC); library(psych)
jr=function(x) return(format(round(100*x,2),justify='right', width=6))
# clear out unused memory
v=0; gc(); gc()

# SET UP TEXT FILE TO SAVE ALL OUTPUT
sink(paste0(mydir,'random forest main output ',mydate(),'.txt'), split=T)
# SET UP pdf FILE TO SAVE PARTIAL PLOTS
w=0; pdf(paste0(mydir,'AUROCs and partial plots ',mydate(),'.pdf'))

# define contrasts of interest
gcons=c('ANHAC v ME/CFS','ANHAC v IBS' ,'ANHAC v Ca')
j='genus'; g=0; k=1
for (h in gcons) { g=g+1

# set random number seed, for reproducibility
set.seed(19321215)

# define new data frame X for each comparison of interest
X=bdat[,c(1,2,7:100)] #  select only age, sex and abundances
if (h=='ANHAC v Ca') { X$ill=as.factor(as.numeric(bdat$dgn%in%c(4)));
X=X[bdat$dgn%in%c(1,4),] }
if (h=='ANHAC v ME/CFS') { X$ill=as.factor(as.numeric(bdat$dgn==4));
X=X[bdat$dgn==4|bdat$dgn==6,] }
if (h=='ANHAC v IBS') { X$ill=as.factor(as.numeric(bdat$dgn==4));
X=X[bdat$dgn==4|bdat$dgn==2,] }
X$ill=as.factor(X$ill); dim(X); print(h); table(X$ill)
# check that X contains the correct diagnostic groups
# iok=rownames(bdat)%in%rownames(X); table(X$ill,bdat$dgn[iok])


w=1; v=v+1; set.seed(jrnd(v)); topvars=NULL

# define all predictors except age, sex and ill
print(paste('+++++++++',h,j,'+++++++++')); gc()
# make case weights to account for imbalances between group sizes
yill = randomForestSRC:::make.wt(X$ill)
```

```
ssize = randomForestSRC:::make.size(X$ill)

# set formulae for random forest models
jf1='ill~age + sex'; jf2='ill~.'

# use preliminary random forest model to select variables for definitive
discrimination model
v1=try(var.select.rfsrc(formula=formula(jf2), data=X, case.wt=yill, ntree=1e4,
nodesize=1, nsplit=10, nodedepth=5,
refit=F, verbose=F, forest=T, save.memory=T, importance='permute',
splitrule='auc',sampsize = ssize, seed=-jrnd(v)), silent=T)
# print the output of the selection stage
print(paste('=========== var selection ==========='))
print(v1$errate); errvars[[h]]=v1$errate
print(v1$varselect[1:25,]); varsel[[h]]=v1$varselect

# save the selected predictors; if none selected, just use age and sex
topvars= v1$topvars; if (identical(v1$topvars,character(0)))
topvars=c('age','sex'); print(topvars)
if ('age'%in%topvars) X=X[,!names(X)=='age.1']
if ('sex'%in%topvars) X=X[,!names(X)=='sex.1']
agen=regexpr('age',names(X))==1; if (table(agen)[2]>1) stop('>1 age')
sexn=regexpr('sex',names(X))==1; if (any(regexpr('sex',names(X))) & table(sexn)
[2]>1) stop('>1 sex')
dvars=names(X)[3:(dim(X)[2]-1)]

# now run the definitive discrimination random forest model
# first, create the formula to use on the selected variable from the preceding
variable selection RF
jf3=paste0('ill~',paste(topvars,collapse='+'))
#next, run the definitive discrimination analysis
v1=try( rfsrc(formula=as.formula(jf3), data=X, case.wt=yill, ntree=2e5,
nodedepth=5, nodesize=1,
nsplit=10, mtry=2, refit=F, verbose=F, forest=T, save.memory=T,
importance='permute', splitrule='auc',sampsize = ssize), silent=T)
print(v1)
# plot(v1) # plot the development of the RF, if wanted
# save the definitive discrimination model
saveRDS(v1,file=paste0('final ',ifelse(h=='ANHAC v ME/CFS','ANHAC v CFS',h),'
',j,' ',mydate(),'.rds'))
nx5=length(topvars)

# extract the variable importance measures from the definitive RF
iill=v1$importance; vimps[[h]]=iill
# print the importances
if (nx5==1) print(iill) else iill=iill[order(-iill[,1]),][1:nx5,]

# save the actual diagnostic group for each person together with their out-of-
bag prediction
jprcs[[h]]=cbind(X$ill,v1$predicted.oob[,2])
# calculate and save error rates for IBS and ME/CFS
errate[w,]=apply(v1$err.rate,2,mean,na.rm=T)

# if wanted, look for possible interactions between predictors in the definitive
RF
#v2=find.interaction.rfsrc(v1, method='vimp', nrep=5)
#saveRDS(v2,file=paste0('final ',ifelse(h=='ANHAC v ME/CFS','ANHAC v CFS',h),'
',j,' interactions ',mydate(),'.rds'))
# and print the interactions
#print(v2); plot(v2); print(v2[abs(v2[,5])>(abs(v2[,3])+abs(v2[,4])/2),])

# calculate and save AUROCs for IBS and ME/CFS
pred1=prediction(predictions=v1$predicted.oob[,2],labels=v1$yvar)
# now calculate confidence intervals for the AUROC
```

```
illci=ci.cvAUC(v1$predicted.oob[,2],v1$yvar,confidence=.95)
aucs[w,1]=illci$cvAUC; aucs[w,2]=illci$se; aucs[w,3:4]=illci$ci
# precision-recall curve, if wanted
# pr1=pr.curve(scores.class0=v1$predicted.oob[,2],
weights.class0=as.numeric(as.character(v1$yvar)), curve=T)
# plot(pr1, main=paste(h,j)); abline(h=table(X$ill)[2]/dim(X)[1], lty=3)
# prcs[[h]][w,1]=pr1[[2]]; prcs[w,2]=pr1[[3]]

# plot AUROCs for IBS and ME/CFS (to file)
if (w==1) { perf1=performance(pred1,'tpr','fpr'); plot(perf1, main=paste(h,j))
abline(0,1,lty=3); legend('bottomright',legend=paste0('AUC =
',round(100*aucs[w,1],1))) }

# set up labels for partial plots of predictors in definitive discrimination RF
clab=list(c('ME/CFS','ANHAC'), c('IBS','ANHAC'),c('Cancer','ANHAC'))
plab=c('p(ME/CFS)','p(IBS)','p(cancer)')

for (i in rownames(iill)) { # loop through the important variables in the
discrimination RF
m = paste(clab[[g]][1],clab[[g]][2],sep=' vs ') # create title for each plot
# create y-axis labels
yl=ifelse (i %in%'sex',i,paste(i,'(%)')) ; if (i=='age') yl='years'
## generate box-and-whisker plot to show overall effect of each predictor
#boxplot(X[,i]~X$ill, varwidth=T, notch=T, col='lightgrey', xlab='clinical
group', xaxt='n', at=c(2,1),
#main=paste0(m,':',i), ylab=yl)
# special case x-axis labels for age and sex
#if (i!='sex') axis(side=1, at=c(2,1), labels=clab[[g]])
#if (i=='sex') axis(side=1, at=c(2,1), labels=c('M','F'))

# generate the partial plot values, but don't plot them
p1=plot.variable(v1, oob=T, xvar.names=i, partial=T, ylab=plab[g],
xlab=gsub('Unknown.','Unclassified ',i), main=m, xaxt=ifelse(i=='sex','n','y'),
smooth.lines=T, show.plots=F)
# special case axis names for age and sex
if (i=='sex') {axis(side=1, at=c(2,1), labels=c('1','0'), col='white');
axis(side=1, at=c(2,1), labels=c('M','F'))}

# standardise partial plots to same y limits
jp1=p1$pData[[1]]; ix=paste(gsub('Unknown.','Unclassified
',i),ifelse(i=='age','','(%)'))
# and plot them
plot(jp1$x.uniq,jp1$yhat,pch=19,col=0,ylim=c(.3,.9), ylab=plab[g], xlab=ix,
main=m)
lines(lowess(jp1$x.uniq,jp1$yhat,1/3),lty=2,lwd=2,col=2)
lines(lowess(jp1$x.uniq,jp1$yhat+1.65*jp1$yhat.se,1/3),lty=3,col=2)
lines(lowess(jp1$x.uniq,jp1$yhat-1.65*jp1$yhat.se,1/3),lty=3,col=2)
points(jp1$x.uniq,jp1$yhat,pch=19)
} # end for i in rownames(iill) =  end loop through important variables

rm(v1); gc(); gc() # remove discrimination RF, to save memory, and tidy up
# output some intermediate data - AUROC, its confidence intervals, error rate
cat(w,date(),round(aucs[w,],3),' error:
',round(errate[w,1],3),round(prcs[w,2],3),'\n')
print(aucs[1,]); if (w>2) print(apply(aucs[-c(1:2),],2,quantile,na.rm=T))
print('------------------------'); print(''); print('')

# end loop over comparisons between ANHAC and other groups
} # end for h


# finally, save the outputs
saveRDS(jprcs, paste0(mydir,'retry final ',j,' jprcs to compute PRCS
',mydate(),' rerun.rds'))
```

```
saveRDS(errvars,paste0(mydir,'retry final ',j,' auc agesex selected variables
',mydate(),' rerun.rds'))
saveRDS(varsel,paste0(mydir,'retry final ',j,' varused and varsel ',mydate(),'
rerun.rds'))
saveRDS(vimps,paste(mydir,'retry final ',j,' variable importances
',mydate(),' .rds'))
# precision-recall curve values, if wanted
# saveRDS(prcs, paste0(mydir,'retry final ',j,' prcs agesex trend for selected
variables ',mydate(),' rerun.rds'))
# close all text and pdf output files
sink(); dev.off(4); dev.off(3); dev.off(2); dev.list()

#############################################################################
########################################################################
#= END OF R CODE TO ANALYSE DATA ======================================#
#= USING RANDOM FOREST, FOR MAIN REPORT ===============================#
#======================================================================#
#======================================================================#
#======================================================================#
#======================================================================#
#======================================================================#
#======================================================================#
#======================================================================#
########################################################################
```
