## Supplementary material for "An approach to finding specific forms of dysbiosis that associate with different disorders": R code for analysis of Cao's dataset

```
# Cao's supplementary table 2, available at:
# https://www.sciencedirect.com/science/article/pii/S0969996123004114?via
%3Dihub#ec0010
# we downloaded the above table and saved it as "Cao_tab2.csv"
# read in Cao Supplementary Table 2
cdat=read.csv(paste0(mydir,'Cao_tab2.csv'),skip=1)

# extract genera from data file
dim(cdat); cdat=cdat[substr(cdat$Microbiota,1,6)=='genus.',]; dim(cdat)
cdat$Microbiota=gsub('[.]','X',cdat$Microbiota)
cdat$Microbiota=gsub('genusX','',cdat$Microbiota)
gstop=regexpr('XidX',cdat$Microbiota)-1
cdat$Microbiota=substr(cdat$Microbiota,1,gstop)
cdat$Microbiota=gsub('X','',cdat$Microbiota)

# rename genera to be compatible with those of main report
# (in order to be able to detect overlap for all available genera
# that can, in fact, correspond between Cao's data and ours)

cdat$Microbiota=gsub("Coprococcus3","Coprococcus.3",cdat$Microbiota)
cdat$Microbiota=gsub("LachnospiraceaeUCG001","Lachnospiraceae.UCG.001",cdat$Micr
obiota)
cdat$Microbiota=gsub("RuminococcaceaeUCG002","Ruminococcaceae.UCG.002",cdat$Micr
obiota)
cdat$Microbiota=gsub(".Eubacteriumeligensgroup","Eubacterium.eligens.group",cdat
$Microbiota)
cdat$Microbiota=gsub("Ruminococcus1","Ruminococcus.group.1",cdat$Microbiota)
cdat$Microbiota=gsub("Ruminococcus2","Ruminococcus.group.2",cdat$Microbiota)
cdat$Microbiota=gsub("Prevotella9","Prevotella.group.9",cdat$Microbiota)
cdat$Microbiota=gsub("Eubacteriumhalliigroup","Eubacterium.hallii.group",cdat$Mi
crobiota)
cdat$Microbiota=gsub("Eubacteriumruminantiumgroup","Eubacterium.ruminantium.grou
p",cdat$Microbiota)
cdat$Microbiota=gsub("Clostridiumsensustricto1","Clostridium.sensu.stricto.1",cd
at$Microbiota)
cdat$Microbiota=gsub("Clostridiuminnocuumgroup","Clostridium.innocuum.group",cda
t$Microbiota)
cdat$Microbiota=gsub("CandidatusSoleaferrea","Candidatus.Soleaferrea",cdat$Micro
biota)
cdat$Microbiota=gsub("ErysipelotrichaceaeUCG003","Erysipelotrichaceae.UCG.003",c
dat$Microbiota)
cdat$Microbiota=gsub("EscherichiaShigella","Escherichia.Shigella",cdat$Microbiot
a)
cdat$Microbiota=gsub("Ruminiclostridium1","Ruminiclostridium.group.1",cdat$Micro
biota)
cdat$Microbiota=gsub("Ruminiclostridium6","Ruminiclostridium.group.6",cdat$Micro
biota)
cdat$Microbiota=gsub("Ruminiclostridium9","Ruminiclostridium.group.9",cdat$Micro
biota)
cdat$Microbiota=gsub("Tyzzerella3","Tyzzerella",cdat$Microbiota)

# extract unique genera names
jum=unique(cdat$Microbiota); table(jum%in%names(bdat))
jum[!jum%in%names(bdat)]; names(bdat)[!names(bdat)%in%jum]

# next, read in our data
# read in data
mydir='/home/jonathan/Desktop/Atlas/'
bdat=read.csv(paste0(mydir,'Dove_Atlas data 200424.csv'))
# check bdat structure
dim(bdat); names(bdat)[1:7]; table(bdat$dgn)
str(bdat); bdat[1:5,1:15]
# id is patient number; age is years; sex is 0=F, 1=M
# dgn is 1=cancer, 2=IBS, 4=ANHAC, 6=CFS
```

```
# ibs is presence of IBS; cfs is presence of ME/CFS
# all patients have ill=1 (yes)
#  id age sex dgn ibs cfs ill Actinomyces Adlercreutzia Agathobacter Akkermansia
Alistipes
#1  1  77   0   2   1   0   1        0.02          0.00         0.95        0.02
3.36
#2  2  81   0   1   0   0   1        0.05          0.04         1.01        2.03
7.51
#3  3  75   1   4   0   0   1        0.00          0.00         0.01        0.58
3.36
#4  4  85   0   4   0   0   1        0.01          0.06         0.88        7.87
4.25
#5  5  50   0   2   1   1   1        0.00          0.07         0.04        9.61
3.53

# change all ages to quinquennia for purposes of publication (true ages
available on reasonable request)
# bdat$age=round(bdat$age/5)

# raretax is the sum of proportions of genera with more than 80% zero
observations
all(apply(bdat[,7:100],1,sum)==100) # TRUE
# pathgen is the sum of proportions of genera that Atlas defines as 'pathogenic'
and is unused here

# take the names of variables that are individual genera
bnames=names(bdat)[7:99]

# now that the names of the genera in both Cao's data and our are available
# we can do the permutation analysis to find out how often there would be
overlap of 2 genera
# (namely, Dialister and Roseburia) in random samples of 7 genera from Cao and 3
from our data
# (because Cao reported that 7 genera had causal effects on cognition and our
random forest
# analyses selected 3 taxa - Dialister, Roseburia and Unknown Bacteria - to
predict ME/CFS)

# set seed for repeatability (111 was cricket umpire "Dickie Bird's"
superstitious number)
# since he thought that it signifies the 3 stumps of a cricket wicket and that
when the batsmen
# next scored, one of the stumps (i.e. the wicket) would fall. So, he used to
hop when the score was 111!
# 100000 permutations

set.seed(111); N=1e5
# set up an array to hold the results of the 100000 permutations
res=rep(0,N)
for (i in 1:N) {
# in each run, select 7 genera from Cao's data-set
csamp=sample(jum,7)
# in each run, select 3 genera from data-set of the main report
jsamp=sample(bnames,3)
# record how many genera selected from Cao's data-set overlap those selected
from our data-set
if (any(jsamp%in%csamp)) res[i]=length(jsamp[jsamp%in%csamp])
}
# finally, report the results
table(res)
# res
#     0     1     2     3
# 87916 11634   449     1
# so p = 0.0045
```
