## Supplementary material for "An approach to finding specific forms of dysbiosis that associate with different disorders": R code for analysis of Gacesa's dataset

```
######################################################################
#= START OF R CODE TO ANALYSE DATA ==================================#
#= FROM SUPPLEMENTARY TABLE S3B OF GACESA 2022 ======================#
#====================================================================#
#====================================================================#
#====================================================================#
#====================================================================#
#====================================================================#
#====================================================================#
#====================================================================#
######################################################################

#  A FEW USEFUL FUNCTIONS
ac=function(x) return(as.character(x))
an=function(x) return(as.numeric(x))
anc=function(x) return(an(ac(x)))

# s3b IS TABLE S3B IN SUPPLEMENTARY INFORMATION OF GACESA (2022), available at:
# https://static-content.springer.com/esm/art%3A10.1038%2Fs41586-022-04567-7/
MediaObjects/41586_2022_4567_MOESM4_ESM.xlsx
# we downloaded this an converted it to a comma-separated values (csv) file -
"Gacesa_S3b.csv'
# this file is 14.8MB and is readily available, as above; therefore we do not
include it, here

mydir='/home/jonathan/Desktop/Atlas/' # set working directory
s3b=read.table(paste0(mydir,'Gacesa_S3b.csv'),sep=',',header=T)

# eliminate multi-category phenotypes
s3b$effect_size=anc(gsub('0:','',s3b$effect_size))

# check effect sizes
quantile(s3b$effect_size,na.rm=T)
dim(s3b); names(s3b); s3b[101:110,]

# find effect sizes for genera
gstart=regexpr('.g__',ac(s3b$taxon)); table(gstart)
gstop=regexpr('.s__',ac(s3b$taxon)); table(gstop)
glen=nchar(ac(s3b$taxon))
gok=gstart>0 & gstop<0 & gstart<glen # gstop<0 -> select overall genera, not
individual species
s3b$genus=NA; s3b$genus[gok]=substr(ac(s3b$taxon[gok]),gstart[gok]+4,glen[gok])
# Gacesa reports effect sizes for 59 genera
length(table(s3b$genus)) # 59

# find effect sizes for species
gstart=regexpr('.s__',ac(s3b$taxon)); glen=nchar(ac(s3b$taxon))
gok=gstart>0 & gstart<glen # gstop<0 -> select individual species
s3b$species=NA; s3b$species[gok]=substr(ac(s3b$taxon[gok]),gstart[gok]
+4,glen[gok])
# Gacesa reports effect sizes for 136 species in the 59 genera
length(table(s3b$species)) # 136

# we want to compare the effect sizes for genera that our RF analyses
# identify as important for discriminating cancer, CFS, IBS or CFS+IBS
# with effect sizes for genera that our RF analyses do not identify as important

# here in Gacesa's data:-
# anhac is the equivalent of the ANHAC group in our study
# cancer is the equivalent of the cancer group
# ibs is the equivalent of pure IBS - which does not exist in our study

# retain only effect sizes for genera
s3j=s3b[!is.na(s3b$genus),]; dim(s3j)
```

```
# find names of all phenotypes that are medical diseases/conditions
alldis=regexpr('MED.DISEASES',ac(s3j$phenotype))>0; table(alldis)
# retain only effect sizes of genera on diseases/conditions
dim(s3j); s3j=s3j[alldis,]; dim(s3j)

# these are the diseases/conditions that we selected to study here
okudis=c(1,3,12,20:28,33,42,44,47,48,49,51:53,57:68) #
unique(s3j$phenotype)[okudis]
# [1] "MED.DISEASES.Blood.Anemia"
# [2] "MED.DISEASES.Cancer.AnyNonBasal"
# [3] "MED.DISEASES.Cardiovascular.Atherosclerosis"
# [4] "MED.DISEASES.Cardiovascular.Hypertension"
# [5] "MED.DISEASES.Endocrine.Autoimmune.DiabetesT1"
# [6] "MED.DISEASES.Endocrine.DiabetesT2"
# [7] "MED.DISEASES.Gastrointestinal.Autoimmune.Celiac"
# [8] "MED.DISEASES.Gastrointestinal.Autoimmune.IBD.CD"
# [9] "MED.DISEASES.Gastrointestinal.Autoimmune.IBD.UC"
#[10] "MED.DISEASES.Gastrointestinal.Stomach.Ulcer"
#[11] "MED.DISEASES.Hepatologic.Gallstones"
#[12] "MED.DISEASES.Hepatologic.Hepatitis"
#[13] "MED.DISEASES.Mental.Any"
#[14] "MED.DISEASES.Neurological.Autoimmune.Multiple.Sclerosis"
#[15] "MED.DISEASES.Neurological.Epilepsy"
#[16] "MED.DISEASES.Neurological.Mental.Fatigue.Chronic"
#[17] "MED.DISEASES.Neurological.Mental.Fibromyalgia"
#[18] "MED.DISEASES.Neurological.Migraine"
#[19] "MED.DISEASES.Neurological.Stroke"
#[20] "MED.DISEASES.None.No.Diseases"
#[21] "MED.DISEASES.Other.Autoimmune.Rheumatoid.Artritis"
#[22] "MED.DISEASES.Other.Fractures.Hip"
#[23] "MED.DISEASES.Other.Fractures.Other"
#[24] "MED.DISEASES.Other.Kidney.Stones"
#[25] "MED.DISEASES.Other.Osteoarthritis"
#[26] "MED.DISEASES.Other.Osteoporosis"
#[27] "MED.DISEASES.Pulmonary.Autoimmune.Asthma"
#[28] "MED.DISEASES.Pulmonary.COPD"
#[29] "MED.DISEASES.Pulmonary.Pulmonary.Embolism"
#[30] "MED.DISEASES.Skin.Autoimmune.Atopic.dermatitis"
#[31] "MED.DISEASES.Skin.Autoimmune.Psoriasis"
#[32] "MED.DISEASES.Skin.Autoimmune.Severe.acne"
#[33] "MED.DISEASES.Gastrointestinal.Rome3_IBS.Any"

# use only the above selected diseases/conditions
s3j=s3j[as.factor(s3j$phenotype)%in%unique(s3j$phenotype)[okudis],]; dim(s3j)
table(s3j$phenotype) # effect sizes of 59 genera for each disease/condition

# extract size of group with each disease/condition
jtn=unlist(lapply(s3j[,5],strsplit,':')); ntn=1:length(jtn)
s3j[,5]=jtn[ntn%%2==0]; names(s3j)[5]='N'; table(s3j$N)
s3j$N=anc(s3j$N)

# define groups to analyse
cancer=regexpr('Cancer',ac(s3j$phenotype))>0; table(cancer)
ibs=regexpr('IBS',ac(s3j$phenotype))>0; table(ibs)
cfs=regexpr('atigue',ac(s3j$phenotype))>0; table(cfs)
fma=regexpr('ibromyalgia',ac(s3j$phenotype))>0; table(fma)
hc=regexpr('None.No.Diseases',ac(s3j$phenotype))>0; table(hc)
ms=regexpr('Multiple.Sclerosis',ac(s3j$phenotype))>0; table(ms)
table(cfs,ibs); table(cfs,fma); table(ibs,fma)

# there are, apparently, NO dual-diagnosis cases in the data-set
# this is probably an artefact of using regression-based effect sizes
# (because regression accounts for every other phenotype when defining
```

```
# effect of each taxon on each phenotype
# but Figure 4b also indicates little comorbidity (GP diagnoses?)

# define the anhac group
anhac=!cancer & !ibs & !cfs & !hc & !fma

# check the anhac group (we selected a genus with a short name, for convenience)
s3j[anhac & s3j$genus=='Rothia',c(1,5)]

# COMPUTE EFFECT SIZES OF EACH SPECIFIC DISORDER, *RELATIVE TO ANHAC*
# labels of individual genera in the data
ng=ac(unique(s3j$genus)); ng=ng[order(ng)]; nglen=length(ng); ng
#define specific disorders of interest
specdis=c('HC','cancer','IBS','CFS','FMA','MS','ANHAC')
# set up an array to hold the new data
res=array(NA,c(nglen,6,7),dimnames=list(ng,c('t','p','R2','var','wtx','N'),specd
is))

# for each genus, in turn
j=0; for (i in ng) { sok=s3j$genus==i; j=j+1
# find the effect sizes for all disorders comprising the anhac group
anhac_es=s3j$effect_size[sok & anhac]
# get the variances, standard deviations and means of the anhac group's effect
sizes
vanhac=var(anhac_es,na.rm=T); anhacsd=sqrt(vanhac); ahx=mean(anhac_es)
# also get the Pvalues and group sizes of each anhac group
anhac_p=s3j$Pvalue[sok & anhac]
anhac_n=s3j$N[sok & anhac]
# print the genus, its group size, mean effect size and sd for the anhac group
cat(i,anhac_n,round(ahx,3),round(anhacsd,3),'\n')

# now repeat the above for the cfs, ibs, cancer, fibromyalgia and multiple
sclerosis groups
cfsx=s3j$effect_size[sok & cfs]
hcx=s3j$effect_size[sok & hc]
ibx=s3j$effect_size[sok & ibs]; vibs=var(ibx)
cax=s3j$effect_size[sok & cancer]; vca=var(cax)
fmx=s3j$effect_size[sok & fma]
msx=s3j$effect_size[sok & ms]
res[j,1,1]=hcx
res[j,2,1]=round(1-pt(abs(res[j,1,1]),length(anhac_es)-2),4)
res[j,1,2]=mean(cax)
res[j,2,2]=round(1-pt(abs(res[j,1,2]),length(anhac_es)-2),4)
res[j,1,3]=mean(ibx)
res[j,2,3]=round(1-pt(abs(res[j,1,3]),length(anhac_es)-2),4)
res[j,1,4]=cfsx
res[j,2,4]=round(1-pt(abs(res[j,1,4]),length(anhac_es)-2),4)
res[j,1,5]=fmx
res[j,2,5]=round(1-pt(abs(res[j,1,5]),length(anhac_es)-2),4)
res[j,1,6]=msx
res[j,2,6]=round(1-pt(abs(res[j,1,6]),length(anhac_es)-2),4)
res[j,1,7]=ahx/anhacsd
res[j,2,7]=round(1-pt(abs(res[j,1,7]),length(anhac_es)-2),4)
res[j,3,1]=s3j$R2[sok&hc]
res[j,3,2]=s3j$R2[sok&cancer]
res[j,3,3]=s3j$R2[sok&ibs]
res[j,3,4]=s3j$R2[sok&cfs]
res[j,3,5]=s3j$R2[sok&fma]
res[j,3,6]=s3j$R2[sok&ms]
res[j,3,7]=mean(s3j$R2[sok&anhac])
res[j,4,1]=1 #s3j$R2[sok&hc]
res[j,4,2]=1 #s3j$R2[sok&cancer]
res[j,4,3]=1 #s3j$R2[sok&ibs]
res[j,4,4]=1 #s3j$R2[sok&cfs]
```

```
res[j,4,5]=1 #s3j$R2[sok&fma]
res[j,4,6]=1 #s3j$R2[sok&ms]
res[j,4,7]=vanhac
res[j,5,1]=1
res[j,5,2]=1
res[j,5,3]=1
res[j,5,4]=1
res[j,5,5]=1
res[j,5,6]=1
res[j,5,7]=weighted.mean(anhac_es,log(1/anhac_p))
#round(ks.test(anhac_es,res[j,2,6])$p.value,4)
res[j,6,7]=weighted.mean(anhac_es,anhac_n) #round(ks.test(anhac_es,res[j,2,6])
$p.value,4)
} # end for i in ng
res[,,'ANHAC']

# check that the new data array - 'res' - looks sensible
round(cbind(res[,6,'ANHAC'],res[,1,'IBS'],res[,1,'CFS'],res[,1,'cancer']),2)

# Gacesa abbreviated some species names
# S. salivarius = Streptococcus salivarius
# C. asparagiforme = Clostridium asparagiforme
# R. gnavus = Ruminococcus gnavus
# E. rectale = Eubacterium
# F. prausnitzii = Faecalibacterium
# D. piger = Desulfovibrio
# A. senegalensis = Alistipes
# B. longum = Bifidobacterium

# FROM GACESA FIGURE 4A
# genera that associate negatively with healthy controls
hcneg=c('Clostridium','Anaerotruncus','Ruminococcus','Ruminococcaceae',
'Eggerthella','Flavonifractor','Oscillibacter','Pseudoflavonifractor',
'Holdemania','Streptococcus','Veillonella','Bifidobacterium','Lachnospiraceae')
# genera that associate positively with healthy controls
hcpos=c('Faecalibacterium','Eubacterium','Subdoligranulum','Dorea',
'Bifidobacterium','Butyrovibrio','Alistipes','Desulfovibrio',
'Prevotella','Paraprevotella','Mitsuokella','Barnesiella','Bacteroidiales')
pos_es=res[dimnames(res)[[1]]%in%hcpos,1,'HC']
neg_es=res[dimnames(res)[[1]]%in%hcneg,1,'HC']
t.test(pos_es,neg_es); wilcox.test(pos_es,neg_es)
xl='clinical grouping'; yl='effect size'
boxplot(neg_es,pos_es,notch=T,varwidth=T,xaxt='n',xlab=xl,ylab=yl)
axis(side=1, at=c(1,2), labels=c('negative','positive'))
legend('topleft',legend=c('Clostridium','Anaerotruncus','Ruminococcus',
'Ruminococcaceae','Eggerthella','Flavonifractor','etc.'),cex=0.75)
legend('bottomright',legend=c('Faecalibacterium','Eubacterium','Subdoligranulum'
,
'Dorea','Bifidobacterium','etc.'),cex=0.75)
xl="effect sizes of genera for HC (cf Gacesa's Fig 4a)"
# compare with Gacesa Figure 4a

# now, we need to match the genera in our data-set with those in Gacesa's
dovegen=c("Actinomyces","Adlercreutzia","Agathobacter",
"Akkermansia","Alistipes","Anaerostipes", "Anaerotruncus","Bacteroides",
"Barnesiella",  "Bifidobacterium","Bilophila","Blautia","Butyricicoccus",
"Butyricimonas","Candidatus.Soleaferrea","Chloroplast.group.2",
"Clostridium.sensu.stricto.1","Collinsella","Coprobacter","Coprococcus.3",
"Desulfovibrio","Dialister","Dorea","DTU089","Eggerthella","Eisenbergiella",
"Erysipelatoclostridium","Erysipelotrichaceae.UCG.003","Escherichia.Shigella",
"Eubacterium.eligens.group","Eubacterium.hallii.group","Faecalibacterium",
"Flavonifractor","Gordonibacter","Haemophilus","Holdemanella","Holdemania",
"Hungatella","Intestinibacter","Intestinimonas","Lachnoclostridium","Lachnospira
",
```

```
"Lachnospiraceae.UCG.001","Lactobacillus","Methanobrevibacter","Mitsuokella",
"Mogibacterium","Muribaculum","Odoribacter","Olsenella","Oscillibacter","Oxaloba
cter",
"Papillibacter","Parabacteroides","Parasutterella","Phascolarctobacterium","Phoc
ea",
"Prevotella.group.9","Romboutsia","Roseburia","Rothia","Ruminiclostridium",
"Ruminiclostridium.group.1","Ruminiclostridium.group.6","Ruminiclostridium.group
.9",
"Ruminococcaceae.UCG.002","Ruminococcus.group.1","Ruminococcus.group.2",
"Ruminococcus.Lachnospiraceae","Senegalimassilia","Slackia","Streptococcus",
"Sutterella","Terrisporobacter","Turicibacter","Tyzzerella","UBA1819",
"Unknown.Bacteria","Unknown.Bacteroidales","Unknown.Burkholderiaceae",
"Unknown.Christensenellaceae","Unknown.Clostridiales","Unknown.Coriobacteriales"
,
"Unknown.Enterobacteriaceae","Unknown.Erysipelotrichaceae","Unknown.Firmicutes",
"Unknown.Lachnospiraceae","Unknown.Ruminococcaceae","Veillonella","Victivallis")
# which of Gacesa's genera do not occur in our complete list of genera
# (complete = before selection of genera to discriminate ANHAC group from single
disorders)
na.omit(unique(s3j$genus)[!s3j$genus%in%dovegen])
# which of our genera do not occur in Gacesa's list of genera
na.omit(unique(dovegen)[!dovegen%in%s3j$genus])
unique(s3j$genus)[order(unique(s3j$genus))]
s3gen=c("Acidaminococcus","Actinomyces",  "Adlercreutzia","Akkermansia",
"Alistipes","Anaerostipes", "Anaerotruncus","Bacteroidales_noname",
"Bacteroides","Barnesiella","Bifidobacterium","Bilophila","Blautia",
"Burkholderiales_noname","Butyrivibrio", "Catenibacterium",
"Clostridiales_noname","Clostridium","Collinsella","Coprobacter",
"Coprococcus","Desulfovibrio","Dialister","Dorea","Eggerthella",
"Erysipelotrichaceae_noname","Escherichia","Eubacterium",
"Faecalibacterium","Flavonifractor","Gordonibacter","Haemophilus",
"Holdemania","Klebsiella","Lachnospiraceae_noname","Lactobacillus",
"Megamonas","Megasphaera","Methanobrevibacter","Mitsuokella",
"Odoribacter","Oscillibacter","Oxalobacter","Parabacteroides",
"Paraprevotella","Parasutterella","Peptostreptococcaceae_noname",
"Phascolarctobacterium","Prevotella","Pseudoflavonifractor","Roseburia",
"Rothia","Ruminococcaceae_noname","Ruminococcus","Streptococcus",
"Subdoligranulum","Sutterella","Sutterellaceae_unclassified","Veillonella")

# rename some genera in our study to match those in Gacesa's
# (we are not specialists in taxonomy of microbiota -
# so, we apologise if there are errors)
dovegen[dovegen=='Unknown.Clostridiales']='Clostridiales_noname'
dovegen[dovegen=='Unknown.Ruminococcaceae']='Ruminococcaceae_noname'
dovegen[dovegen=='Unknown.Bacteroidales']='Bacteroidales_noname'
dovegen[dovegen=='Unknown.Lachnospiraceae']='Lachnospiraceae_noname'
dovegen[dovegen=="Eubacterium.hallii.group"]="Eubacterium"
dovegen[dovegen=="Eubacterium.eligens.group"]="Eubacterium"
dovegen[dovegen=="Clostridium.senus.stricto.1"]="Clostridium"
dovegen[dovegen=="Escherichia.Shigella"]="Escherichia"
dovegen[dovegen=="Prevotella.group.9"]="Prevotella"
dovegen[dovegen=="Erysipelotrichaceae.UCG.003"]="Erysipelotrichaceae_noname"
dovegen[dovegen=="Ruminococcus.group.1"]="Ruminococcus"
dovegen[dovegen=="Ruminococcus.group.2"]="Ruminococcus"
dovegen[dovegen=="Ruminococcus.Lachnospiraceae"]="Ruminococcaceae_noname"
dovegen[dovegen=="Ruminococcaceae.UCG.002"]="Ruminococcaceae_noname"

dovegen[dovegen%in%s3gen] # 49 of our genera occur also in Gacesa's genera
dovegen[!dovegen%in%s3gen] # 41 of our genera do not occur in Gacesa's genera
s3gen[s3gen%in%dovegen] # 44 of Gacesa's genera occur in our genera (why not
49?)
s3gen[!s3gen%in%dovegen] # 14 of Gacesa's genera do not occur in our genera

okgen=unique(dovegen[dovegen%in%s3gen]); okgen # 45 genera in common
```

```
# trim down the working data-set to include only those 45 genera
# and check the structure of the working data-set
res=res[okgen,,]; str(res)

# load libraries with robust regression routines
library(MASS); library(robustbase)

# plot variance of effect sizes (y-axis) against their mean (x-axis)
xl='mean(ANHAC effect sizes)'; yl='var(ANHAC effect sizes)'
plot(res[,6,'ANHAC'],res[,4,'ANHAC'],ylab=yl,xlab=xl); abline(v=0,lty=3)
rqud=lmrob(res[,4,'ANHAC']~res[,6,'ANHAC']+I(res[,6,'ANHAC']^2))
summary(rqud); curve(coef(rqud)[1]+coef(rqud)[2]*x+coef(rqud)[3]*x^2,add=T)
# so, for some reason, the mean and variance of the ANHAC effect sizes covary
# it often happens that means and variances covary in biological data
# should we take account of this covariance? - and if we should, then how?
# 2 possibilities - (1) divide mean by variance; (2) weight mean by variance

# NOW CHECK IF THERE IS A NEGATIVE RELATIONSHIP BETWEEN EFFECT SIZES OF GENERA
# ON ANHAC AND HC GROUPS (cf GACESA'S FIGURE 4)
rc0=lmrob(scale(res[,1,'ANHAC'])~scale(res[,1,'HC']))  # QUOTED
round(summary(rc0)$coef,3); summary(rc0)$sigma; summary(rc0)$adj.r.squared
rc1=lmrob(scale(res[,6,'ANHAC'])~res[,1,'HC'])
round(summary(rc1)$coef,3); summary(rc1)$sigma; summary(rc1)$adj.r.squared
rc0a=lmrob(scale(res[,1,'ANHAC'])~res[,1,'HC'],weights=1/res[,4,'ANHAC'])
round(summary(rc0a)$coef,3); summary(rc0a)$sigma; summary(rc0a)$adj.r.squared
rc1a=lmrob(scale(res[,6,'ANHAC'])~res[,1,'HC'],weights=1/res[,4,'ANHAC'])
round(summary(rc1a)$coef,3); summary(rc1a)$sigma; summary(rc1a)$adj.r.squared
# empirically, the raw mean effect sizes explain more variance of genera
# than the weighted-mean effect sizes, in regression of ANHAC on HC values
# therefore, use raw mean effect sizes in further analyses
# plot the raw mean effect sizes of ANHAC vs HC - Figure S1 in Supplementary
Information
xl='effect sizes in Healthy controls'; yl='effect sizes in ANHAC grouping'
plot(scale(res[,6,'ANHAC'])~scale(res[,1,'HC']),xlab=xl,ylab=yl)
abline(lmrob(scale(res[,6,'ANHAC'])~scale(res[,1,'HC'])))


############### CANCER ###################
# first, how far does Gacesa's cancer grouping resemble their anhac grouping
ms0a=lmrob(scale(res[,1,'cancer'])~scale(res[,6,'ANHAC'])) # QUOTED -and see
Figure S3, below
round(summary(ms0a)$coef,3); summary(ms0a)$sigma; summary(ms0a)$adj.r.squared
# adding the effect sizes of genera for HC group does not alter the above
regression
ms0b=lmrob(scale(res[,1,'cancer'])~scale(res[,6,'ANHAC'])+scale(res[,1,'HC']))
round(summary(ms0b)$coef,3); summary(ms0b)$sigma; summary(ms0b)$adj.r.squared
anova(ms0a,ms0b)

# the deviations from the above relationship (from Gacesa's data)
# may indicate genera that associate specifically with cancer
# So, analyse those deviations according to whether random forest selected those
genera
# to discriminate the ANHAC and cancer groupings in *our* data.
cagen=c("Escherichia.Shigella","Ruminococcaceae_noname",
"Collinsella","Bifidobacterium","Unknown.Firmicutes",
"UBA1819","Bilophila","Ruminococcus","Coriobacteriales_noname",
"Enterobacteriaceae_noname","Lachnoclostridium",
"Intestinibacter","Romboutsia","Haemophilus","Intestinimonas",
"Ruminiclostridium.group.9","Chloroplast.group.2",
"Olsenella","Coprobacter","Parasutterella","Lachnospira",
"Dialister","Streptococcus","Erysipelotrichaceae",
"Faecalibacterium","Flavonifractor","Holdemania",
"Ruminiclostridium","Dorea","Victivallis","
Eubacterium","Sutterella","Parabacteroides")
```

```
caok=as.numeric(dimnames(res)[[1]]%in%cagen); table(caok) # 2

# plot the dependence of Gacesa's effect sizes in cancer on those in their anhac
group
scr_anhac=scale(res[,6,'ANHAC']); scr_ca=scale(res[,1,'cancer'])
xl='genus-wise mean effect sizes in ANHAC grouping'
yl='effect sizes in cancer grouping'
# the deviations from the above relationship (from Gacesa's data)
# may indicate genera that associate specifically with cancer.
# So, highlight those genera in the figure
plot(scale(res[,6,'ANHAC']),scale(res[,1,'cancer']), xlab=xl, ylab=yl) # Figure
S3
abline(coef(ms0a)[1:2])
for (i in cagen[cagen%in%dimnames(res)[[1]]])
points(scr_anhac[i,1],scr_ca[i,1],pch=19)
resca=resid(ms0a)[caok==1]; resanhac1=resid(ms0a)[caok==0]

# plot the densities of the residuals, according to whether RF selected them in
our data
plot(density(resid(ms0a)[caok==0]),xlim=c(-3.5,3), main='', xlab='unexplained
effect')
lines(density(resid(ms0a)[caok==1]),lty=2);
legend('topleft',lty=c(0,1,2),legend=c('RF','No','Yes'))
# it appears that the effect sizes - in Gacesa's data - of genera that RF
selected to discriminate
# our ANHAC and cancer groups have greater variance than the genera that RF did
not select

# Finally, test if the deviations from the association between anhac and cancer
groupings
# in Gacesa's data are greater for genera that our RF analyses selected to
discriminate
# ANHAC from cancer than for those that our RF analyses did not select
# specifically, do the variances of the residuals of the selected and unselected
genera differ?
jF=var(resid(ms0a)[caok==1])/var(resid(ms0a)[caok==0]);
wilcox.test(abs(resid(ms0a))~caok)
jt=table(caok); jF; jt; 1-pf(jF,jt[2]-1,jt[1]-1) # F=2.13, df=15/28, p=0.041 #
QUOTED
# GENERA THAT ASSOCIATE PARTICULARLY WITH CANCER, OVER-AND-ABOVE THEIR
ASSOCIATION WITH ANHAC
resid(ms0a)[abs(resid(ms0a))>1]
#            Desulfovibrio                 Dialister
Erysipelotrichaceae_noname
#                -1.297263                 -1.365982
1.402215
#              Haemophilus            Parasutterella
Rothia
#                -1.854824                  1.197394                         -
1.478908
#    Ruminococcaceae_noname             Streptococcus
#                 1.386178                 -1.043001

####################### IBS ########################
ibsgen=unique(c("Dialister","Ruminococcaceae","Unknown.Bacteria",
"Bilophila","Coprococcus","Victivallis",
"Mogibacterium","Romboutsia","Ruminococcus",
"Ruminiclostridium","Bifidobacterium","Parabacteroides",
"Roseburia","Erysipelotrichaceae","Holdemania",
"Agathobacter","Parasutterella","Intestinibacter",
"Prevotella","Lachnoclostridium","Dorea",
"Candidatus.Soleaferrea","Bacteroides","Ruminococcus",
"Olsenella","UBA1819","Eubacterium","Flavonifractor"))
length(ibsgen) # 26
```

```
ibsok=as.numeric(dimnames(res)[[1]]%in%ibsgen); table(ibsok) # 14

ms0a=lmrob(scale(res[,1,'IBS'])~scale(res[,6,'ANHAC']))
round(summary(ms0a)$coef,3); summary(ms0a)$sigma; summary(ms0a)$adj.r.squared
ms0b=lmrob(scale(res[,1,'IBS'])~scale(res[,6,'ANHAC'])+scale(res[,1,'HC']))
round(summary(ms0a)$coef,3); summary(ms0a)$sigma; summary(ms0a)$adj.r.squared
anova(ms0a,ms0b)

scr_anhac=scale(res[,6,'ANHAC']); scr_ibs=scale(res[,1,'IBS'])
xl='genus-wise mean effect sizes in ANHAC grouping'
yl='effect sizes in IBS group'
plot(scale(res[,6,'ANHAC']),scale(res[,1,'IBS']), xlab=xl, ylab=yl)
abline(coef(ms0a)[1:2])
for (i in cagen[cagen%in%dimnames(res)[[1]]])
points(scr_anhac[i,1],scr_ibs[i,1],pch=19)

resibs=resid(ms0a)[ibsok==1]; resanhac2=resid(ms0a)[ibsok==0]
jF=var(resid(ms0a)[ibsok==1])/var(resid(ms0a)[ibsok==0])
jt=table(ibsok); jF; jt; 1-pf(jF,jt[2]-1,jt[1]-1) # F=1.34, df=12/31, p=0.25
resid(ms0a)[abs(resid(ms0a))>1]; wilcox.test(abs(resid(ms0a))~ibsok)
#           Holdemania
#              1.00687

# finally, combine the data for IBS and cancer groupings
# make a vector to hold the unselected (anhac) effect sizes
resanhac=rep(NA,length(unique(c(names(resanhac1),names(resanhac2)))))
names(resanhac)=unique(c(names(resanhac1),names(resanhac2)))
# and populate the vector with the mean unselected (anhac) effect sizes
for (i in names(resanhac)) {
if (i%in%names(resanhac1)&i%in%names(resanhac2)) {
resanhac[i]=mean(c(resanhac1[i],resanhac2[i])) }
if (i%in%names(resanhac1)&!i%in%names(resanhac2)) {
resanhac[i]=resanhac1[i] }
if (!i%in%names(resanhac1)&i%in%names(resanhac2)) {
resanhac[i]=resanhac2[i] }}

# make a vector to hold the selected (cancer or IBS) effect sizes
residis=rep(NA,length(unique(c(names(resibs),names(resca)))))
names(residis)=unique(c(names(resibs),names(resca)))
# and populate the vector with the mean selected (cancer or IBS) effect sizes
for (i in names(residis)) {
if (i%in%names(resibs)&i%in%names(resca)) {
residis[i]=mean(c(resibs[i],resca[i])) }
if (i%in%names(resibs)&!i%in%names(resca)) {
residis[i]=resibs[i] }
if (!i%in%names(resibs)&i%in%names(resca)) {
residis[i]=resca[i] }}


# table(names(residis)%in%names(resanhac))
# FALSE  TRUE
#     9    11
var(resanhac[!names(resanhac)%in%names(residis)])
# [1] 0.1361223
var(resanhac[names(resanhac)%in%names(residis)])
# [1] 0.134783

# variance ratio of all residuals of effect sizes in Gacesa's data
# according to whether they were selected to discriminate individual disorders
in our study
var(residis)/var(resanhac); length(residis)-1; length(resanhac)-1
1-pf(var(residis)/var(resanhac),length(residis)-1,length(resanhac)-1)

# exclude residuals that were selected to discriminate both cancer and IBS -
```

```
QUOTED
aok=!names(resanhac)%in%names(residis)
var(residis)/var(resanhac[aok]); length(residis)-1; length(resanhac[aok])-1
1-pf(var(residis)/var(resanhac[aok]),length(residis)-1,length(resanhac[aok])-1)
# F = 3.133, 19/24df, p=0.0046
# exclude residuals that were selected to discriminate only cancer or only IBS
(i.e. not both)
var(residis)/var(resanhac[!aok]); length(residis)-1; length(resanhac[!aok])-1
1-pf(var(residis)/var(resanhac[!aok]),length(residis)-1,length(resanhac[!aok])-
1)
# F = 3.164, 19/10df, p=0.033

plot(density(residis, adjust=1.5),ylim=c(0,1.5),main='',xlab='standardised
residual',lty=2)
lines(density(resanhac[aok], adjust=1.5),lty=1)
legend('topleft',lty=c(1,2),legend=c('not selected','selected'))

###################### CFS #######################
cfsgen=c("Roseburia","Dialister")
cfsok=as.numeric(dimnames(res)[[1]]%in%cfsgen); table(cfsok) # 2

ms0a=lmrob(scale(res[,1,'CFS'])~scale(res[,6,'ANHAC'])+scale(res[,1,'CFS']))
round(summary(ms0a)$coef,3); summary(ms0a)$sigma; summary(ms0a)$adj.r.squared
scr_anhac=scale(res[,6,'ANHAC']); scr_cfs=scale(res[,1,'CFS'])
plot(scale(res[,6,'ANHAC']),scale(res[,1,'CFS'])); abline(coef(ms0a)[1:2])
for (i in cfsgen[cfsgen%in%dimnames(res)[[1]]])
points(scr_anhac[i,1],scr_cfs[i,1],pch=19)
rescfs=resid(ms0a)[cfsok==1]; resanhac3=resid(ms0a)[cfsok==0]

jF=var(resid(ms0a)[cfsok==1])/var(resid(ms0a)[cfsok==0])
jt=table(cfsok); jF; jt; 1-pf(jF,jt[2]-1,jt[1]-1) # F=4.02, 1/42df, p=0.0515
# not quoted, but consistent with the overall concept that departures from the
shared
# signature of dysbiosis in unrelated diseases may reflect genera that
contribute
# causally to individual disorders
resid(ms0a)[abs(resid(ms0a))>1]
# Dialister is only one of 7 genera that do not conform to the shared signature
of disease
#          Barnesiella             Dialister           Escherichia
Oxalobacter
#            -1.660710             -2.201975              1.016102
1.180069
#       Parasutterella Phascolarctobacterium            Prevotella
#            -1.642325              2.786318              2.864754


# code to generate figures of decision trees in supplementary pdf
library(partykit)
bdat=read.csv(paste0(mydir,'Dove_Atlas data 130324.csv')); bdat=bdat[,-1]
b1=bdat[bdat$dgn%in%c(4,6),]
pdf(paste0(mydir,'decision tree examples 040424.pdf'))
for (i in c(18,21)) {
set.seed(i)
b1x=sample(b1$id[b1$dgn==6],21)
b1x=b1[!b1$id%in%b1x,]; cx=ctree_control(alpha=.2, testtype='MonteCarlo')
b1x$fdgn=as.factor(as.numeric(b1x$dgn==6))
c1=ctree(fdgn~Dialister+Roseburia+Unknown.Bacteria, data=b1x, control=cx)
plot(c1)}
dev.off(3); dev.off(2); dev.list()

######################################################################
#=END OF R CODE TO ANALYSE DATA =====================================#
#=FROM SUPPLEMENTARY TABLE S3B OF GACESA 2022========================#
```

```
#======================================================================#
#======================================================================#
#======================================================================#
#======================================================================#
#======================================================================#
#======================================================================#
#======================================================================#
########################################################################
```
