## Supplementary material for "An approach to finding specific forms of dysbiosis that associate with different disorders": R code for heteroscedasticity analysis

######################################################################
#= START OF R CODE TO CHECK HETEROSCEDASTICITY ======================#
#====================================================================#
#====================================================================#
#====================================================================#
#====================================================================#
#====================================================================#
#====================================================================#
#====================================================================#
#====================================================================#
######################################################################

# read in data
mydir='/home/jonathan/Desktop/Atlas/'
bdat=read.csv(paste0(mydir,'Dove_Atlas data 040424.csv')); bdat=bdat[,-1]
# check bdat structure
dim(bdat); names(bdat)[1:7]; table(bdat$dgn)
str(bdat); bdat[1:5,1:15]

# compute mean abundance of each genus in each diagnostic grouping
x60=apply(bdat[bdat$dgn==6,-c(1:7)],2,mean) # ME/CFS
x10=apply(bdat[bdat$dgn==1,-c(1:7)],2,mean) # cancer
x20=apply(bdat[bdat$dgn==2,-c(1:7)],2,mean) # IBS
x40=apply(bdat[bdat$dgn==4,-c(1:7)],2,mean) # ANHAC
round(cbind(x40,x20,x60,x10),2) # compare Supplementary Table S1

# now test if variances of mean abundances are greater in ANHAC grouping than in single disorders
vF=var(x40)/var(x10); vF; 1-pf(vF,length(x40)-1,length(x10)-1) # ANHAC vs cancer
vF=var(x40)/var(x20); vF; 1-pf(vF,length(x40)-1,length(x20)-1) # ANHAC vs IBS
vF=var(x40)/var(x60); vF; 1-pf(vF,length(x40)-1,length(x60)-1) # ANHAC vs ME/CFS
# all p-values > 0.3 

# plot this (even though nothing is 'significant')
xl='log(mean of individual genera)'; yl=c(0,.2)
plot(density(log(x40)),main='',xlab=xl,ylim=yl)
lines(density(log(x10)),lty=2); lines(density(log(x20)),lty=3)
lines(density(log(x60)),lty=4); lines(density((log(x60)+log(x10)+log(x20))/3),col=2)
xl=c('ANHAC','cancer','IBS','ME/CFS','Ca+IBS+ME')
legend('topright', lty=c(1:4,1), col=c(rep(1,4),2), legend=xl)


# now test if proportions of zero observations in each genus differ between diagnostic groupings
# function to count number of zero observations
nz=function(x) return(length(x[x==0]))
# ngens is names of all genera used in the analyses
ngens=names(bdat)[8:101]; ngens
# number of genera with no zero observations in ANHAC group
t0=table(apply(bdat[bdat$dgn==4,ngens],2,nz)==0); t0 #21 - i.e. 21/95 genera had all observations>0
# number of genera with no zero observations in cancer group
t1=table(apply(bdat[bdat$dgn==1,ngens],2,nz)==0); t1 #8 - i.e. 8/95 genera had all observations>0
# number of genera with no zero observations in IBS group
t2=table(apply(bdat[bdat$dgn==2,ngens],2,nz)==0); t2 #9 - i.e. 9/95 genera had all observations>0
# number of genera with no zero observations in ME/CFS group
t6=table(apply(bdat[bdat$dgn==6,ngens],2,nz)==0); t6 # 11 - i.e. 11/95 genera had all observations>0

# do pairwise comparisons between ANHAC and every other group, using Fisher exact tests
# ANHAC vs cancer - lower proportion of genera with no zeros in the cancer group
t01=rbind(t0,t1); t01; fisher.test(t01) # OR = 0.33 (0.12-0.82) p=0.014
# ANHAC vs IBS - lower proportion of genera with no zeros in the IBS group
t02=rbind(t0,t2); t02; fisher.test(t02) # OR = 0.37 (0.14-0.91) p=0.027
# ANHAC vs ME/CFS - similar proportions of genera with no zeros in the ANHAC & ME/CFS groups
t06=rbind(t0,t6); t06; fisher.test(t06) # OR = 0.46 (0.19-1.08) p=0.08

# now compute variance of each genus in each diagnostic group
v1=apply(bdat[bdat$dgn==1,-c(1:7)],2,var) # cancer
v2=apply(bdat[bdat$dgn==2,-c(1:7)],2,var) # IBS
v0=apply(bdat[bdat$dgn==4,-c(1:7)],2,var) # ANHAC
v6=apply(bdat[bdat$dgn==6,-c(1:7)],2,var) # ME/CFS
dvs=cbind(v0,v2,v6,v1); round(dvs[1:20,],2) # compare Table S1
# compare genus-wise variances in ANHAC and cancer groupings
wilcox.test(v0,v1,paired=T) # p = 0.73
# compare genus-wise variances in ANHAC and IBS groups
wilcox.test(v0,v2,paired=T) # p = 0.35
# compare genus-wise variances in ANHAC and ME/CFS groups
wilcox.test(v0,v6,paired=T) # p = 0.55

# now home in on variance of non-zero abundances for each genus in each group
zvar=function(x) return(var(x[x>0]))
zv1=apply(bdat[bdat$dgn==1,-c(1:7)],2,zvar) # cancer
zv2=apply(bdat[bdat$dgn==2,-c(1:7)],2,zvar) # IBS
zv0=apply(bdat[bdat$dgn==4,-c(1:7)],2,zvar) # ANHAC
zv6=apply(bdat[bdat$dgn==6,-c(1:7)],2,zvar) # ME/CFS
dzvs=cbind(zv0,zv1,zv2,zv6); dzvs[1:20,]; table(apply(dzvs,1,min)==0)
# compare genus-wise variances in ANHAC and cancer groupings
wilcox.test(zv0,zv1,paired=T) # p = 0.44
# compare genus-wise variances in ANHAC and IBS groupings
wilcox.test(zv0,zv2,paired=T) # p = 0.27
# compare genus-wise variances in ANHAC and ME/CFS groupings
wilcox.test(zv0,zv6,paired=T) # p = 0.96
# conclusion - no difference in levels of variance between groups
# i.e. no heteroscedasticity


######################################################################
#= END OF R CODE TO CHECK HETEROSCEDASTICITY ======================#
#====================================================================#
#====================================================================#
#====================================================================#
#====================================================================#
#====================================================================#
#====================================================================#
#====================================================================#
#====================================================================#
######################################################################
