## Supplementary figures and images for "An approach to finding specific forms of dysbiosis that associate with different disorders"

### Examples of decision trees of interaction between genera

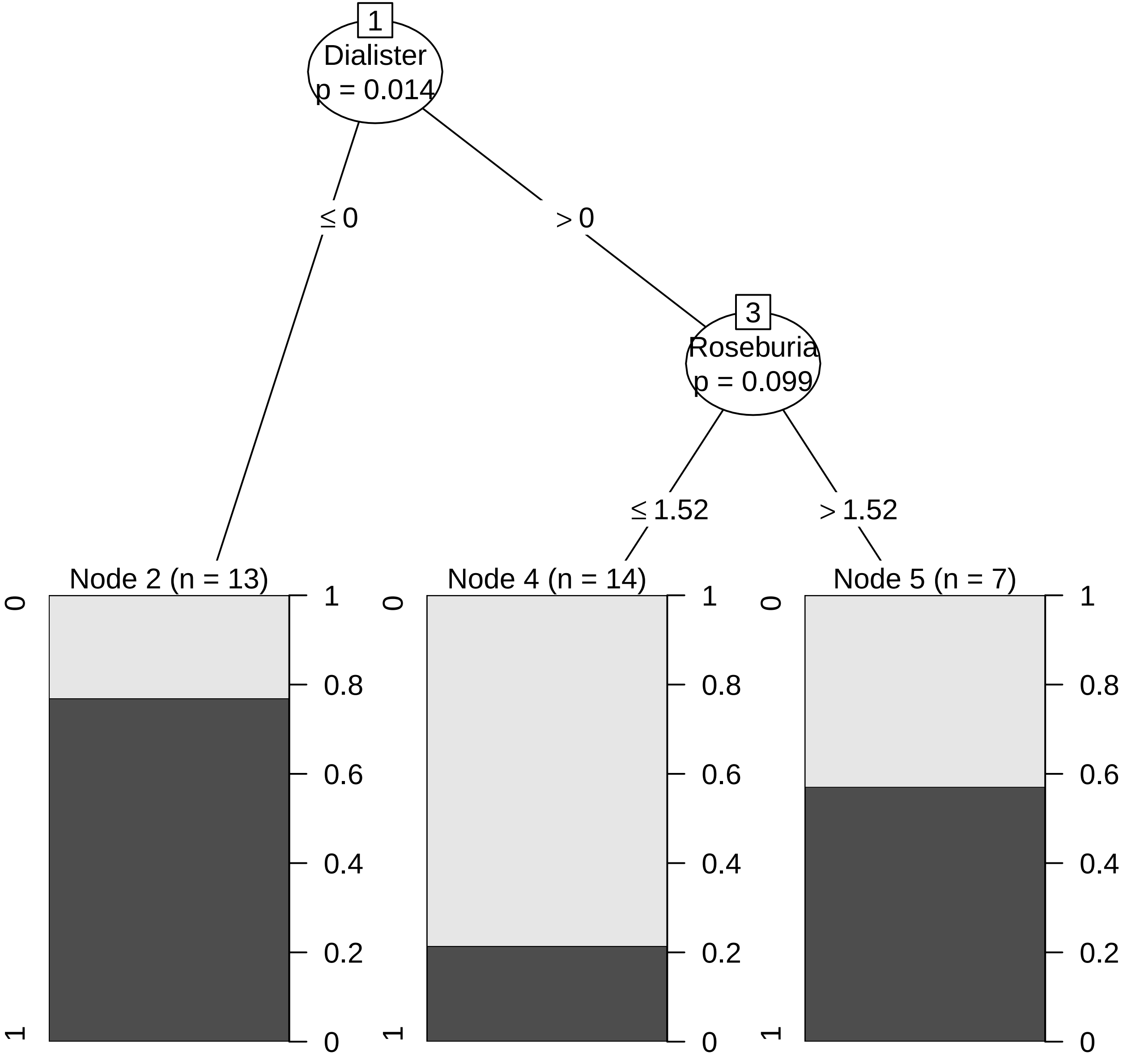

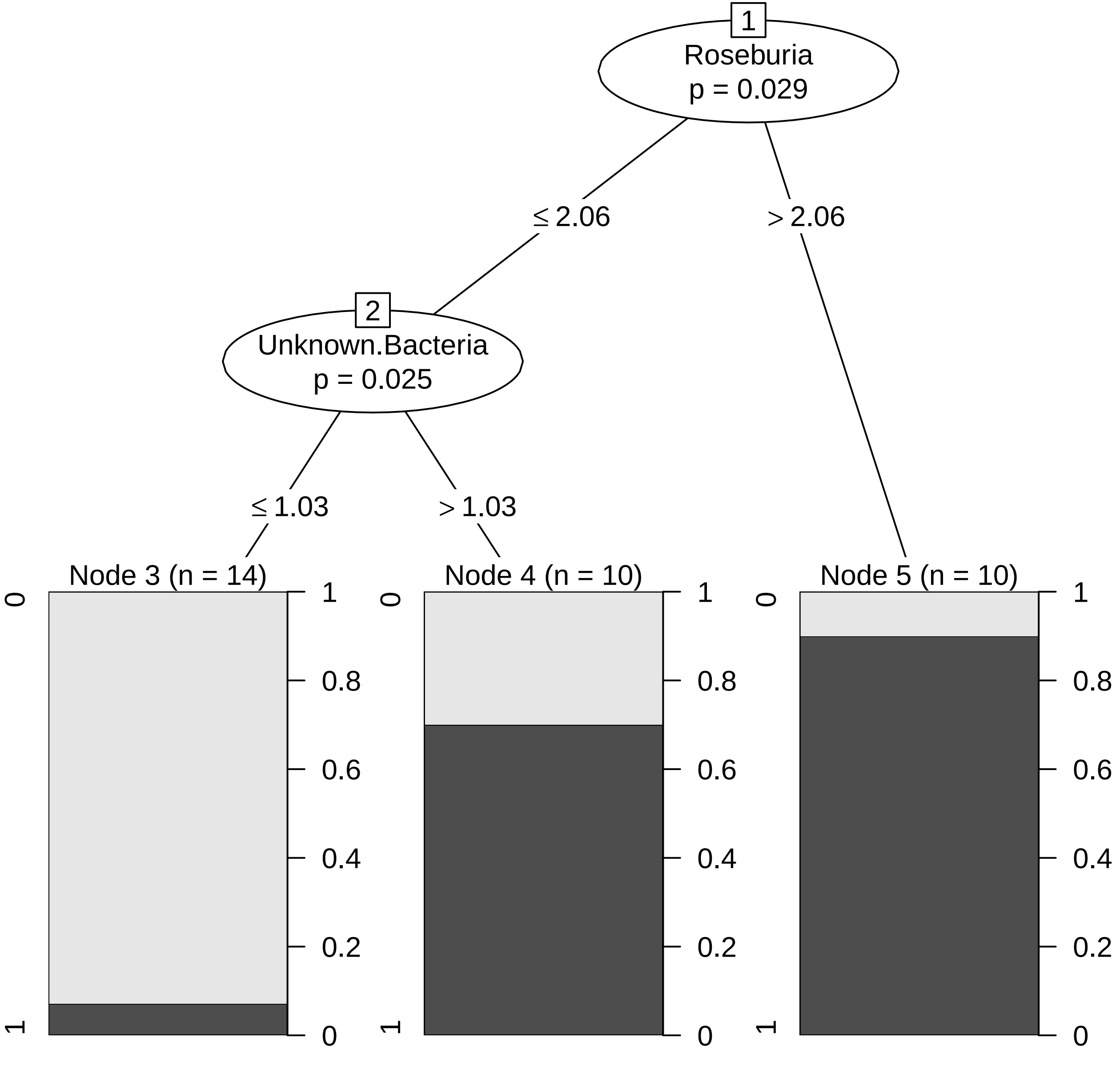
