## Supplementary material for "An approach to finding specific forms of dysbiosis that associate with different disorders": AUROCs and partial plots

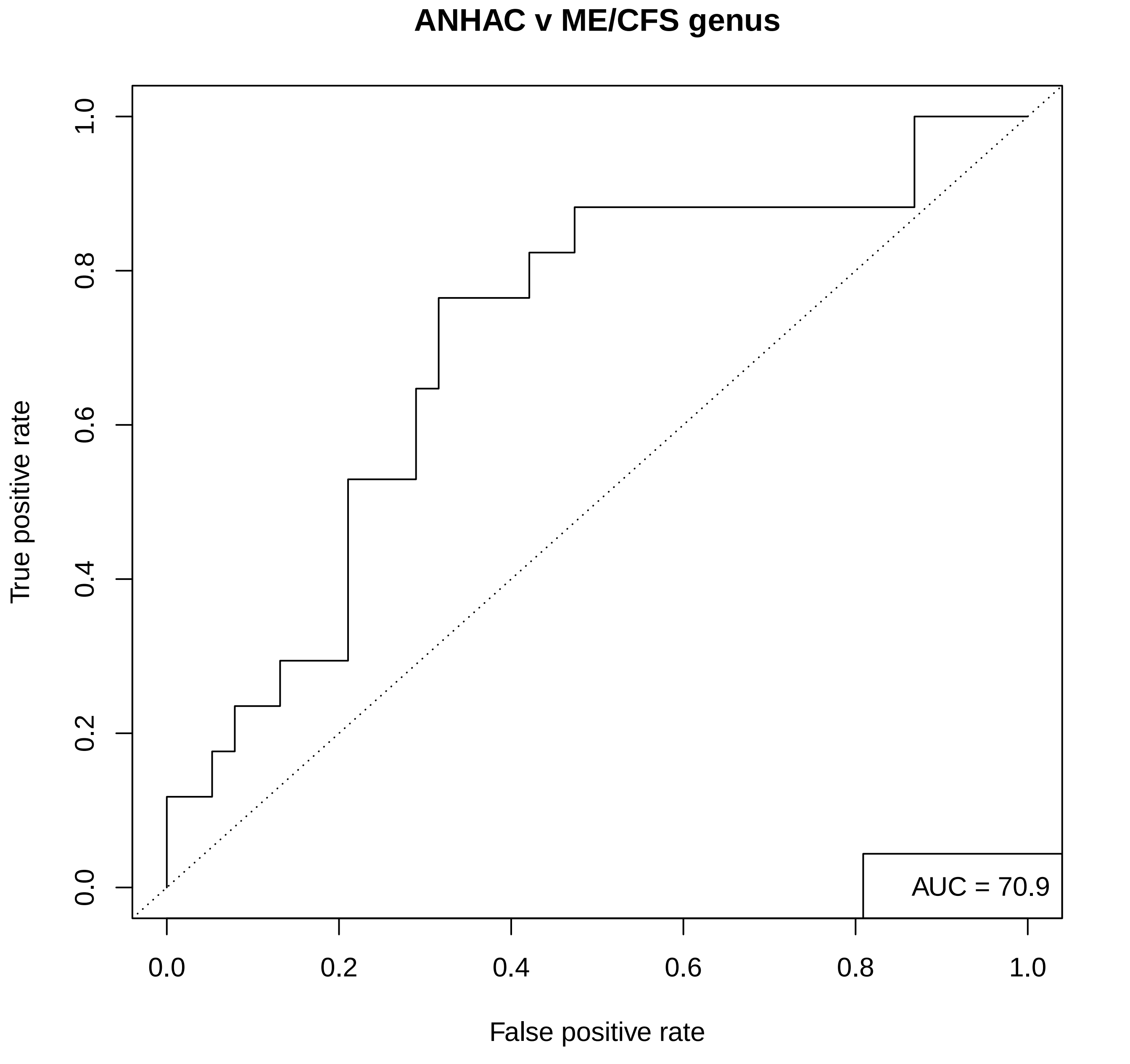
ANHAC v ME/CFS genus
True positive rate
0.0
0.2
0.4
0.6
0.8
1.0
False positive rate
AUC = 70.9

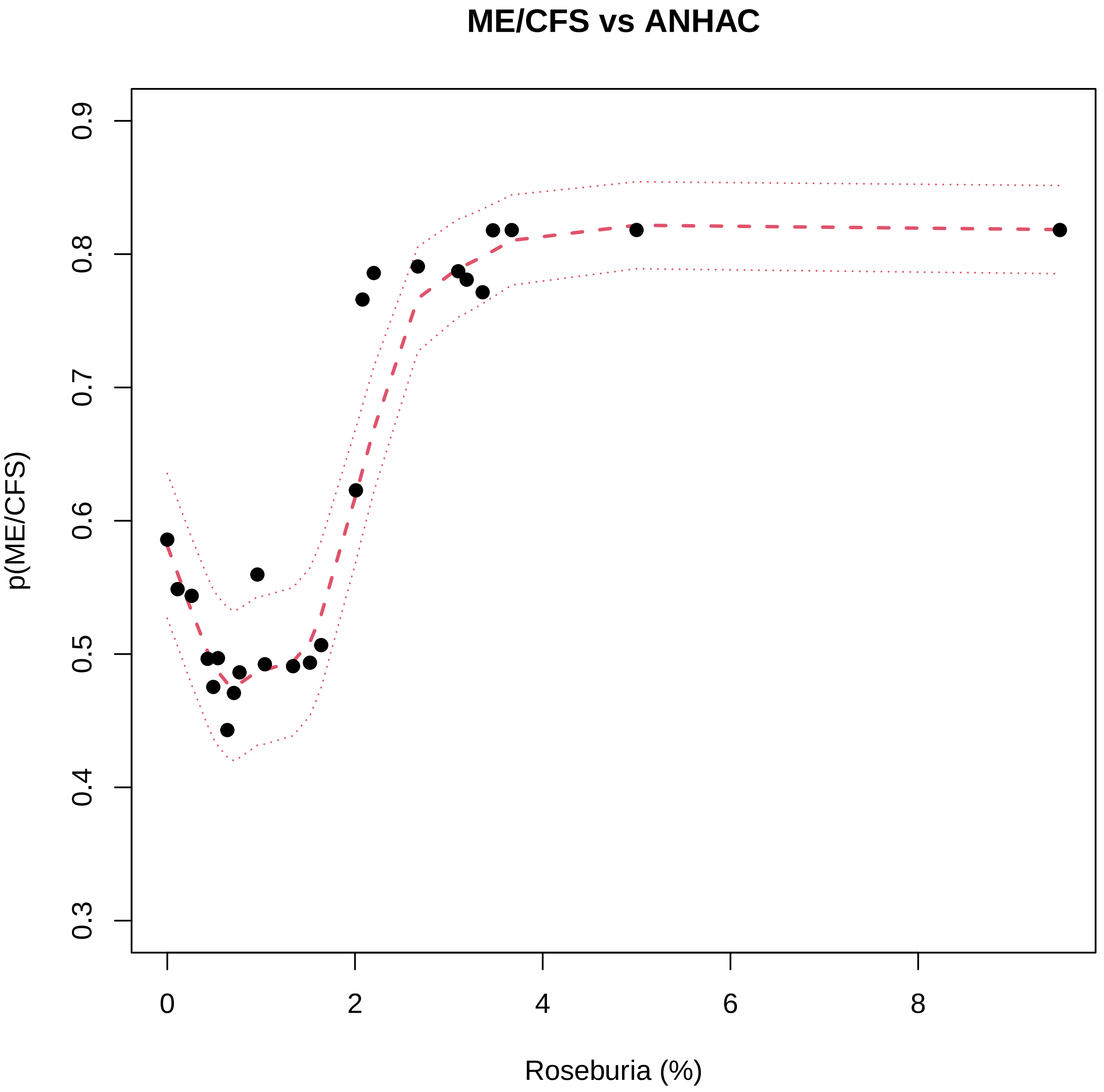

ME/CFS vs ANHAC
p(ME/CFS)
0.3
0.4
0.5
0.6
0.7
0.8
0.9
0
2
4
6
8
Roseburia (%)

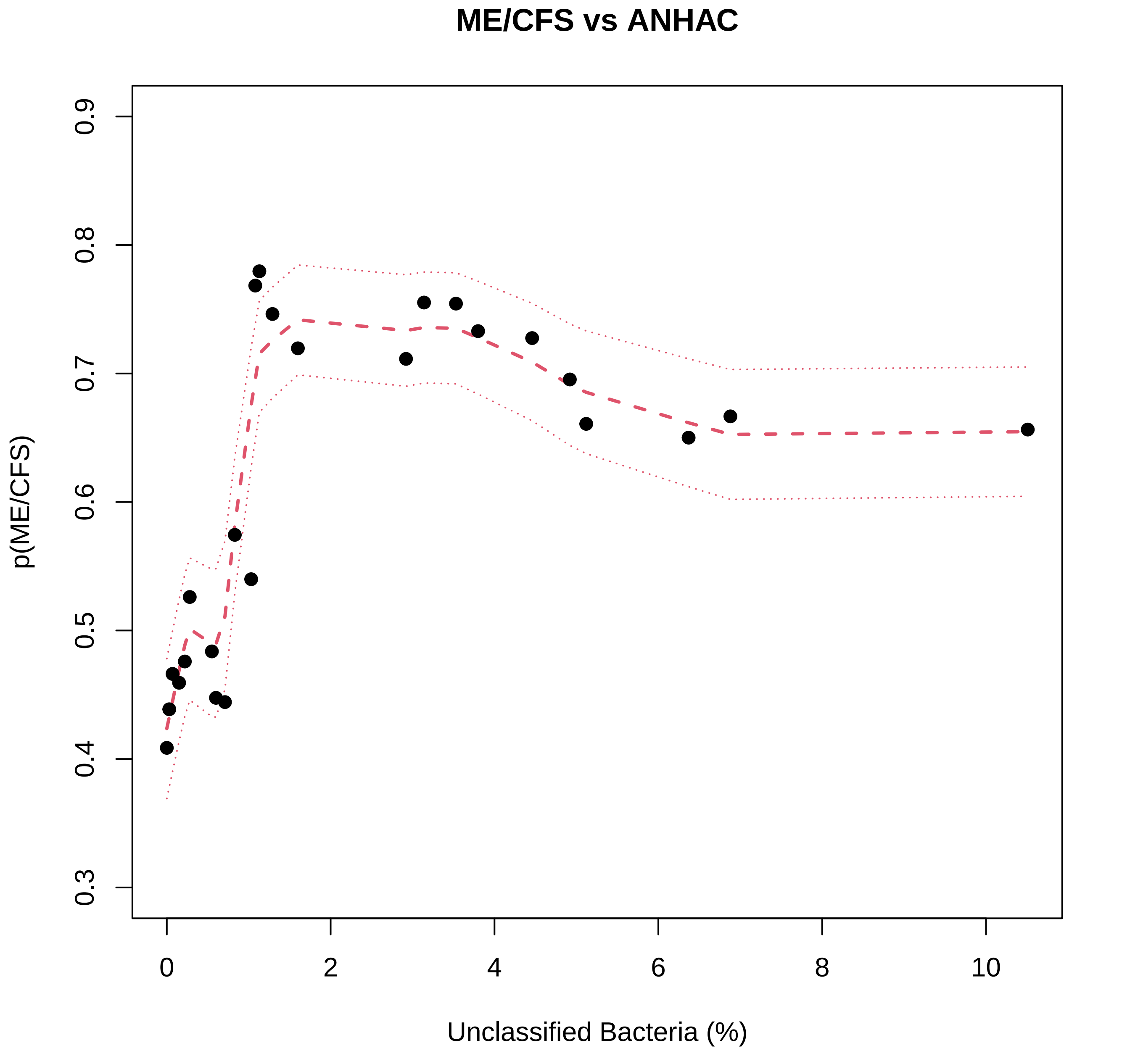
ME/CFS vs ANHAC
p(ME/CFS)
0.3
0.4
0.5
0.6
0.7
0.8
0.9
0
2
4
6
8
10
Unclassified Bacteria (%)

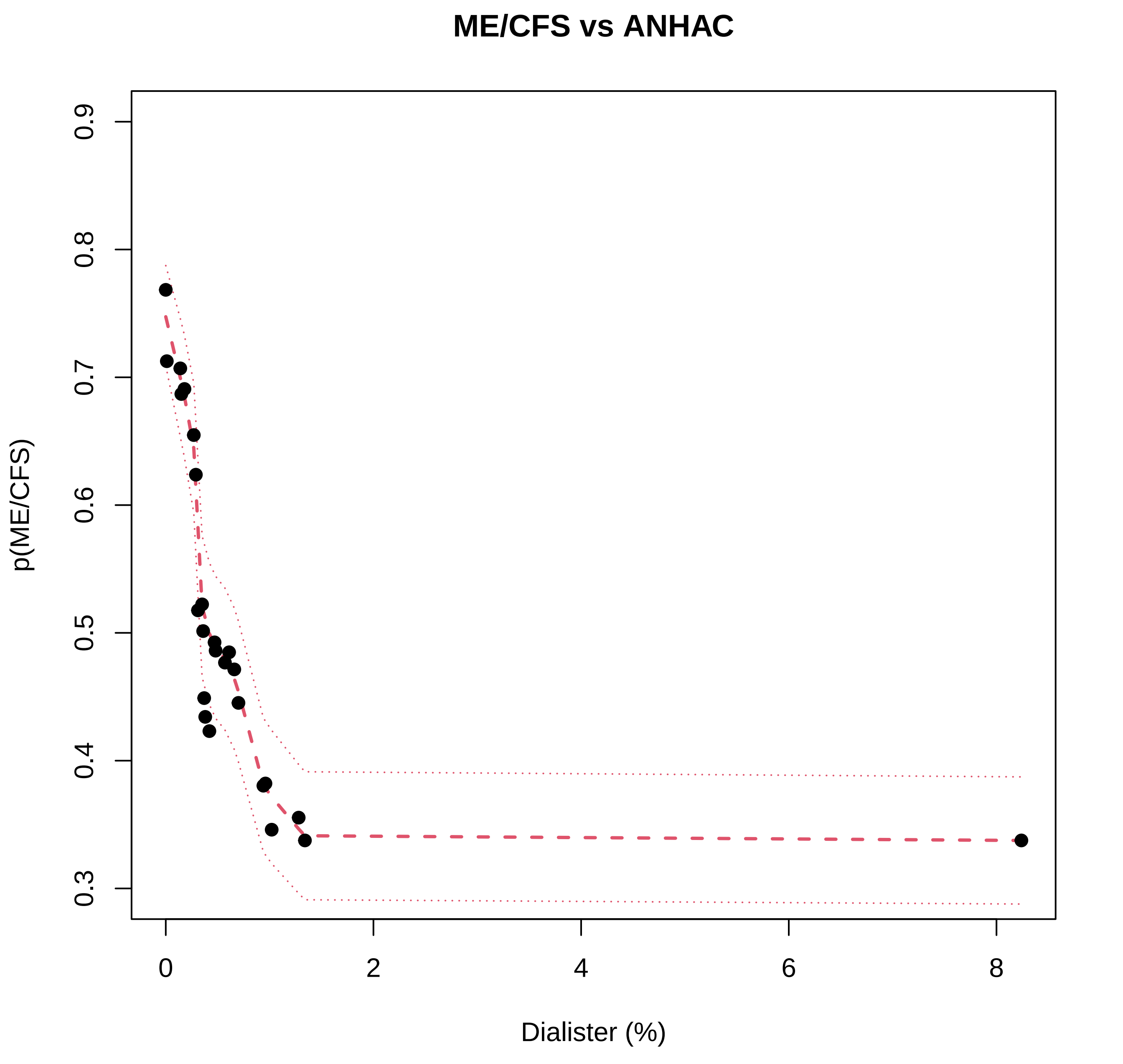
ME/CFS vs ANHAC
p(ME/CFS)
0.3
0.4
0.5
0.6
0.7
0.8
0.9
0
2
4
6
8
Dialister (%)

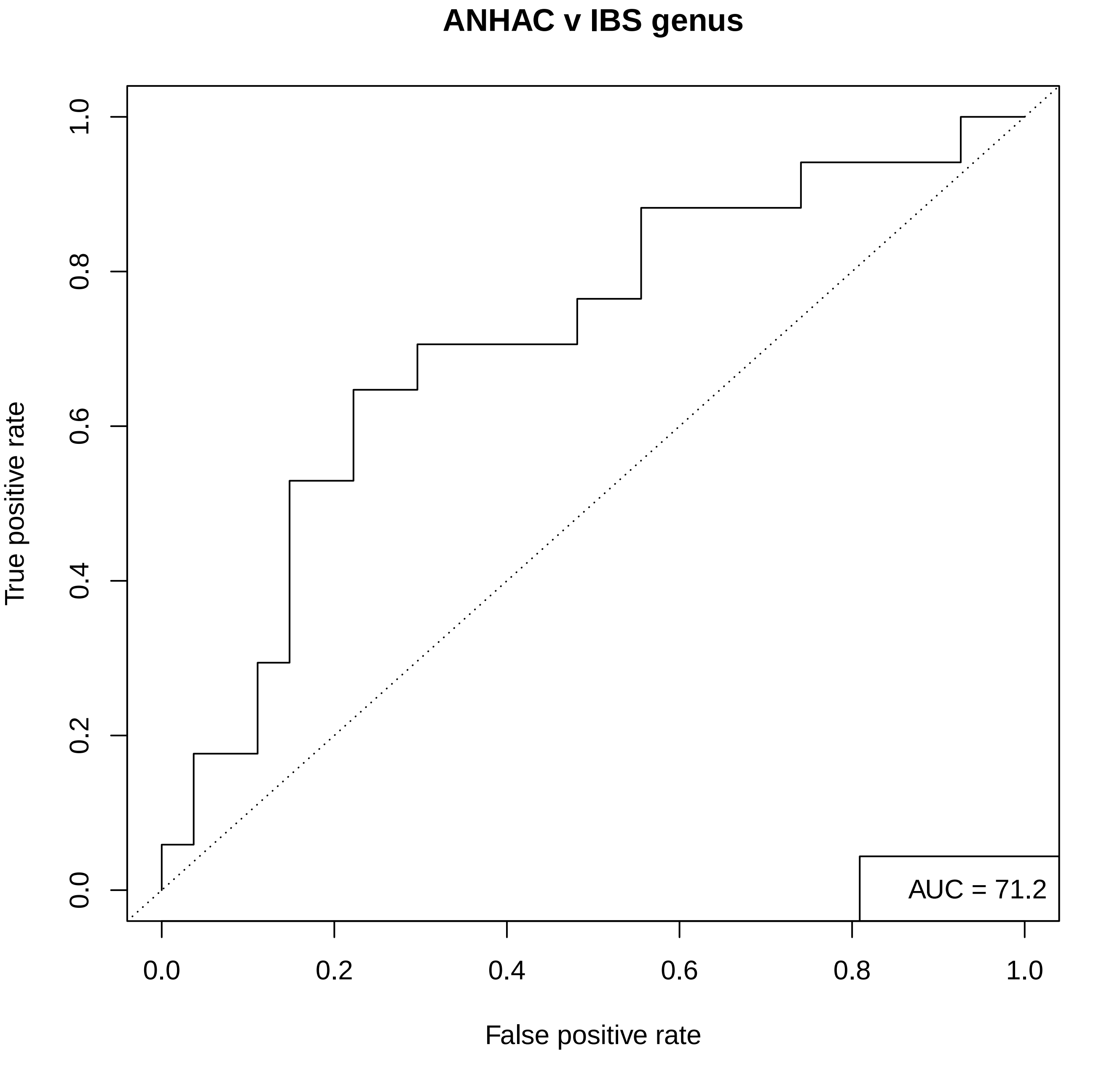
ANHAC v IBS genus
True positive rate
False positive rate
0.0
0.2
0.4
0.6
0.8
1.0
AUC = 71.2

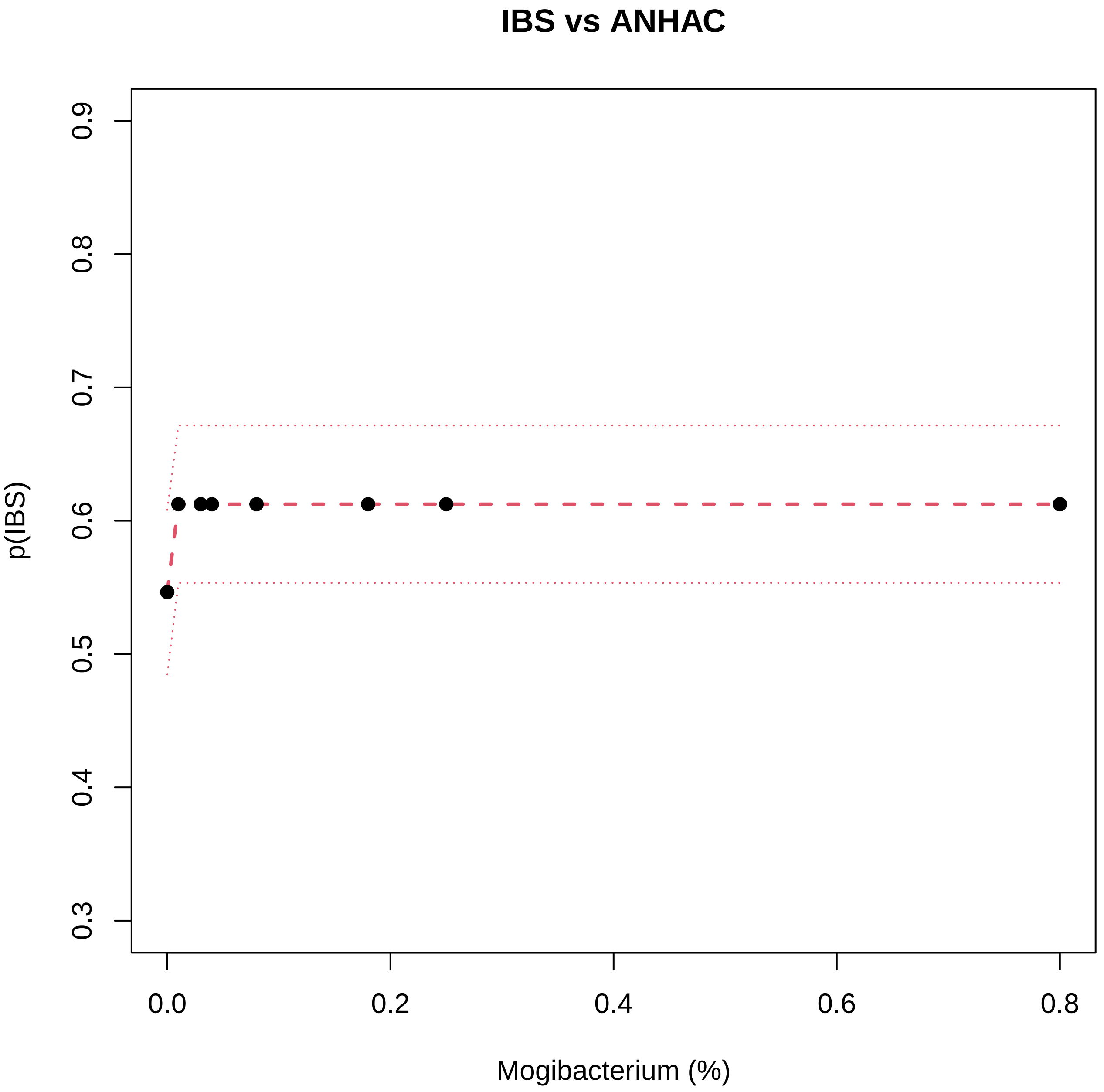

IBS vs ANHAC
p(IBS)
0.3
0.4
0.5
0.6
0.7
0.8
0.9
0.0
0.2
0.4
0.6
0.8
Mogibacterium (%)

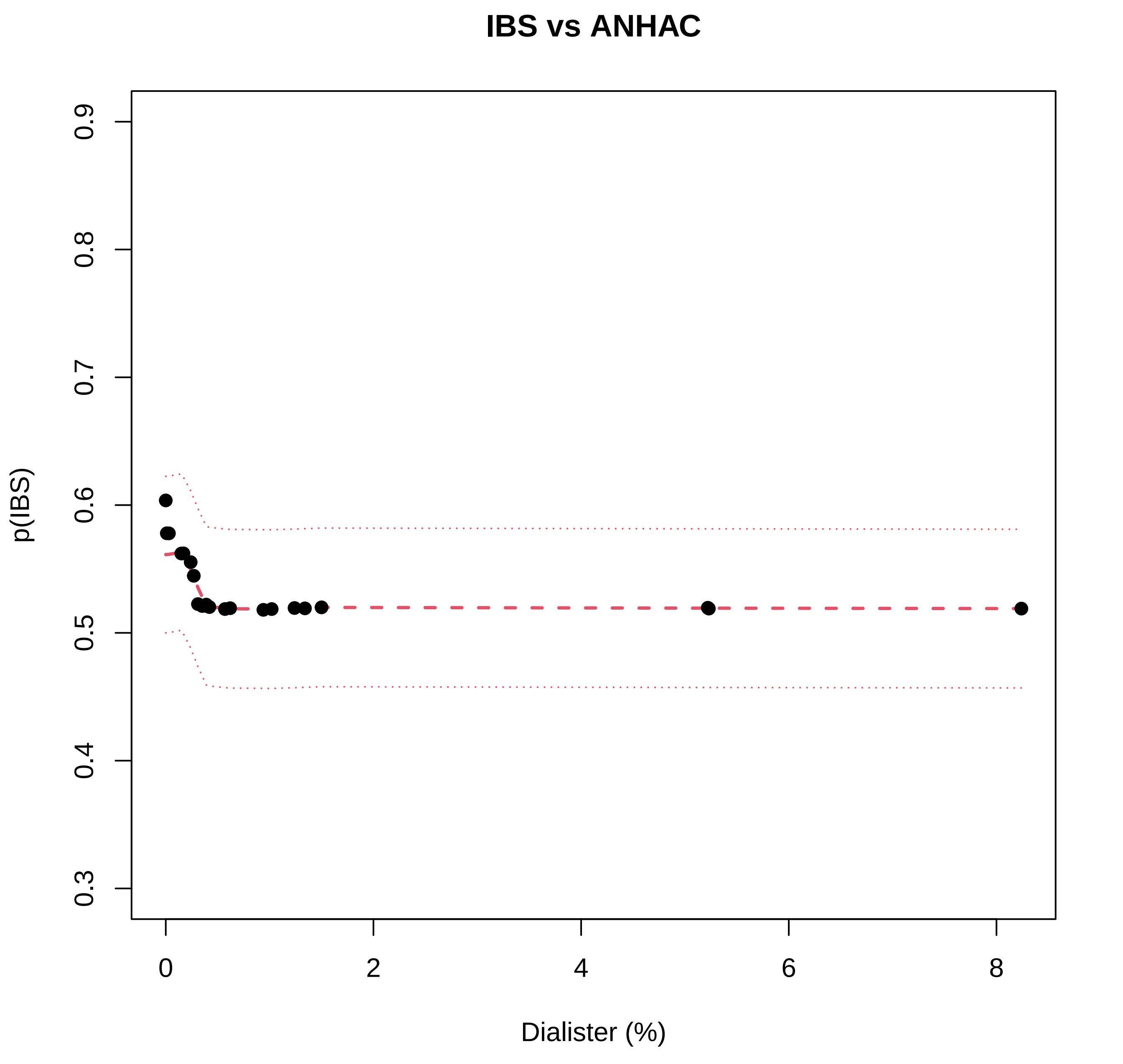
IBS vs ANHAC
p(IBS)
0.3
0.4
0.5
0.6
0.7
0.8
0.9
0
2
4
6
8
Dialister (%)

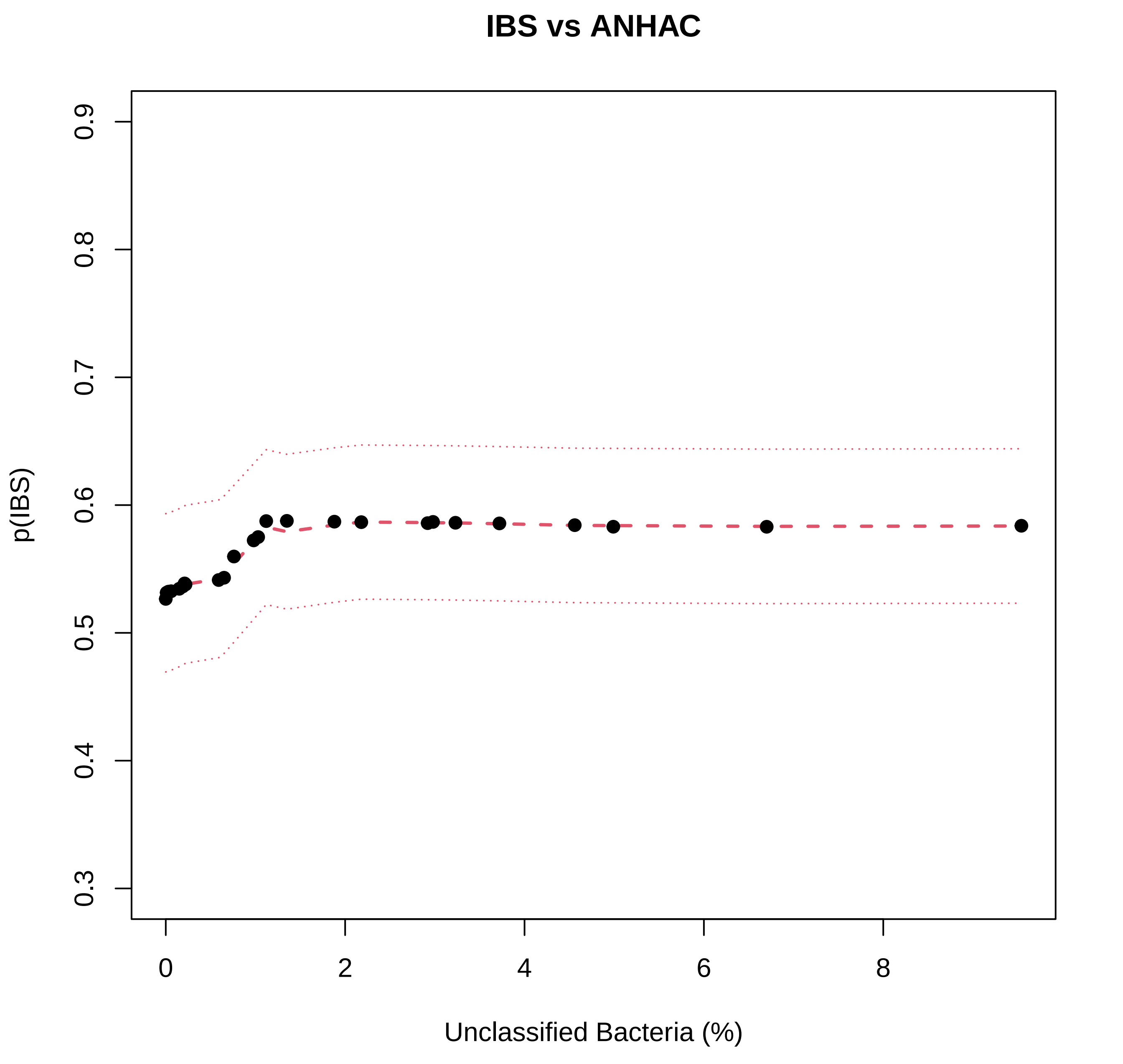
IBS vs ANHAC
p(IBS)
0.3
0.4
0.5
0.6
0.7
0.8
0.9
0
2
4
6
8
Unclassified Bacteria (%)

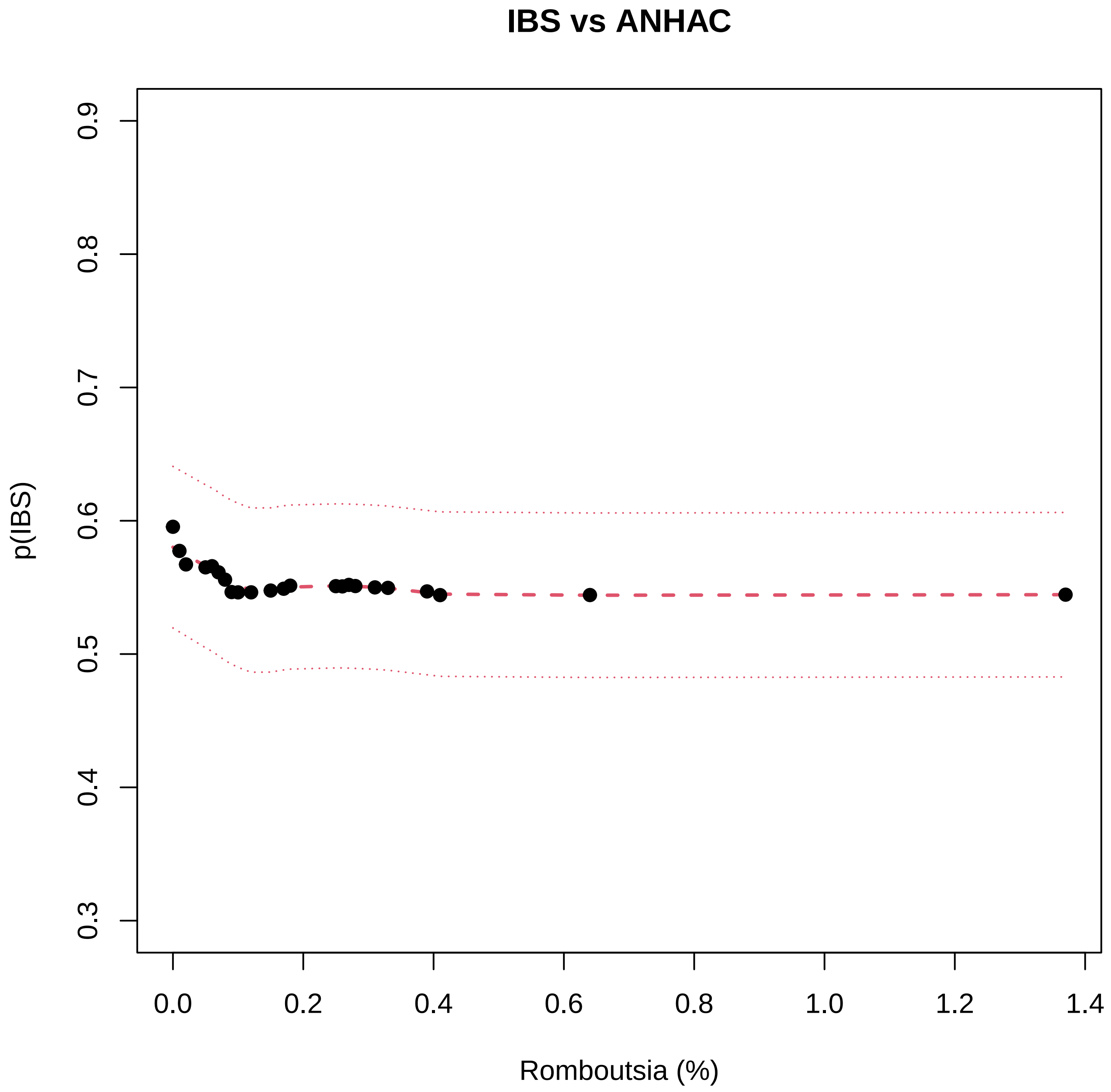

IBS vs ANHAC
p(IBS)
0.3
0.4
0.5
0.6
0.7
0.8
0.9
0.0
0.2
0.4
0.6
0.8
1.0
1.2
1.4
Romboutsia (%)

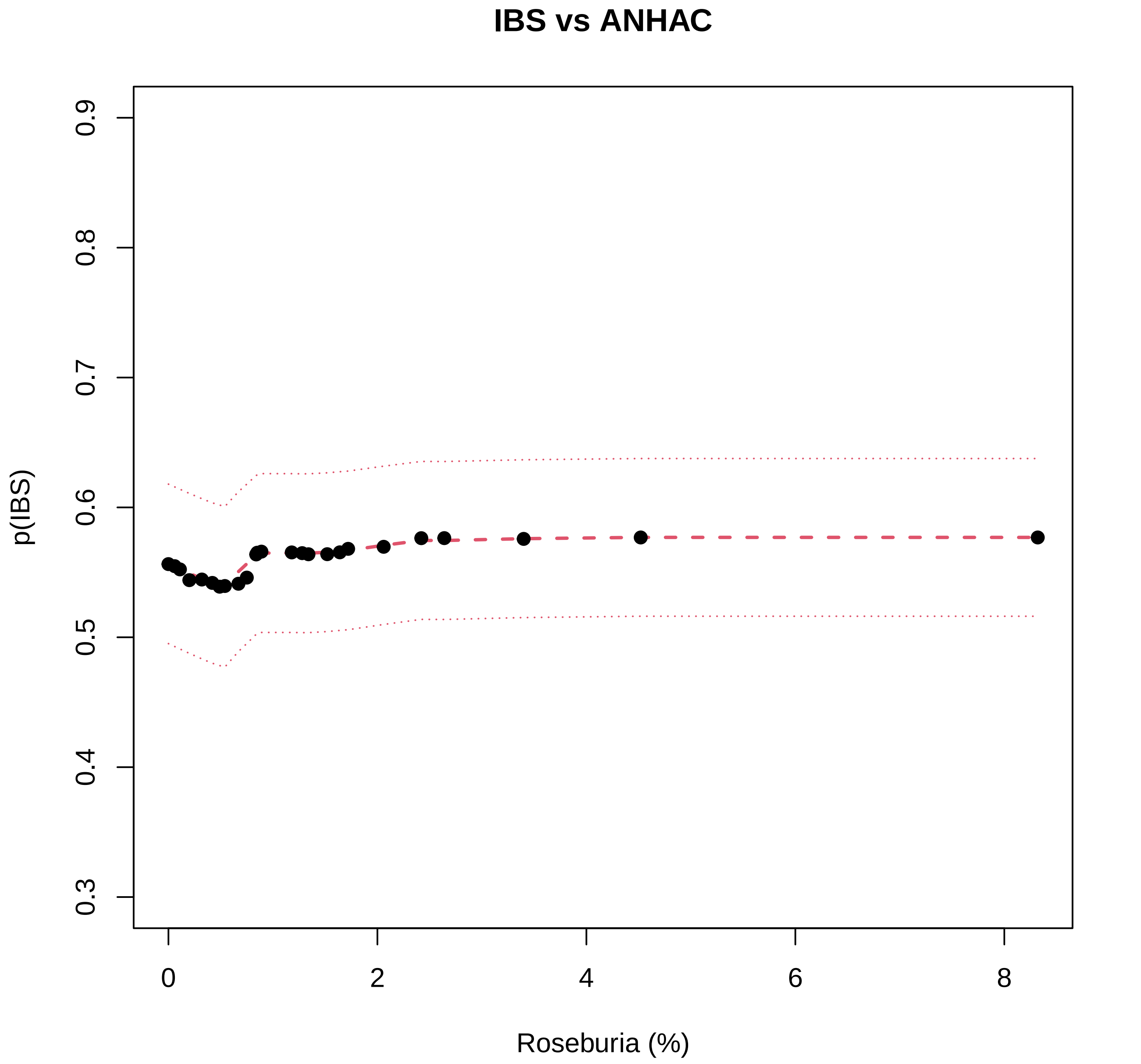

IBS vs ANHAC
p(IBS)
0.3
0.4
0.5
0.6
0.7
0.8
0.9
0
2
4
6
8
Roseburia (%)

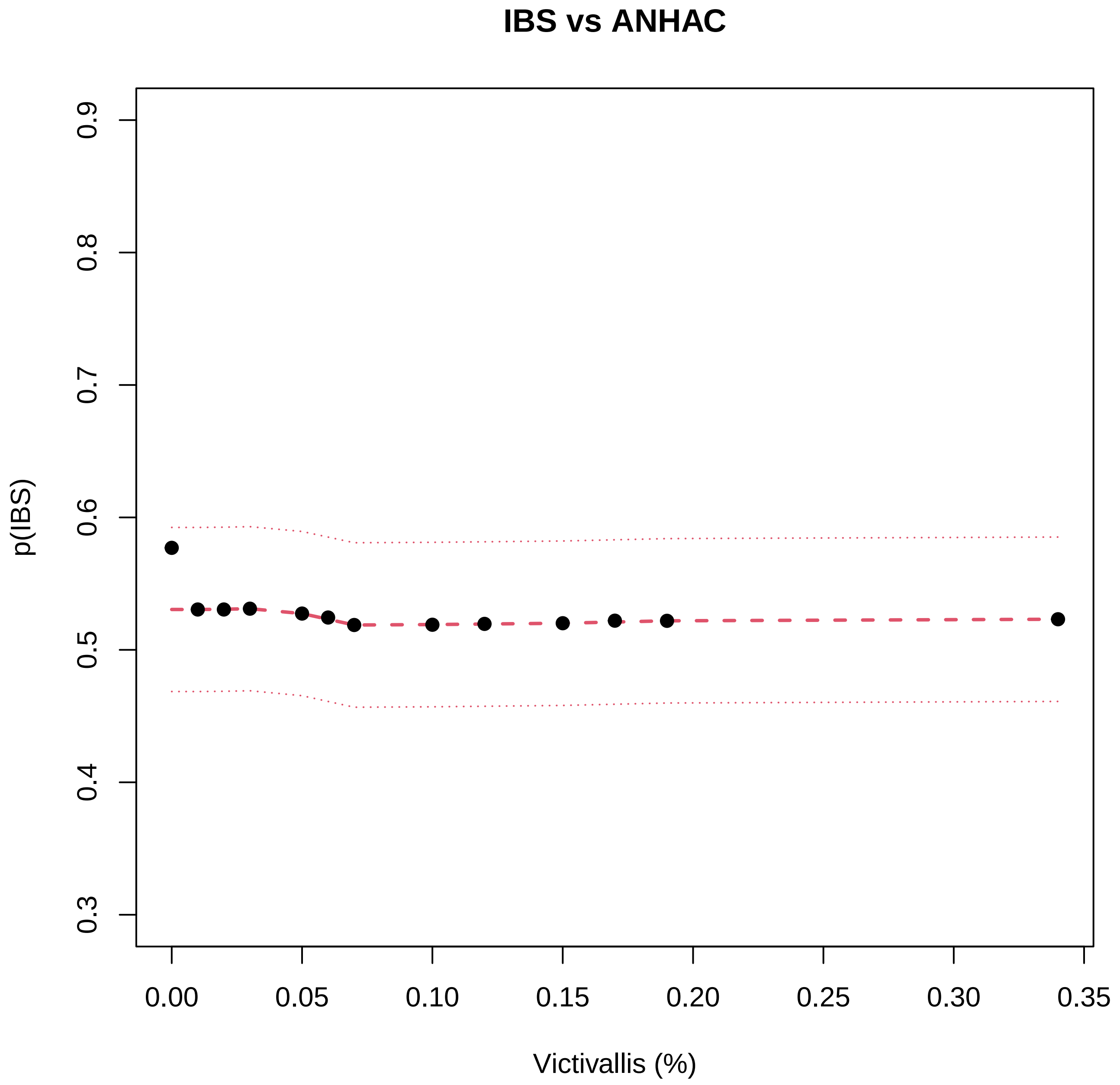
IBS vs ANHAC
p(IBS)
0.3
0.4
0.5
0.6
0.7
0.8
0.9
0.00
0.05
0.10
0.15
0.20
0.25
0.30
0.35
Victivallis (%)

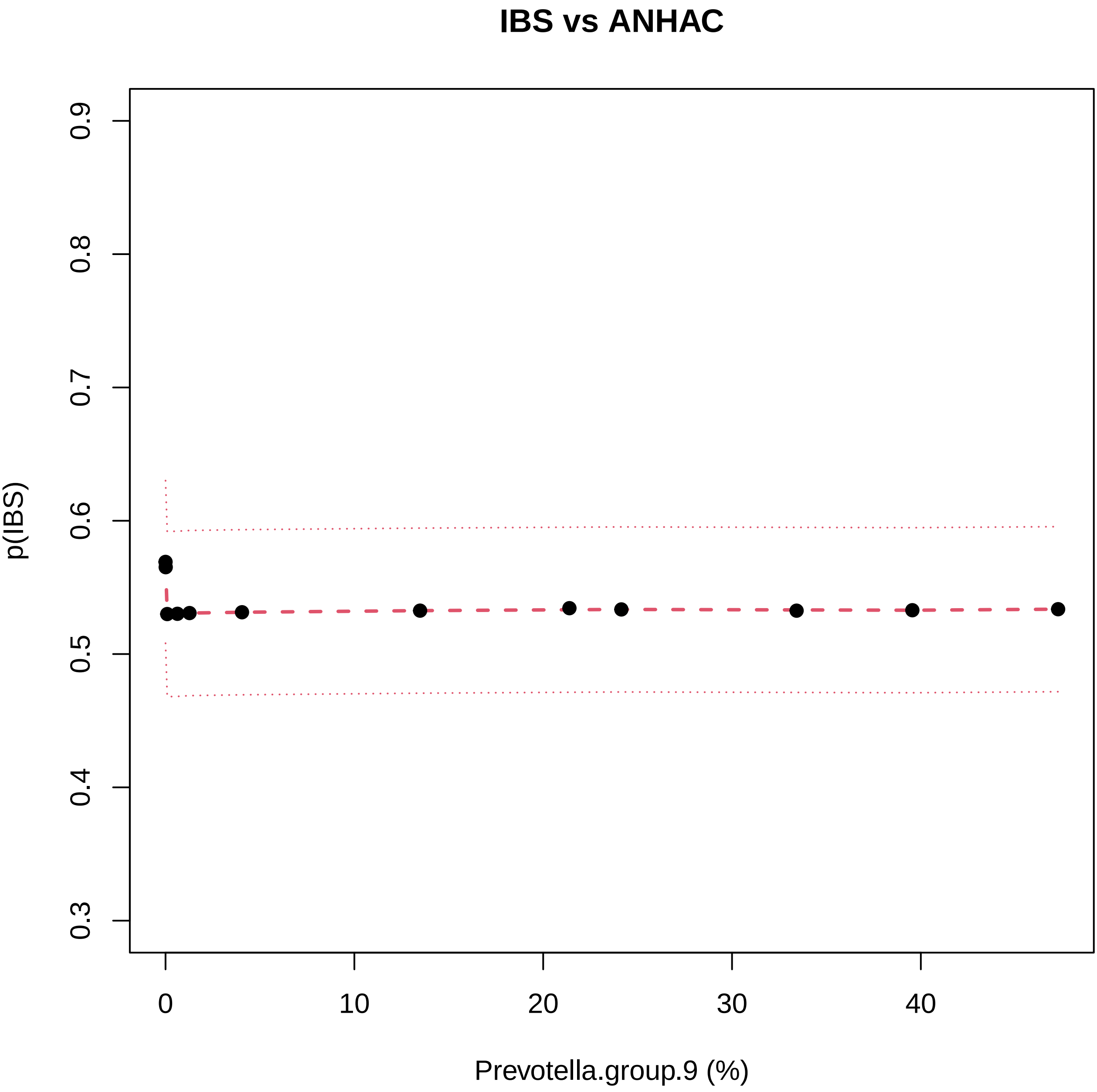
IBS vs ANHAC
p(IBS)
0.3
0.4
0.5
0.6
0.7
0.8
0.9
0
10
20
30
40
Prevotella.group.9 (%)

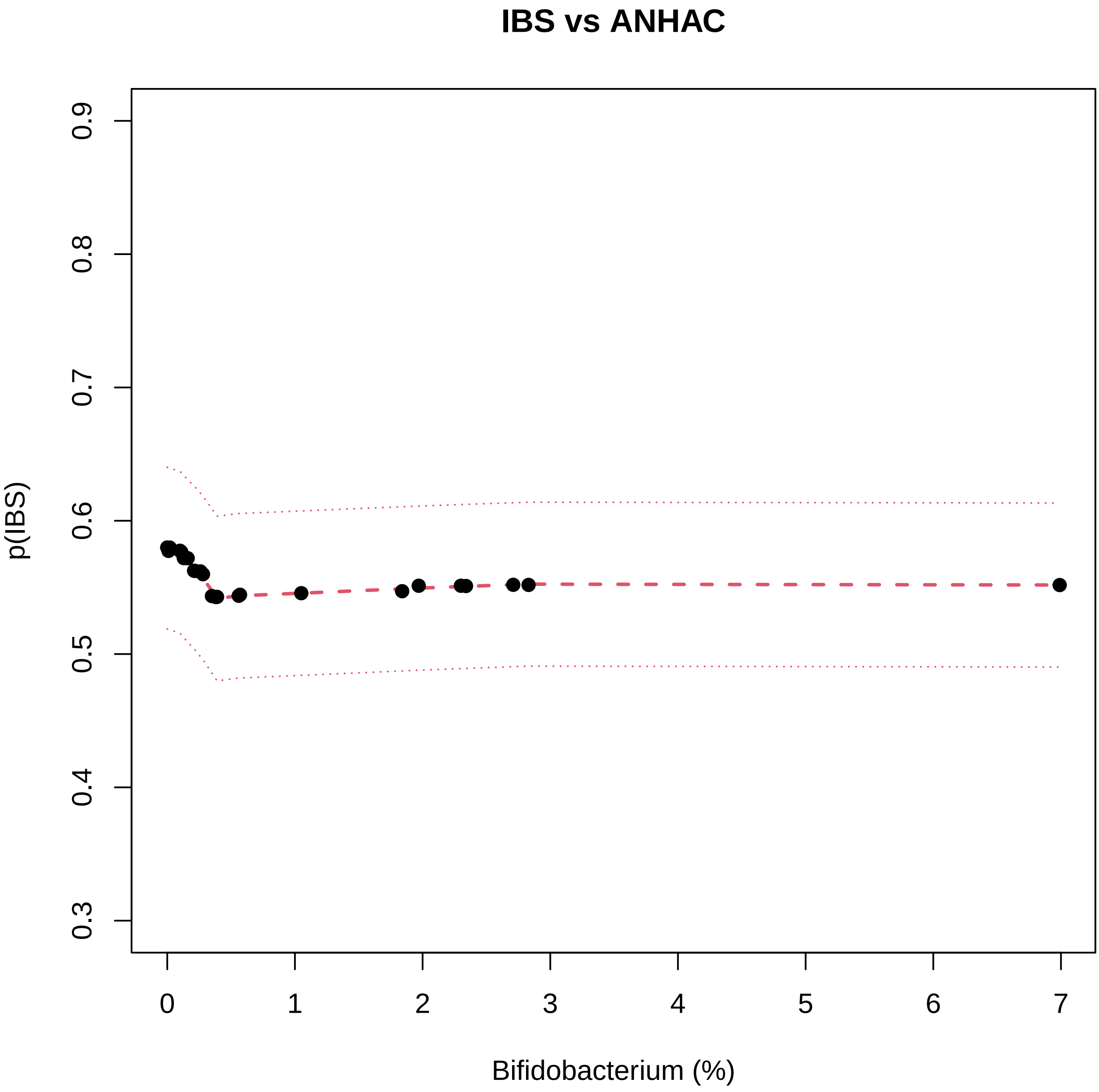
IBS vs ANHAC
p(IBS)
0.3
0.4
0.5
0.6
0.7
0.8
0.9
0
1
2
3
4
5
6
7
Bifidobacterium (%)

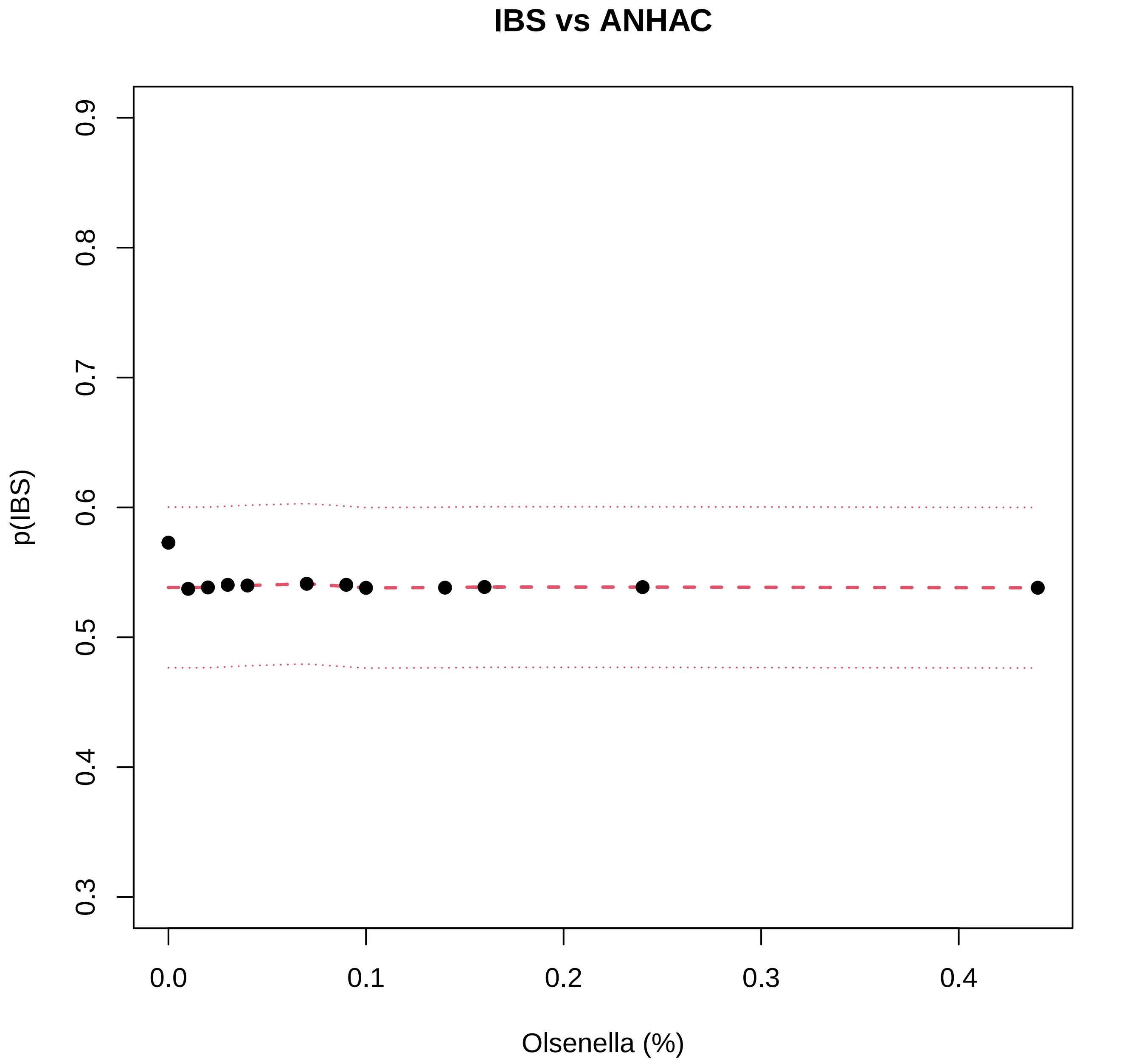
IBS vs ANHAC
p(IBS)
0.3
0.4
0.5
0.6
0.7
0.8
0.9
0.0
0.1
0.2
0.3
0.4
Olsenella (%)

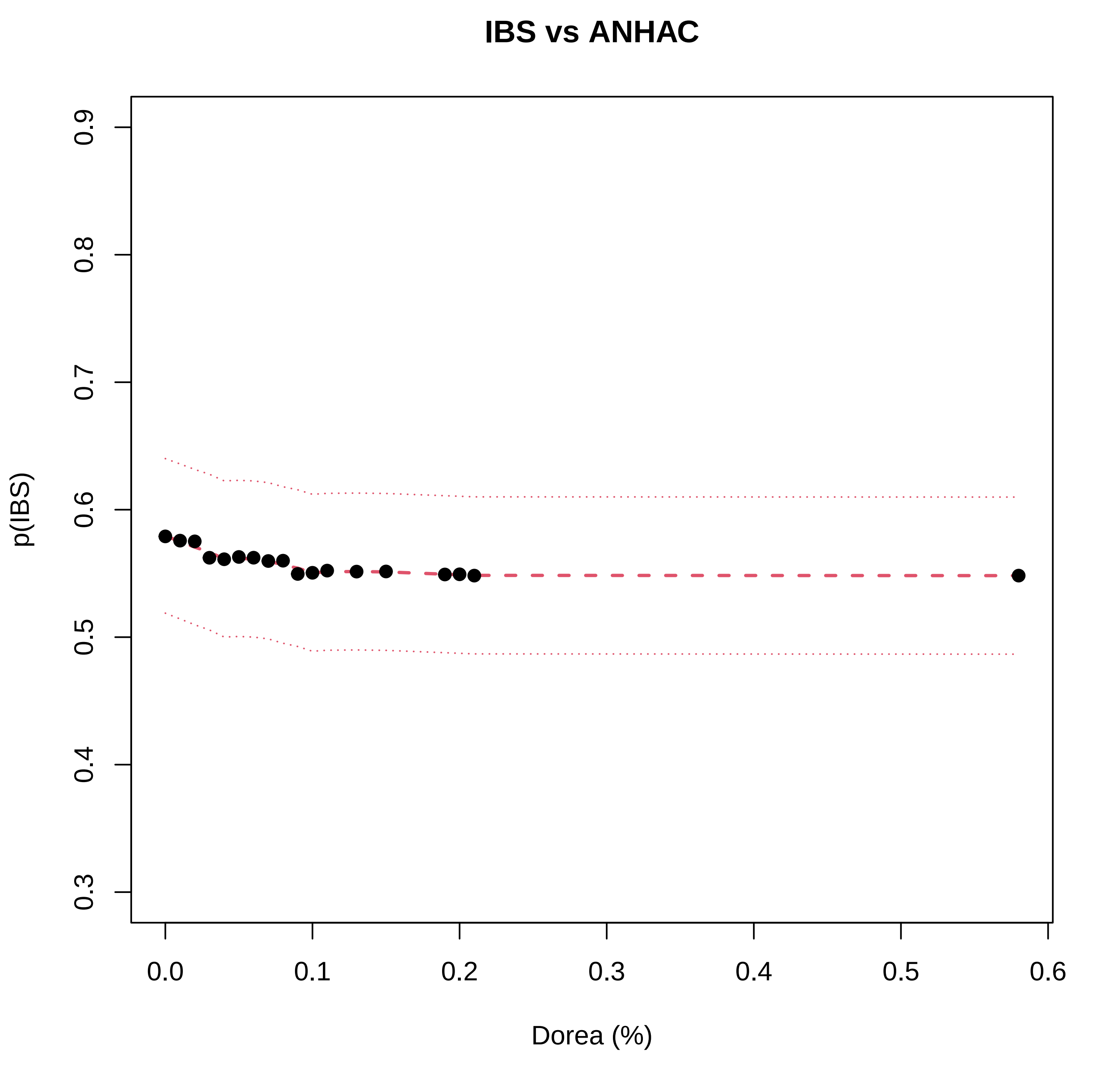
IBS vs ANHAC
p(IBS)
0.3
0.4
0.5
0.6
0.7
0.8
0.9
0.0
0.1
0.2
0.3
0.4
0.5
0.6
Dorea (%)

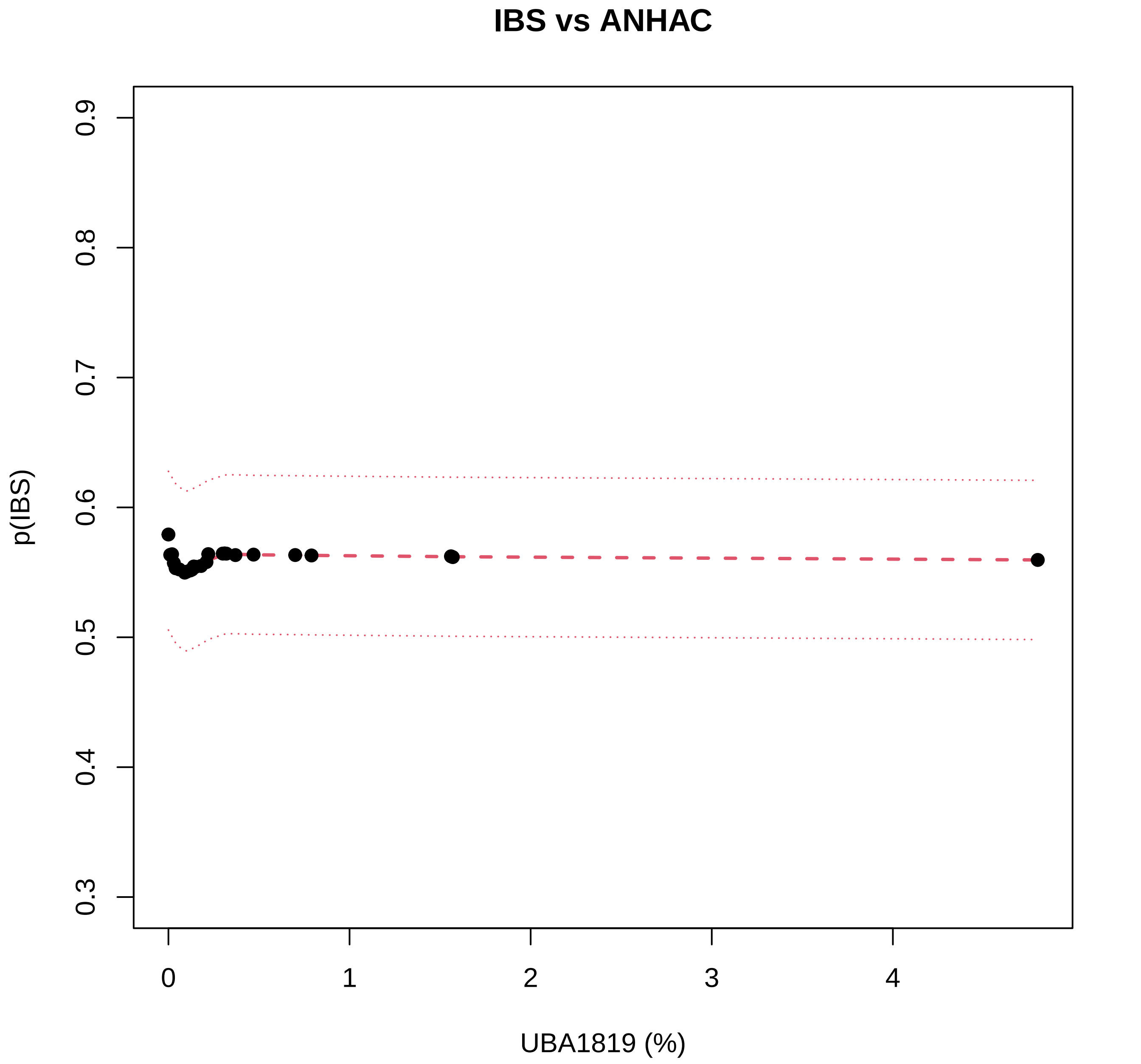
IBS vs ANHAC
p(IBS)
0.3
0.4
0.5
0.6
0.7
0.8
0.9
0
1
2
3
4
UBA1819 (%)

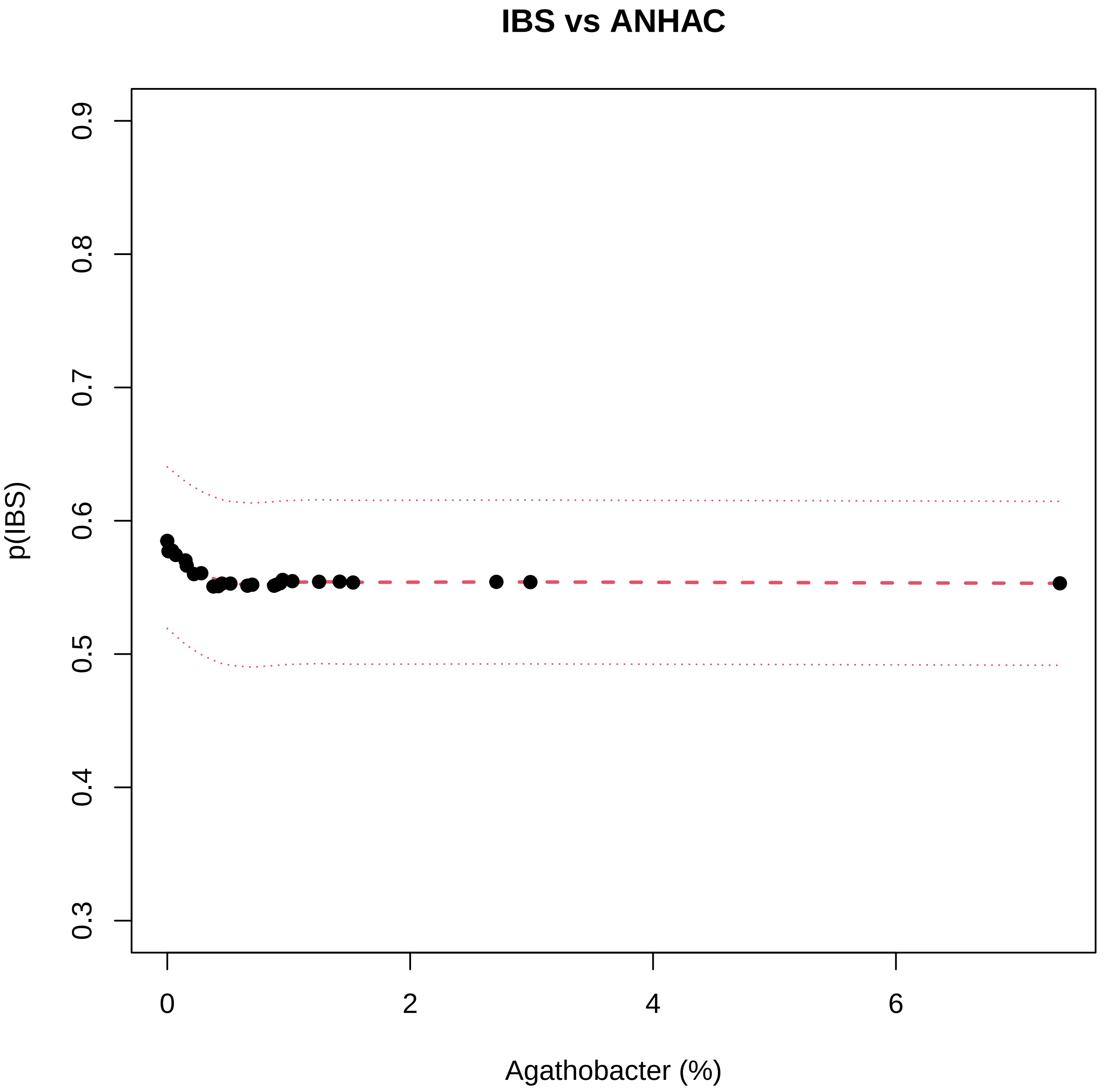
IBS vs ANHAC
p(IBS)
0.3
0.4
0.5
0.6
0.7
0.8
0.9
0
2
4
6
Agathobacter (%)

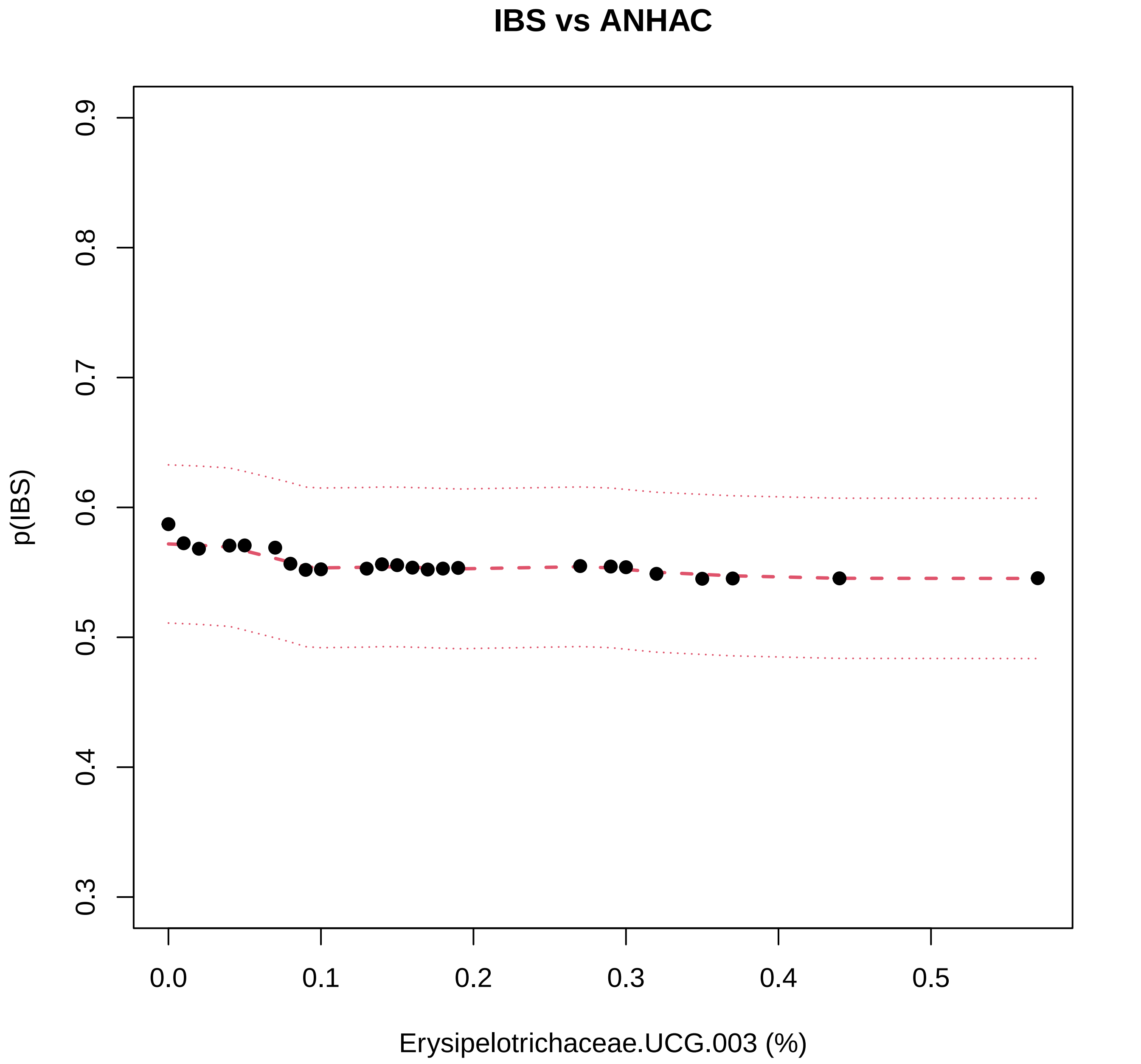
IBS vs ANHAC
p(IBS)
0.3
0.4
0.5
0.6
0.7
0.8
0.9
0.0
0.1
0.2
0.3
0.4
0.5
Erysipelotrichaceae.UCG.003 (%)

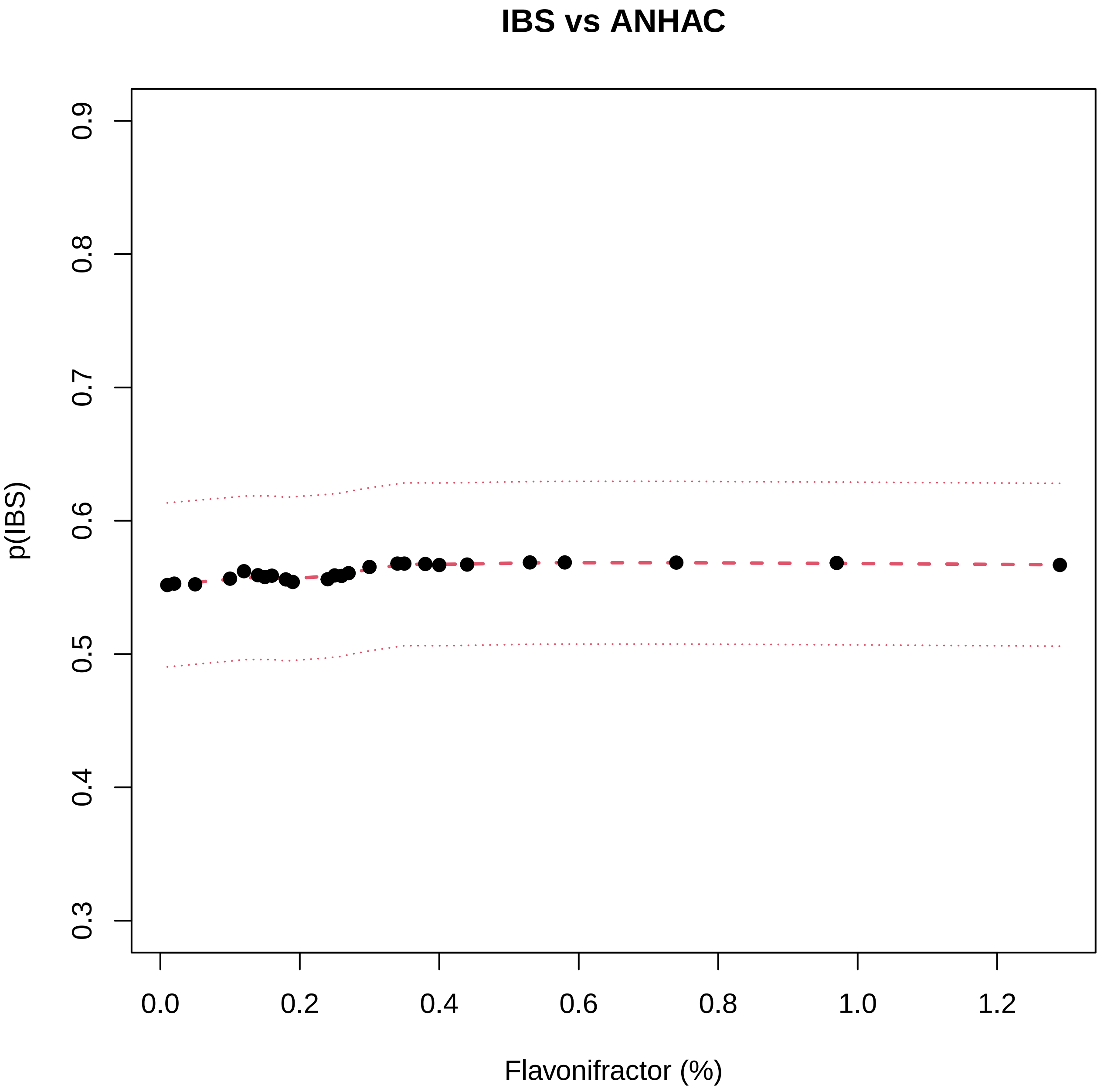

IBS vs ANHAC
p(IBS)
0.3
0.4
0.5
0.6
0.7
0.8
0.9
0.0
0.2
0.4
0.6
0.8
1.0
1.2
Flavonifractor (%)

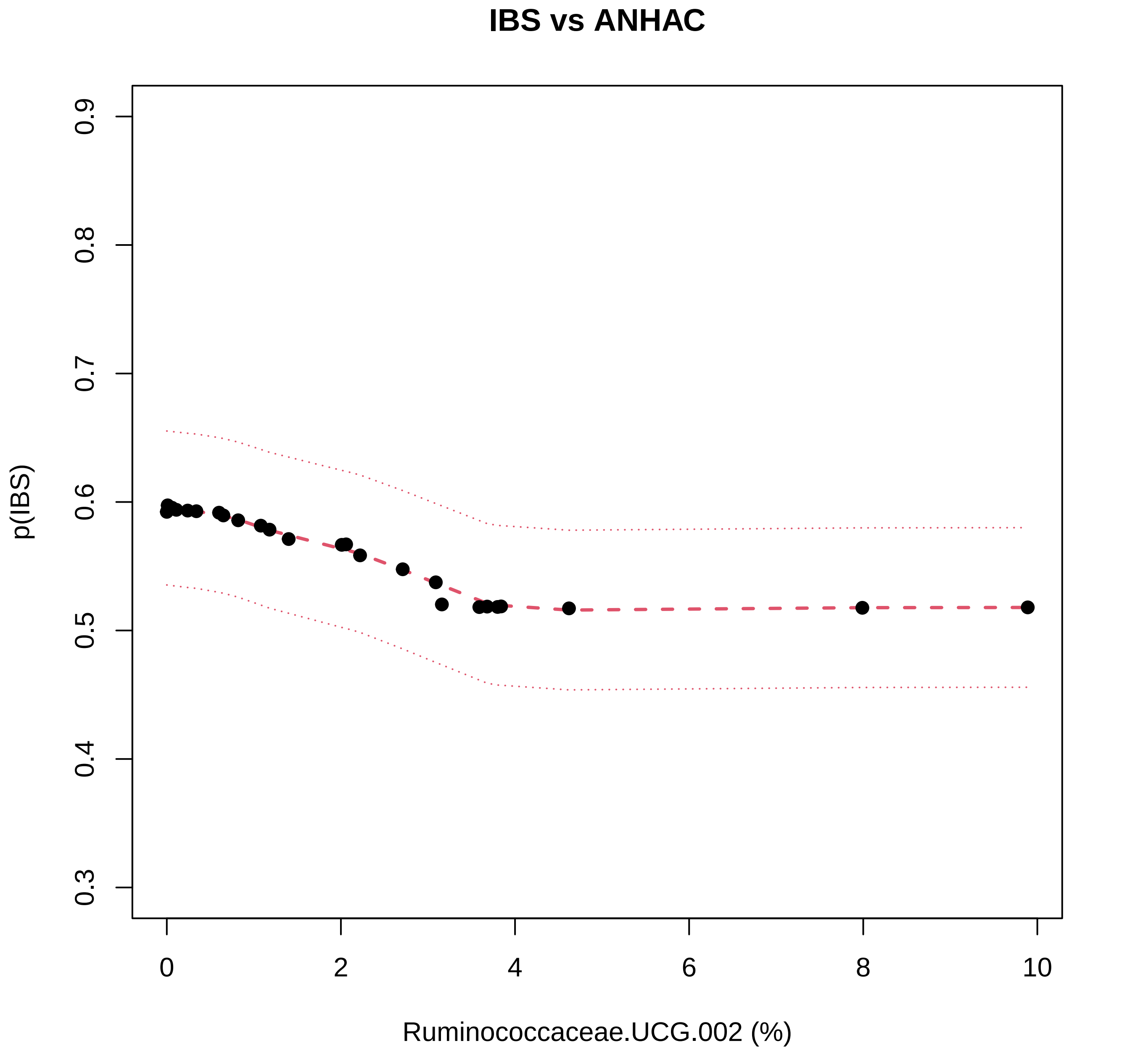
IBS vs ANHAC
p(IBS)
0.3
0.4
0.5
0.6
0.7
0.8
0.9
0
2
4
6
8
10
Ruminococcaceae.UCG.002 (%)

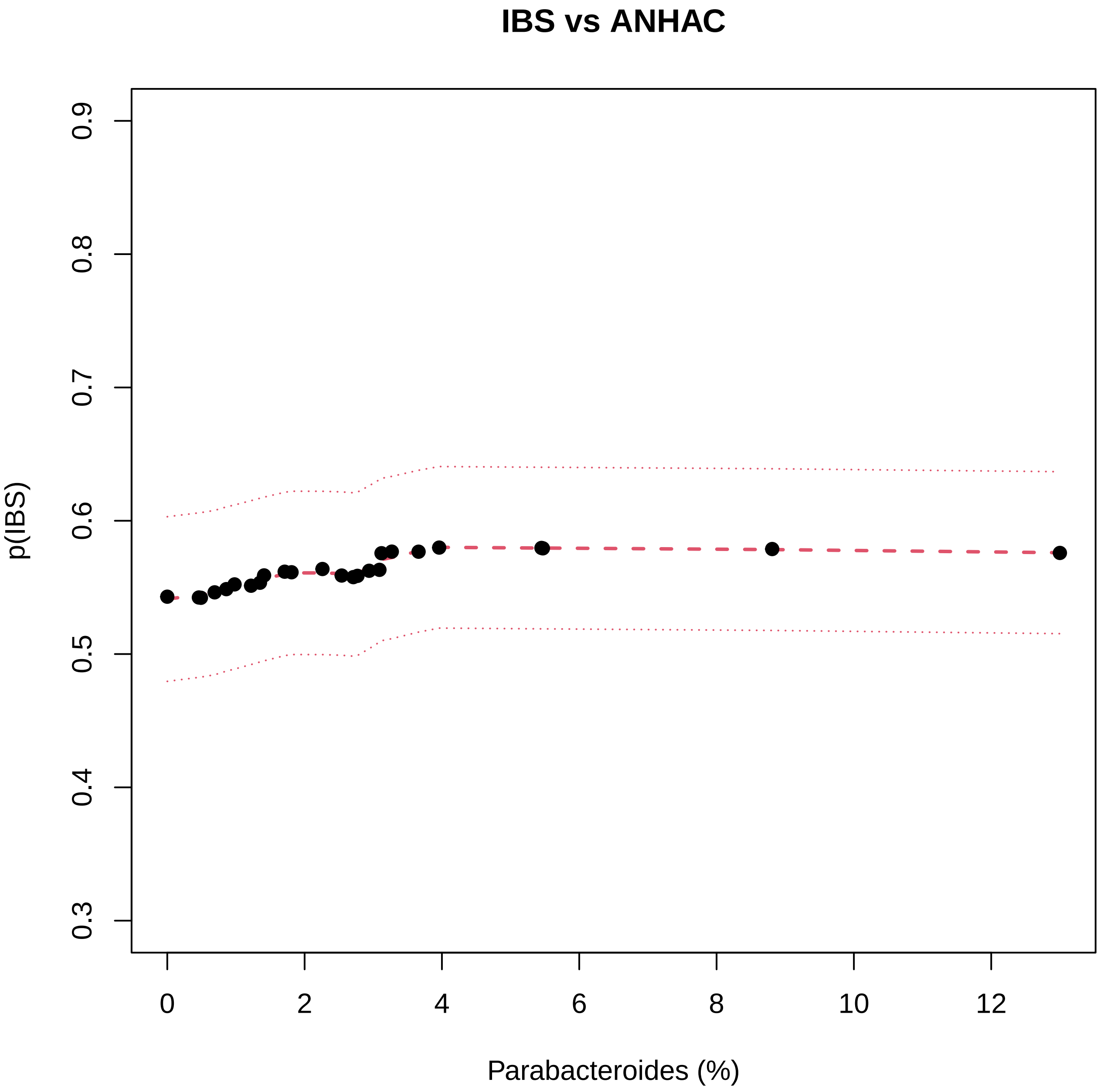

IBS vs ANHAC
p(IBS)
0.3
0.4
0.5
0.6
0.7
0.8
0.9
0
2
4
6
8
10
12
Parabacteroides (%)

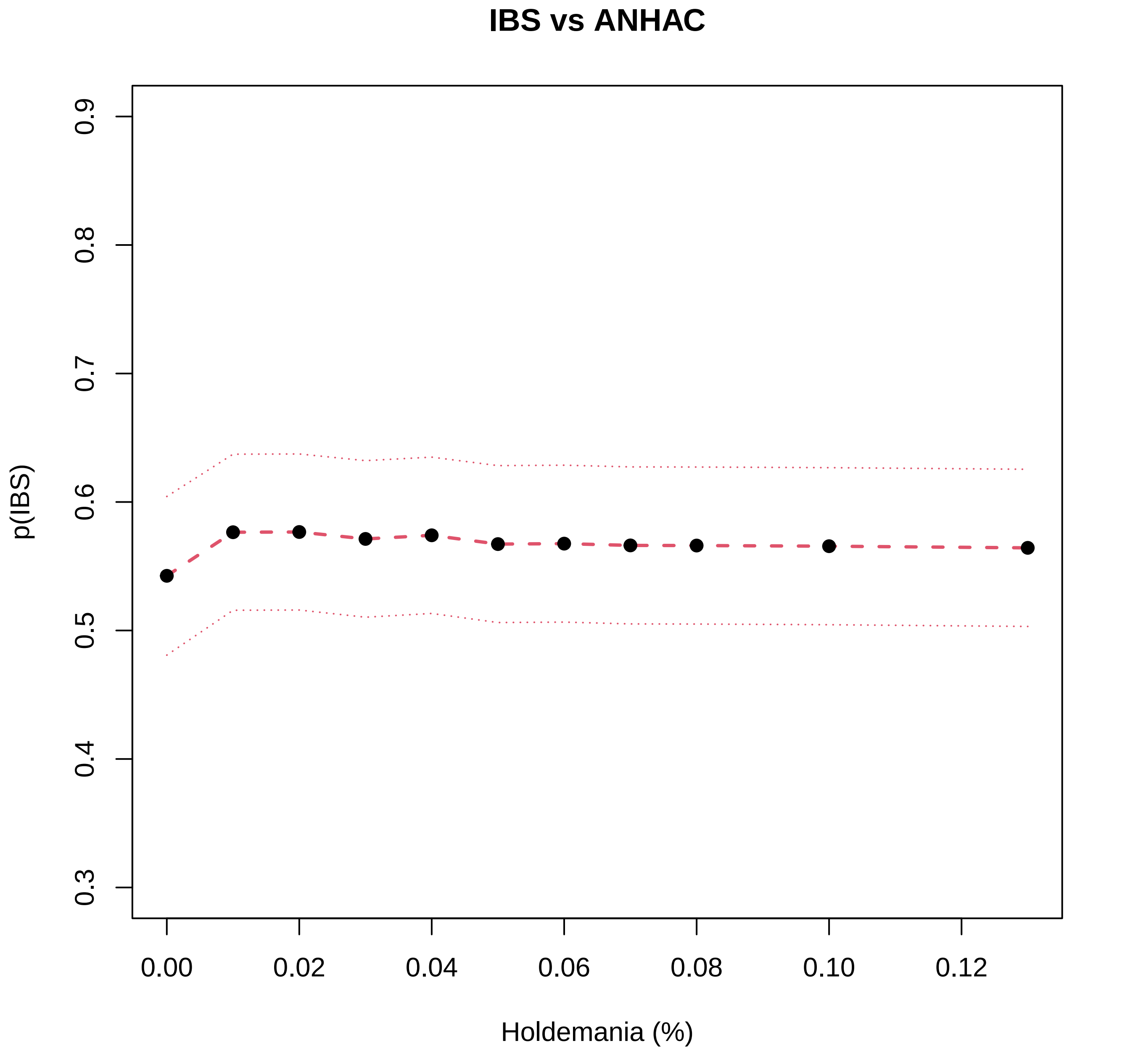
IBS vs ANHAC
p(IBS)
0.3
0.4
0.5
0.6
0.7
0.8
0.9
0.00
0.02
0.04
0.06
0.08
0.10
0.12
Holdemania (%)

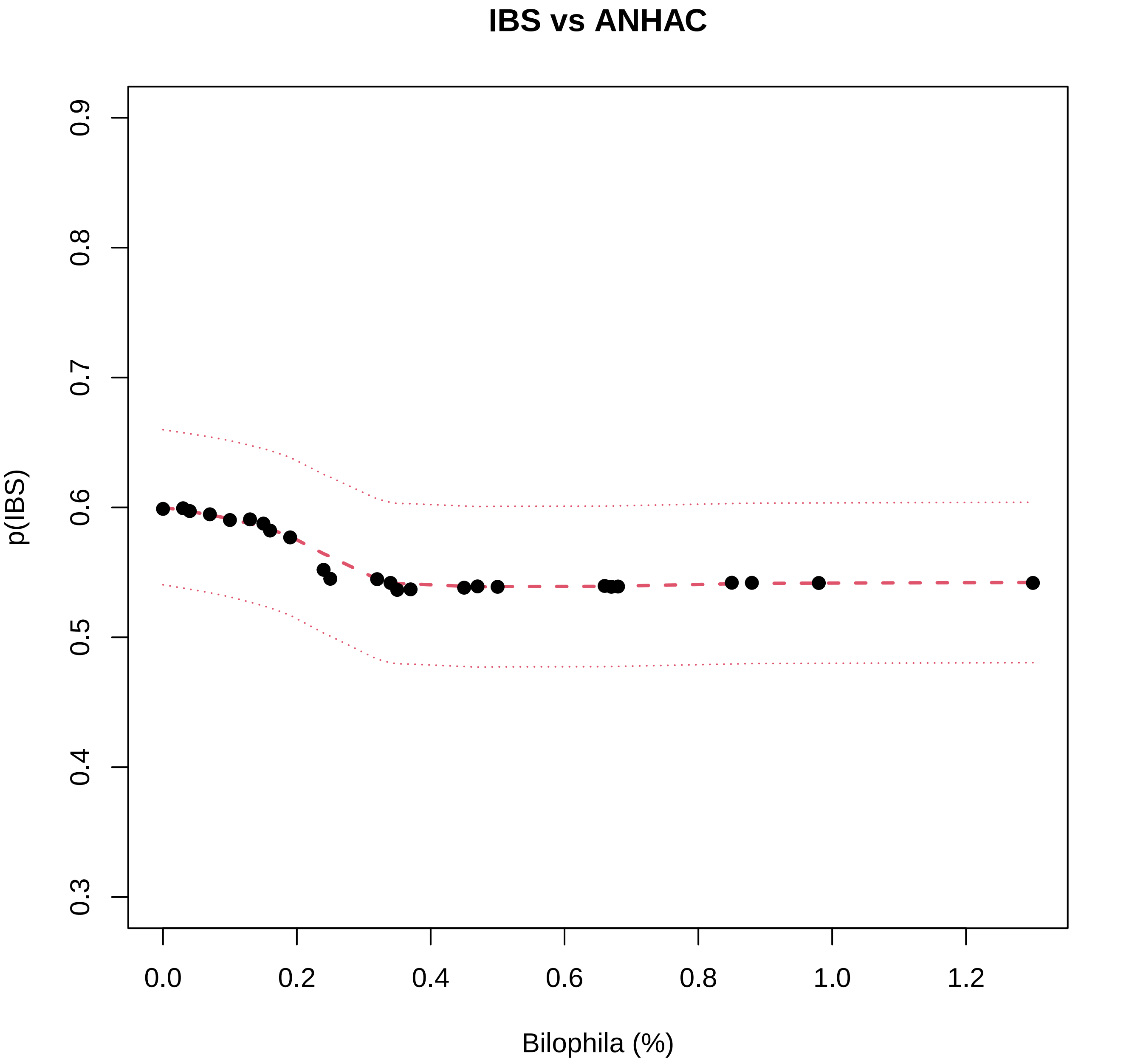
IBS vs ANHAC
p(IBS)
0.3
0.4
0.5
0.6
0.7
0.8
0.9
0.0
0.2
0.4
0.6
0.8
1.0
1.2
Bilophila (%)

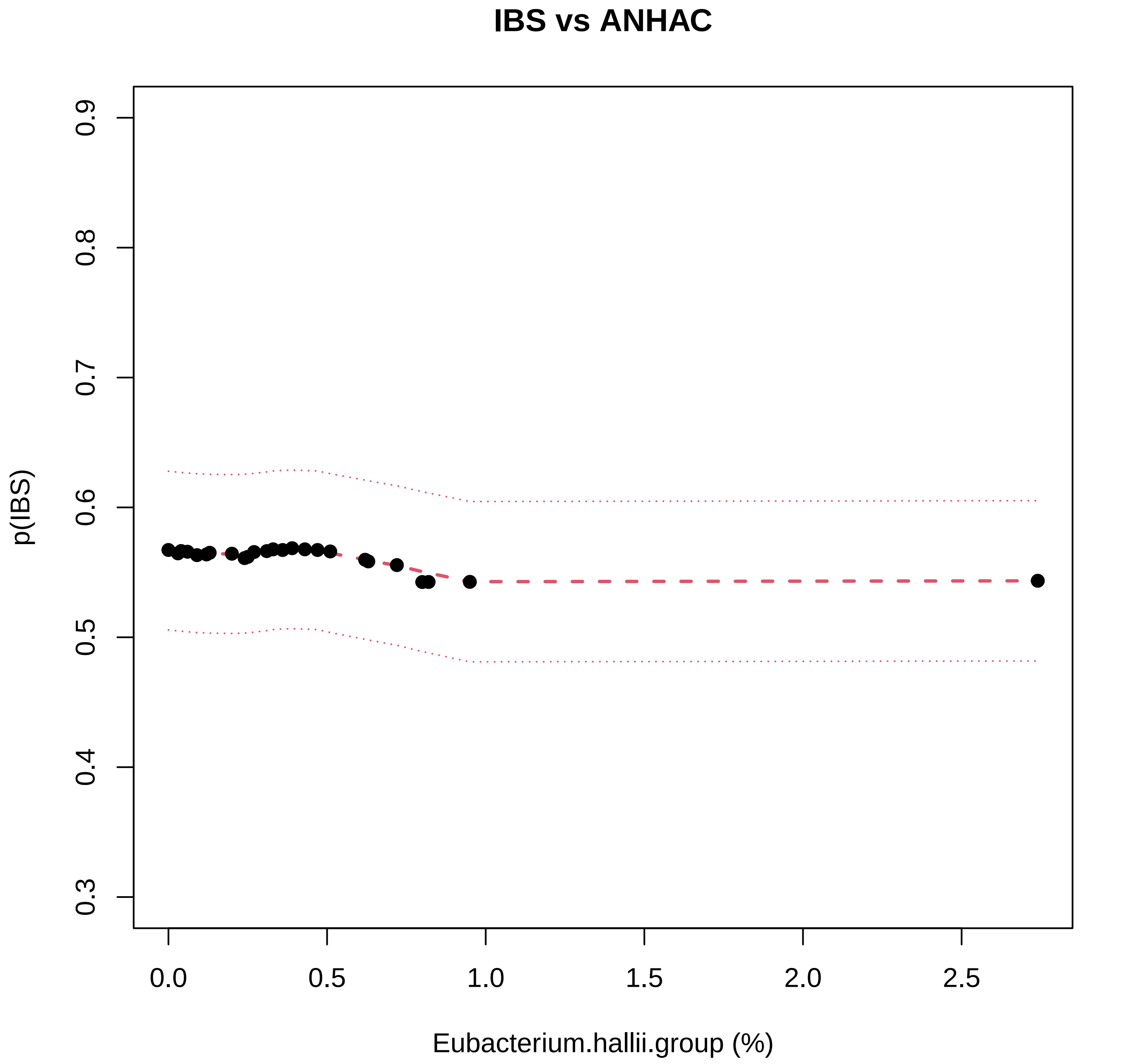
IBS vs ANHAC
p(IBS)
0.3
0.4
0.5
0.6
0.7
0.8
0.9
0.0
0.5
1.0
1.5
2.0
2.5
Eubacterium.hallii.group (%)

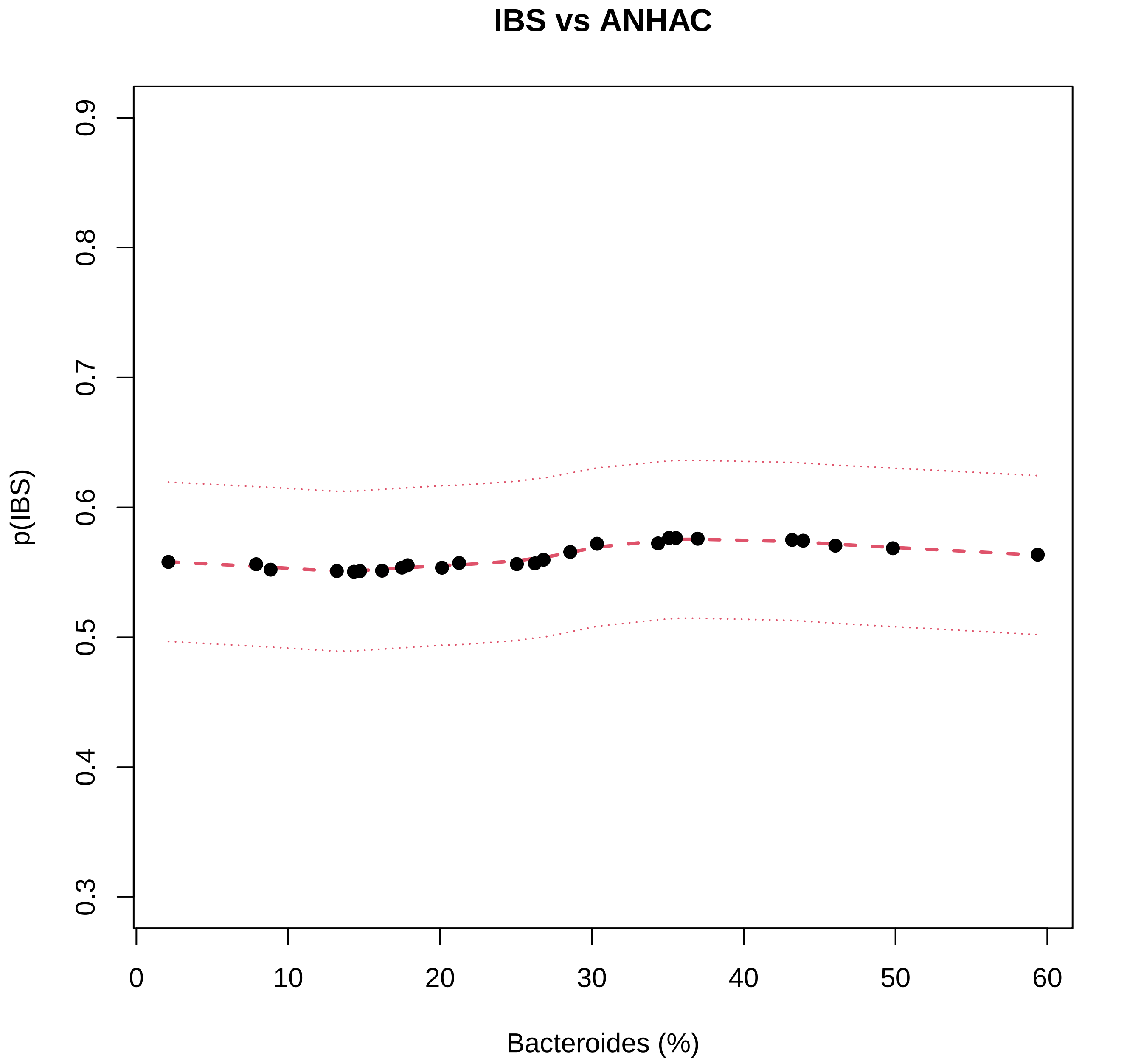
IBS vs ANHAC
p(IBS)
0.3
0.4
0.5
0.6
0.7
0.8
0.9
0
10
20
30
40
50
60
Bacteroides (%)

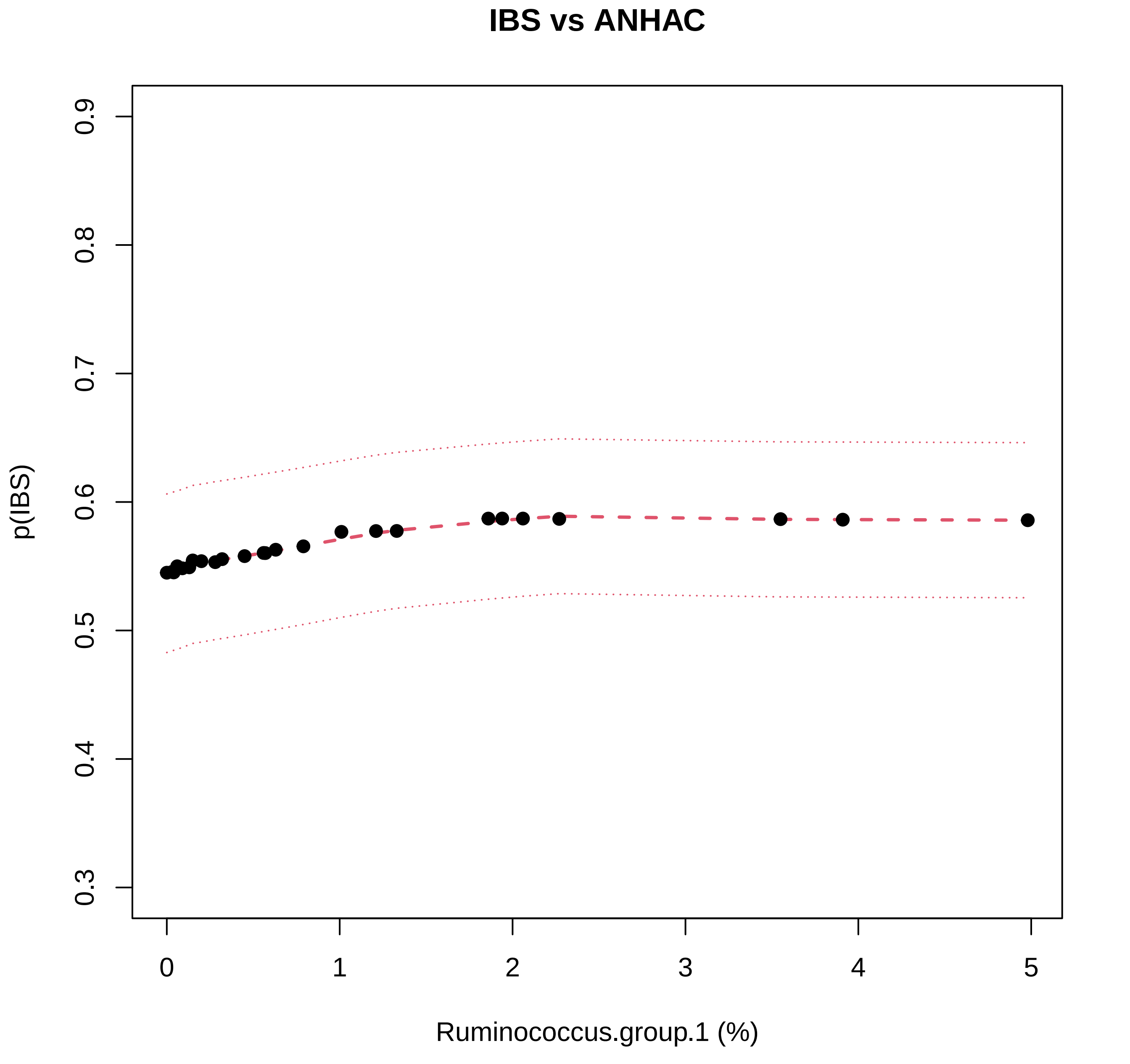

IBS vs ANHAC
p(IBS)
0.3
0.4
0.5
0.6
0.7
0.8
0.9
0
1
2
3
4
5
Ruminococcus.group.1 (%)

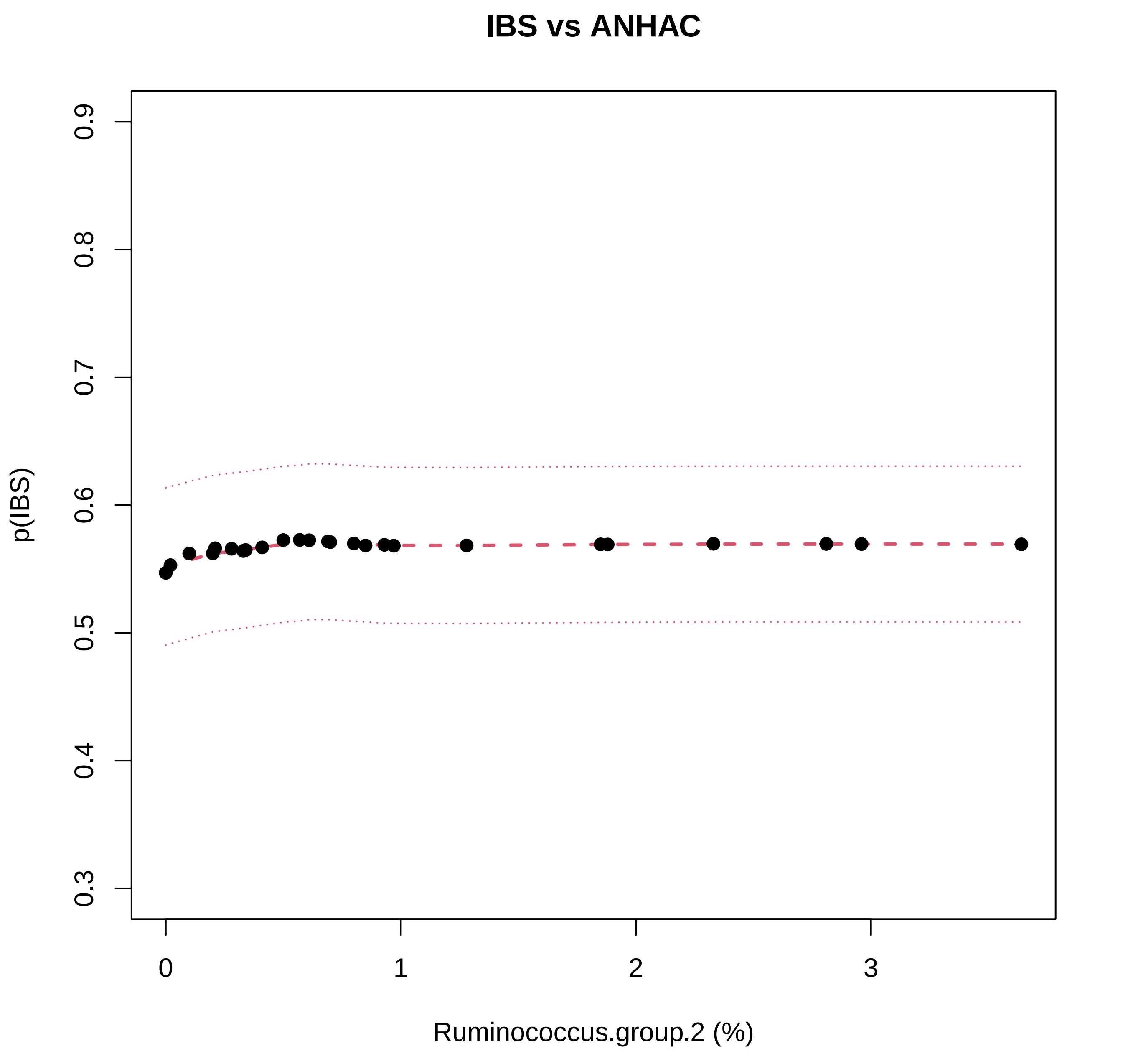

IBS vs ANHAC
p(IBS)
0.3
0.4
0.5
0.6
0.7
0.8
0.9
0
1
2
3
Ruminococcus.group.2 (%)

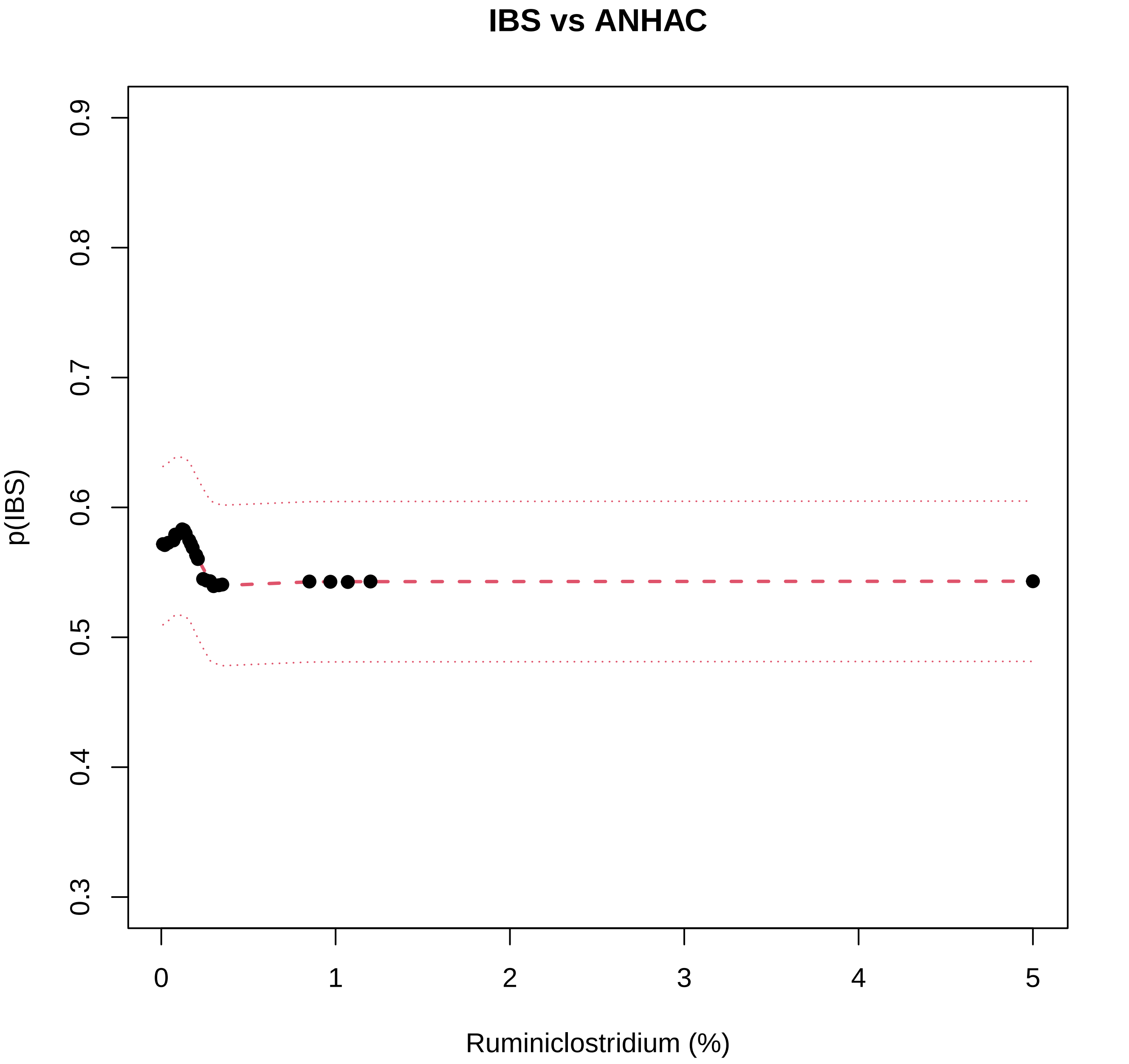

IBS vs ANHAC
p(IBS)
0.3
0.4
0.5
0.6
0.7
0.8
0.9
0
1
2
3
4
5
Ruminiclostridium (%)

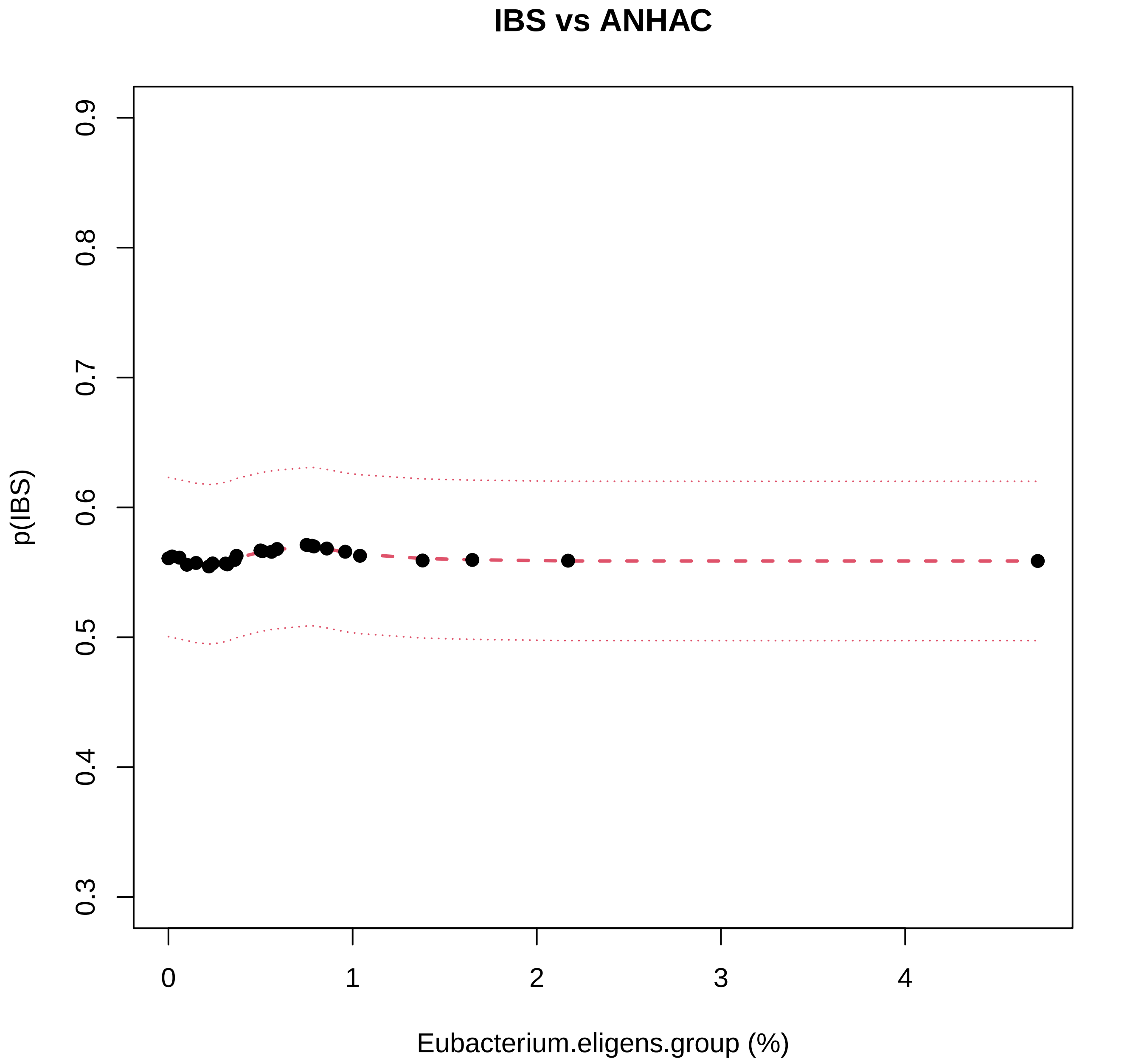

IBS vs ANHAC
p(IBS)
0.3
0.4
0.5
0.6
0.7
0.8
0.9
0
1
2
3
4
Eubacterium.eligens.group (%)

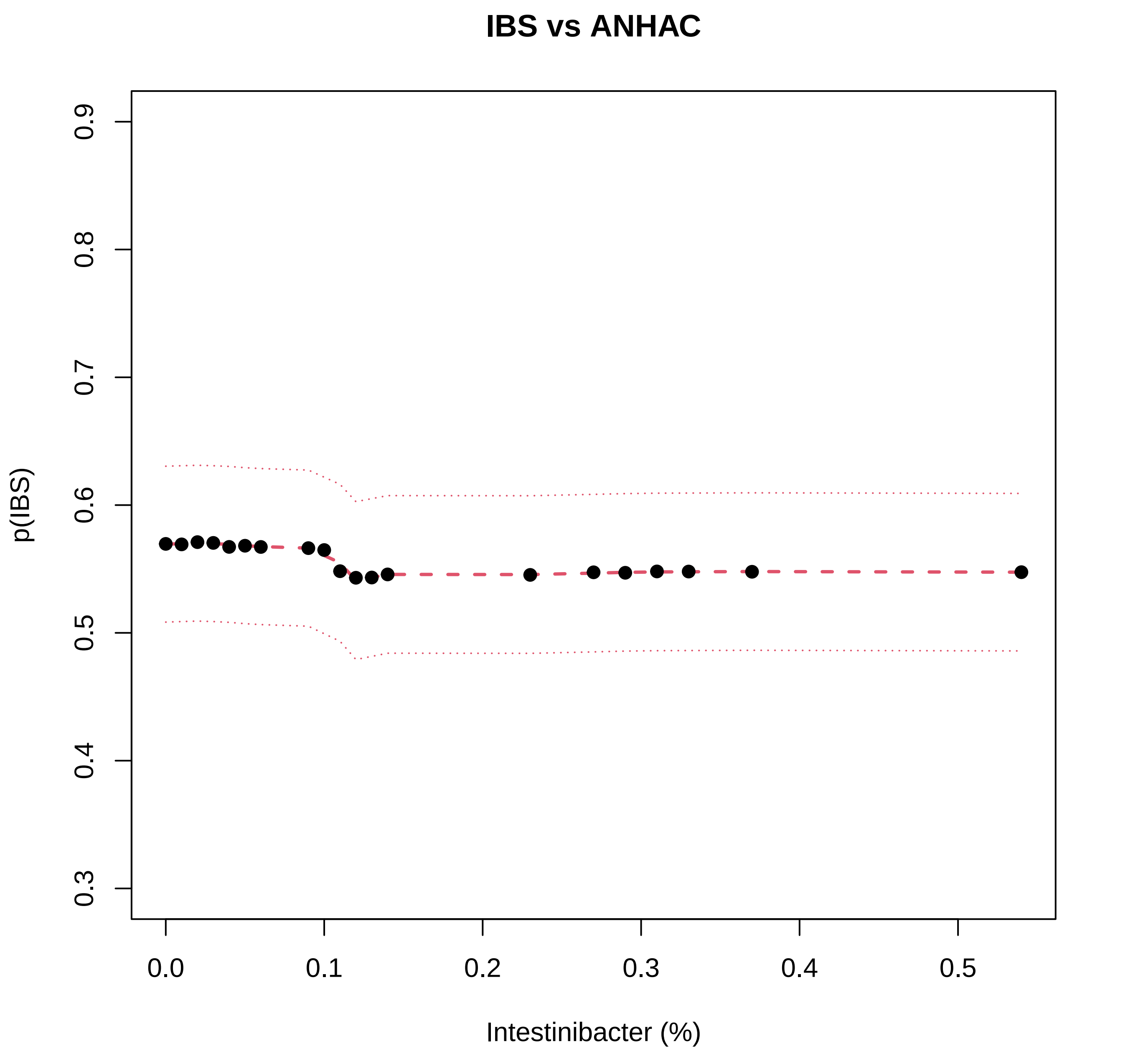
IBS vs ANHAC
p(IBS)
0.3
0.4
0.5
0.6
0.7
0.8
0.9
0.0
0.1
0.2
0.3
0.4
0.5
Intestinibacter (%)

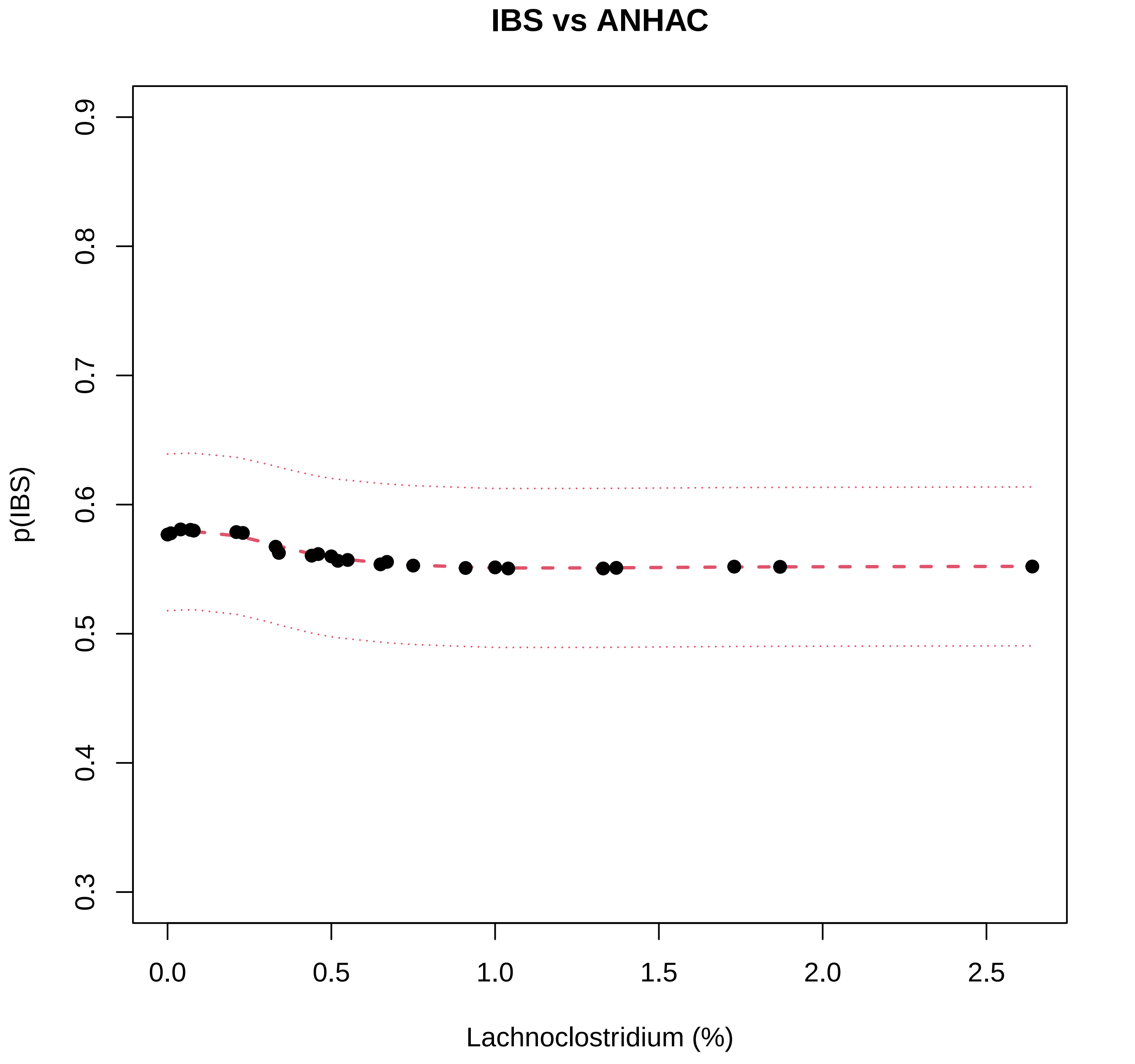
IBS vs ANHAC
p(IBS)
0.3
0.4
0.5
0.6
0.7
0.8
0.9
0.0
0.5
1.0
1.5
2.0
2.5
Lachnoclostridium (%)

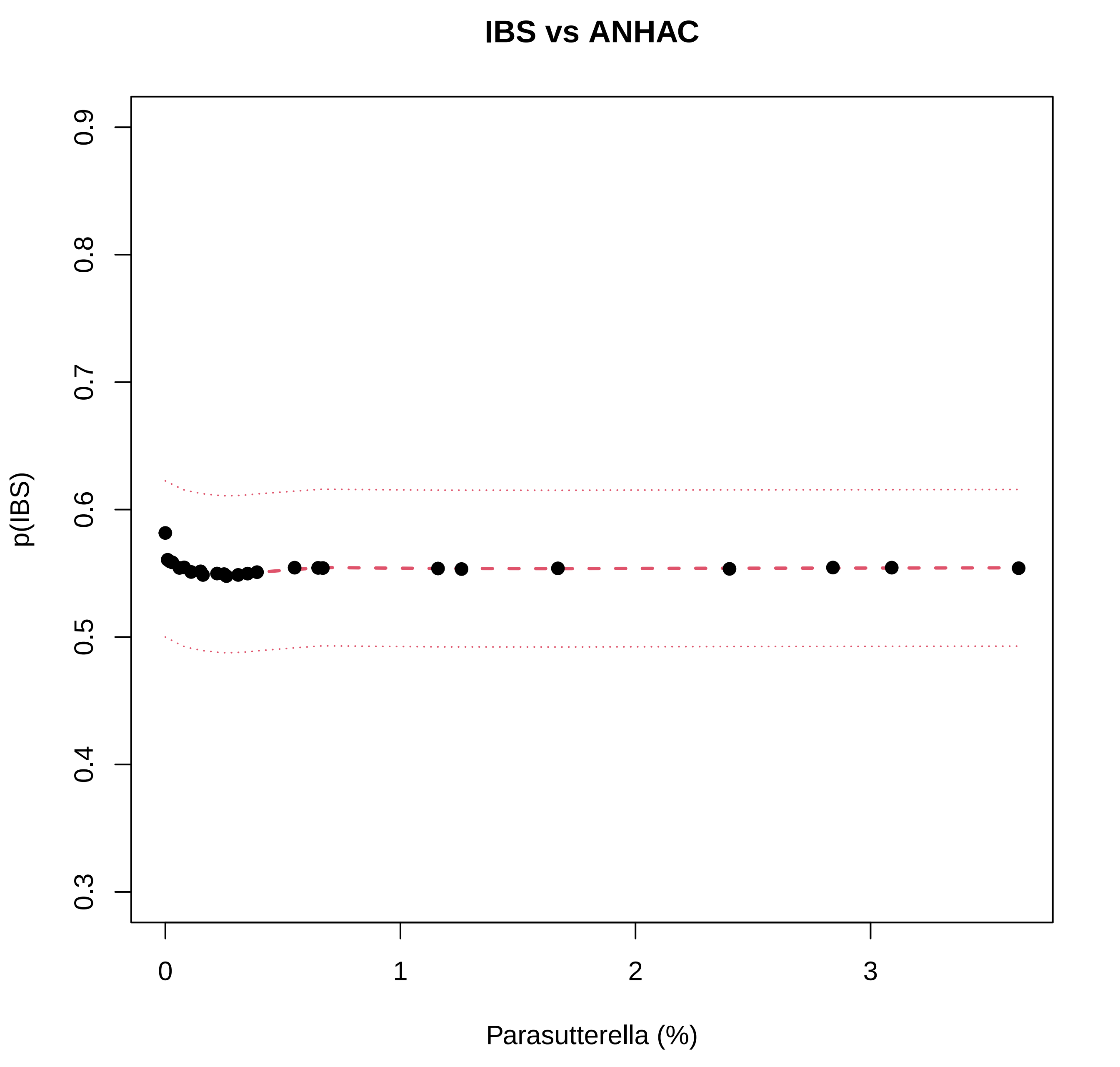
IBS vs ANHAC
p(IBS)
0.3
0.4
0.5
0.6
0.7
0.8
0.9
0
1
2
3
Parasutterella (%)

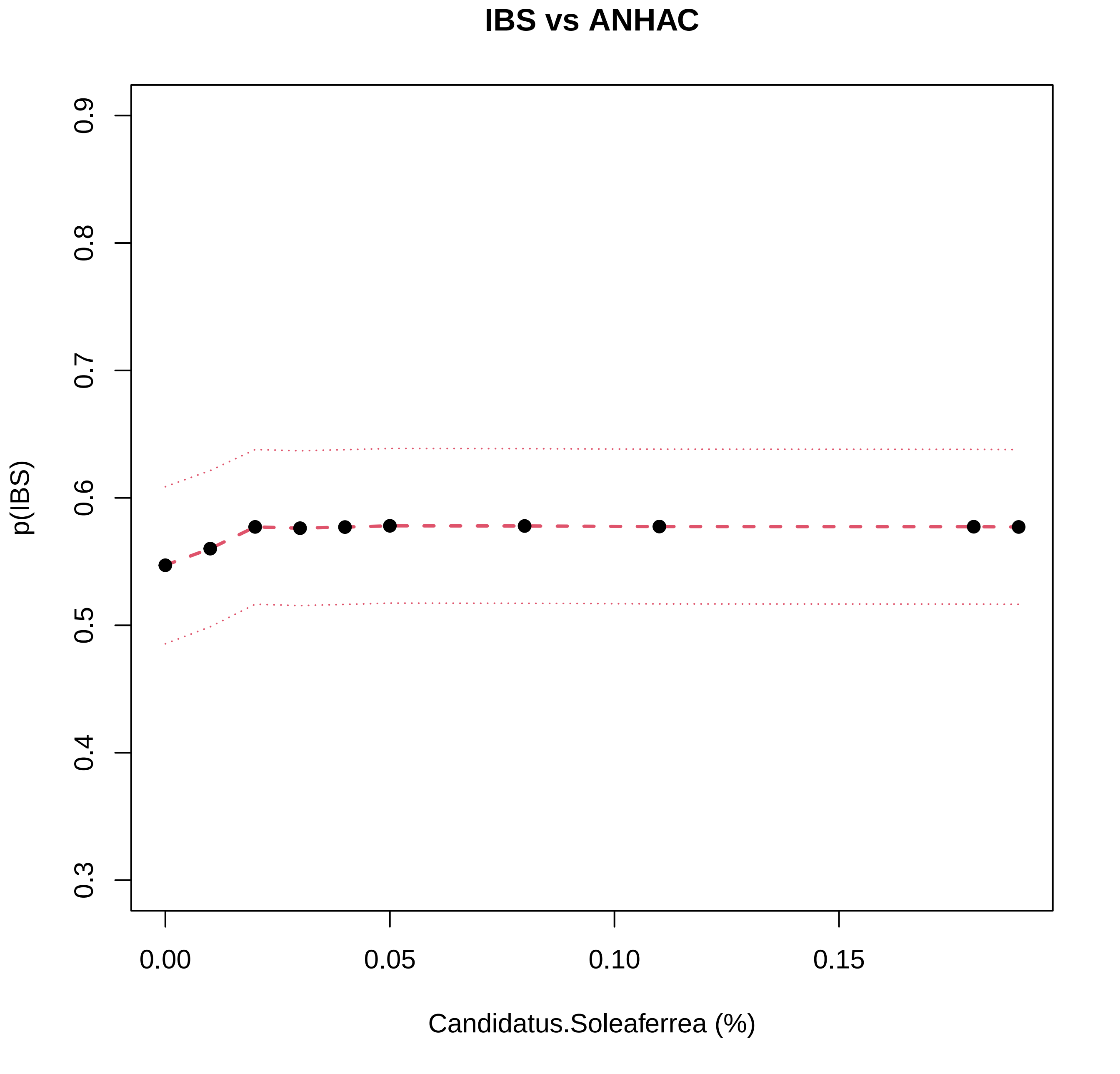
IBS vs ANHAC
p(IBS)
0.3
0.4
0.5
0.6
0.7
0.8
0.9
0.00
0.05
0.10
0.15
Candidatus.Soleaferrea (%)

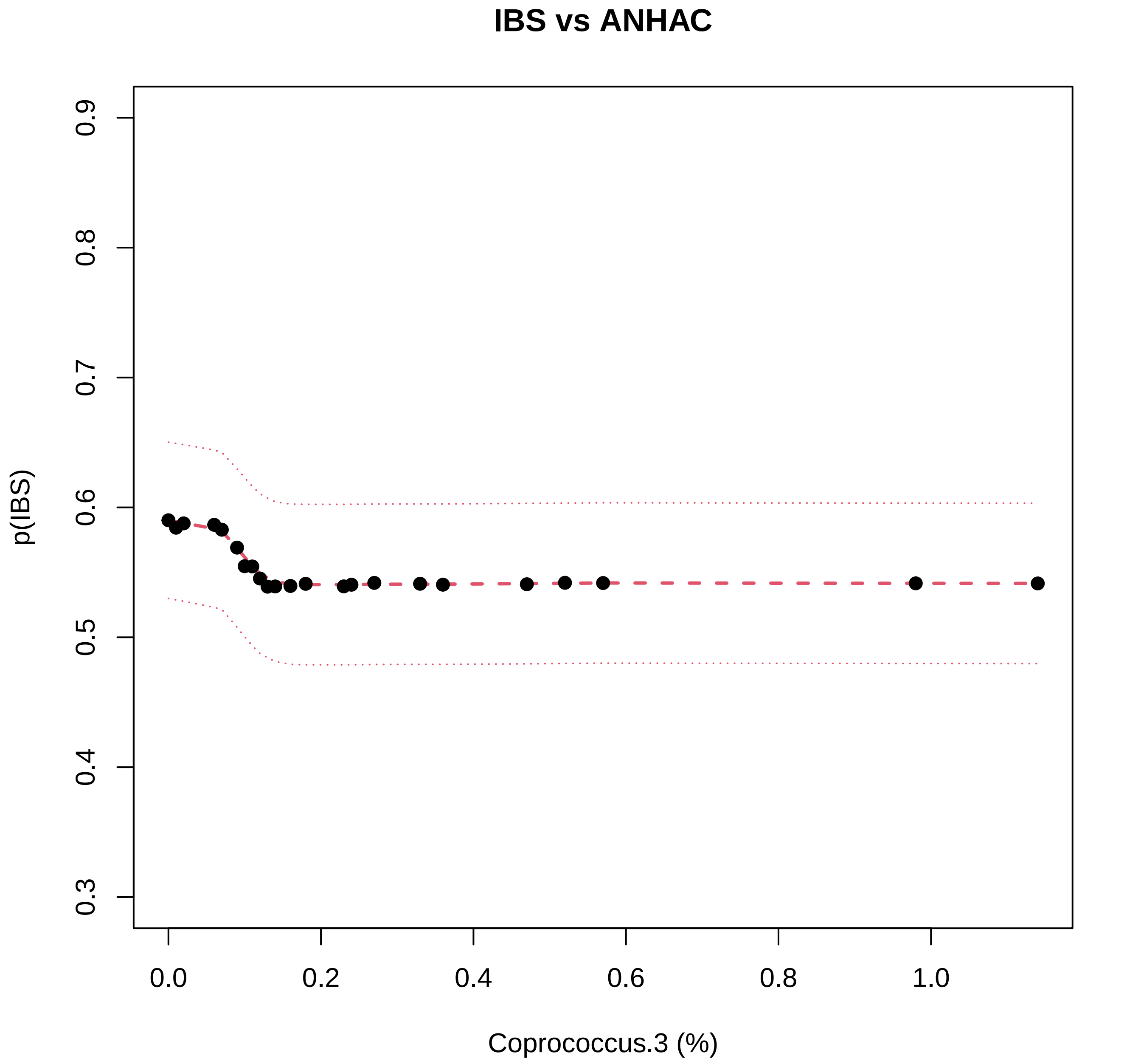

IBS vs ANHAC
p(IBS)
0.3
0.4
0.5
0.6
0.7
0.8
0.9
0.0
0.2
0.4
0.6
0.8
1.0
Coprococcus.3 (%)

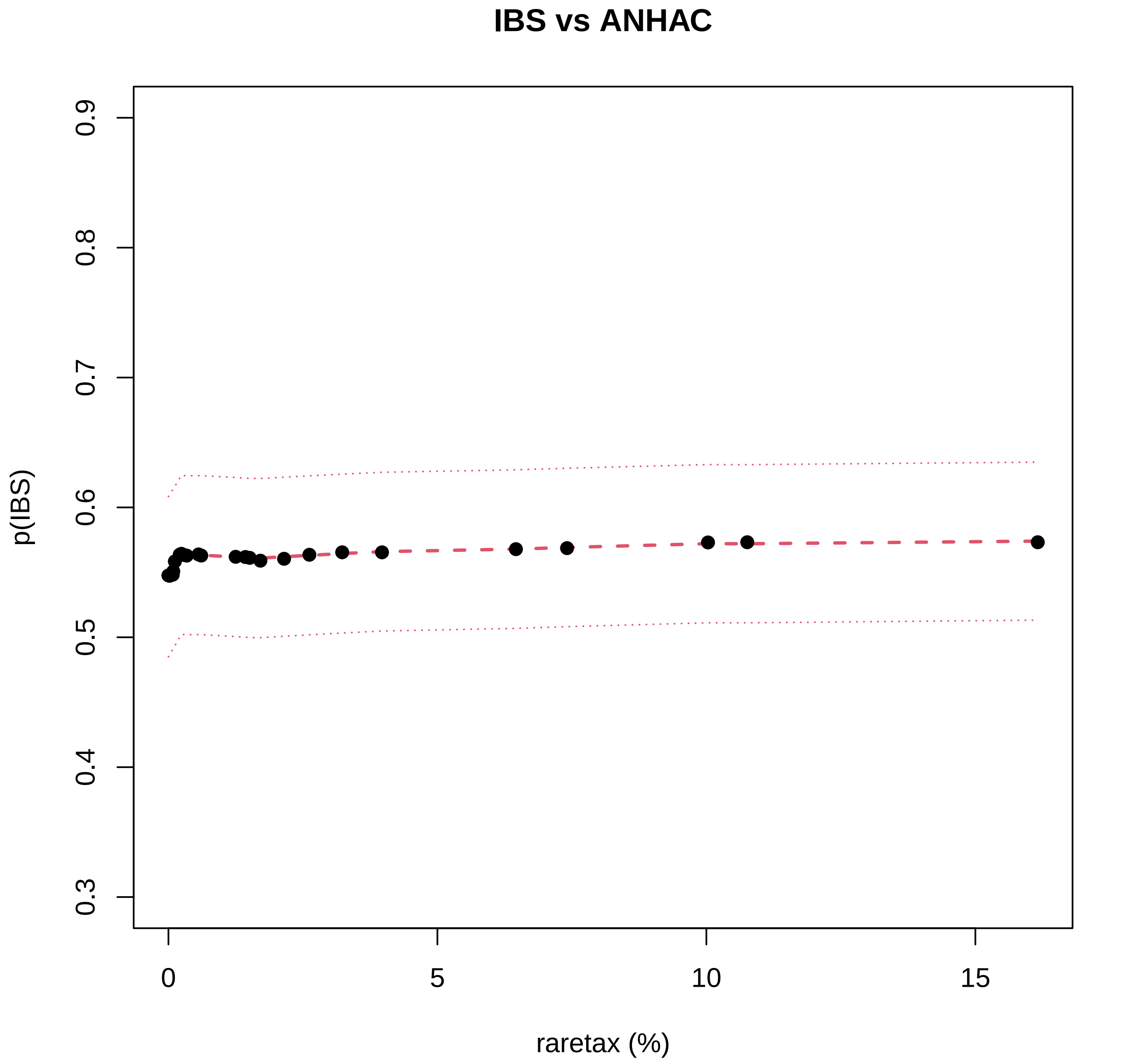

IBS vs ANHAC
p(IBS)
0.3
0.4
0.5
0.6
0.7
0.8
0.9
0
5
10
15
raretax (%)

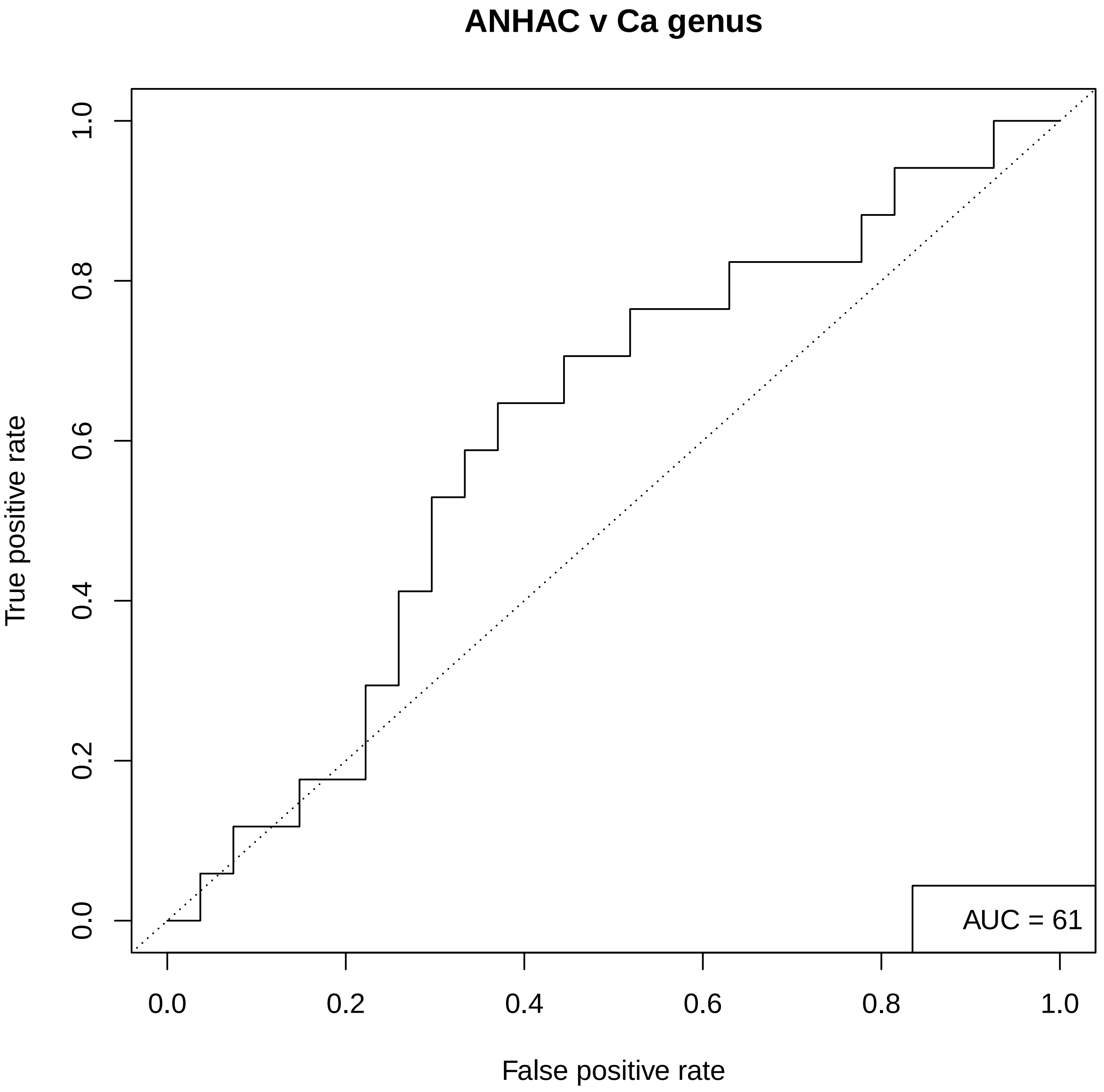
ANHAC v Ca genus
True positive rate
False positive rate
0.0
0.2
0.4
0.6
0.8
1.0
AUC = 61

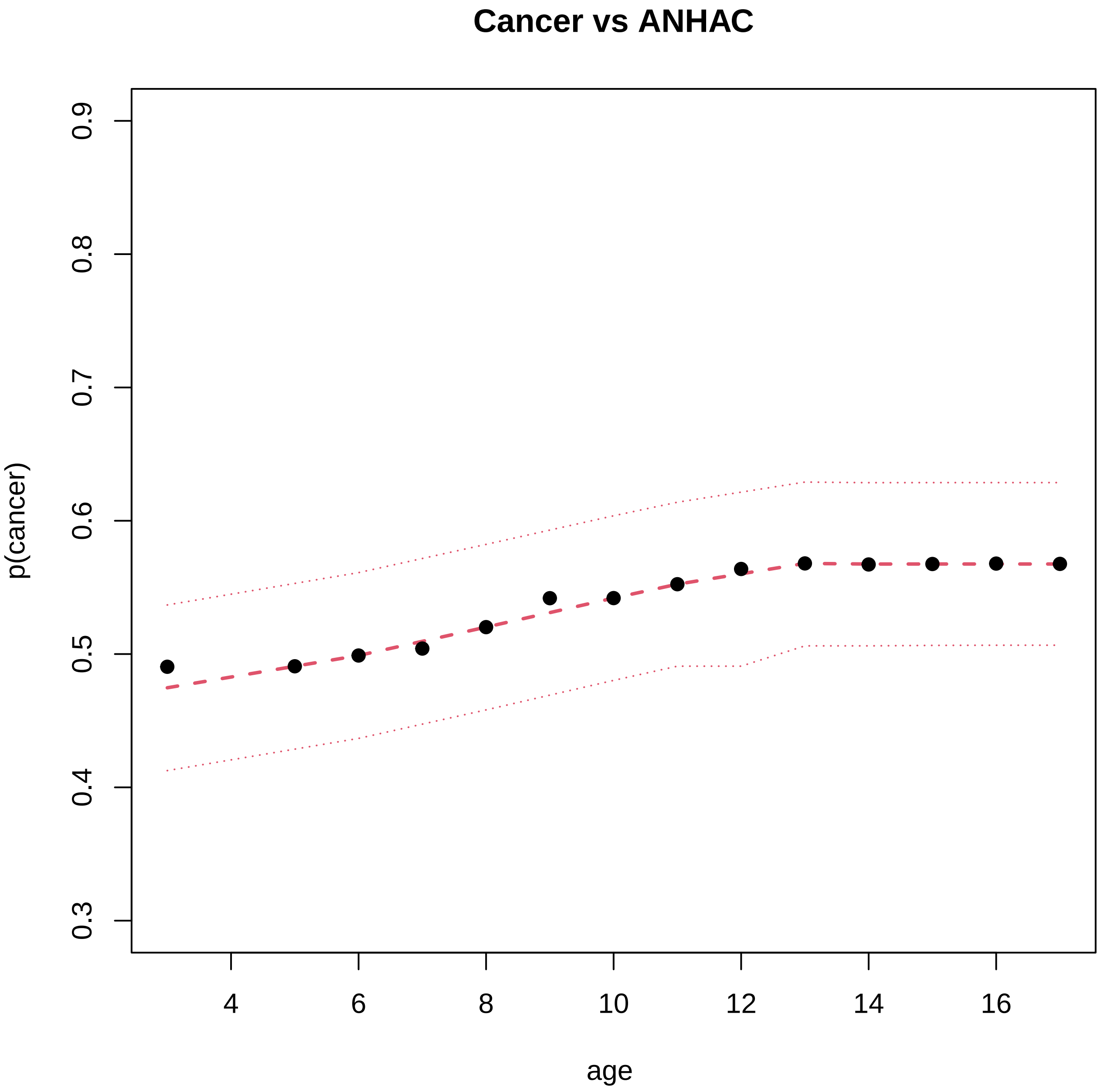

Cancer vs ANHAC
p(cancer)
0.3
0.4
0.5
0.6
0.7
0.8
0.9
4
6
8
10
12
14
16
age

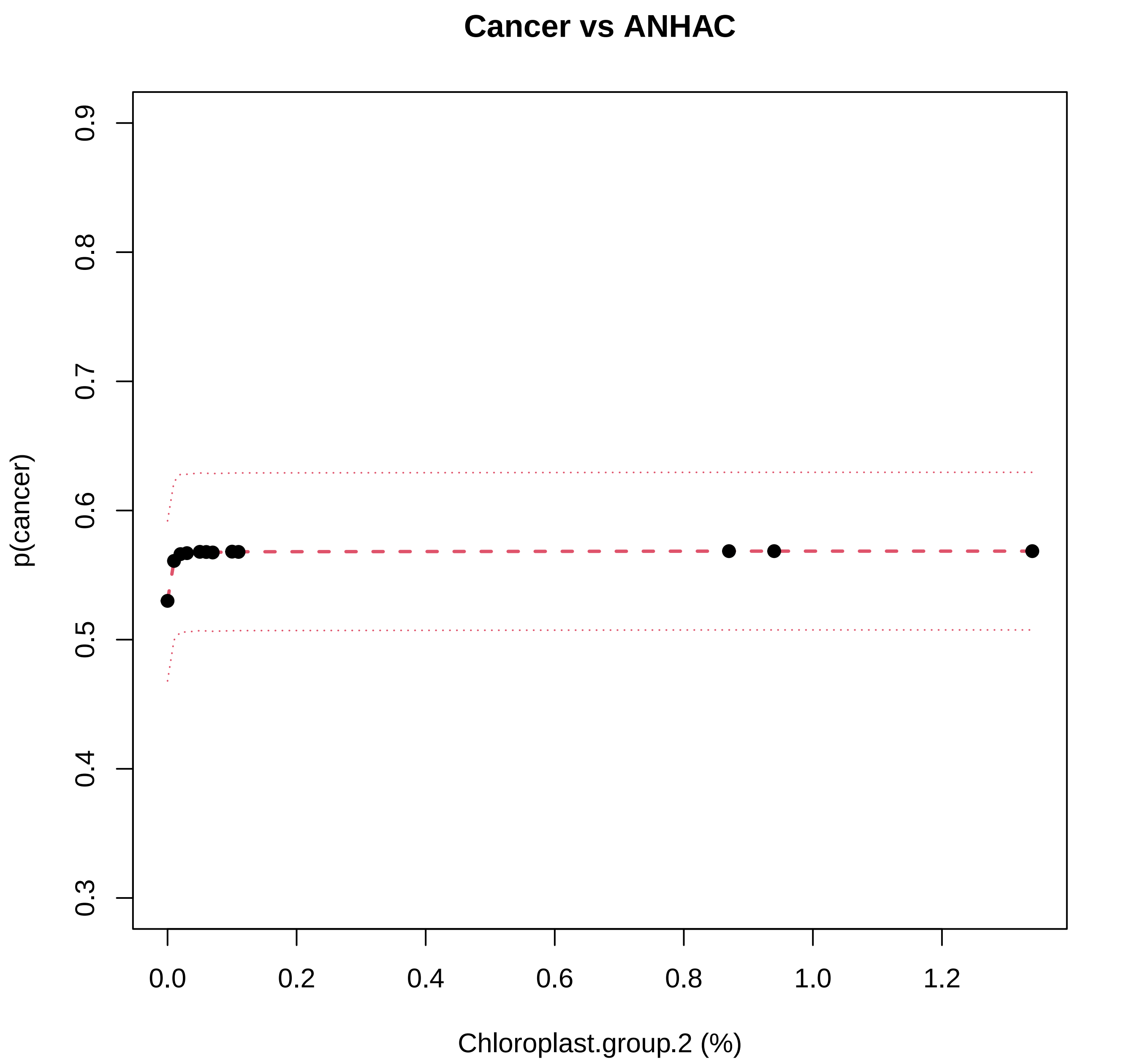
Cancer vs ANHAC
p(cancer)
0.3
0.4
0.5
0.6
0.7
0.8
0.9
0.0
0.2
0.4
0.6
0.8
1.0
1.2
Chloroplast.group.2 (%)

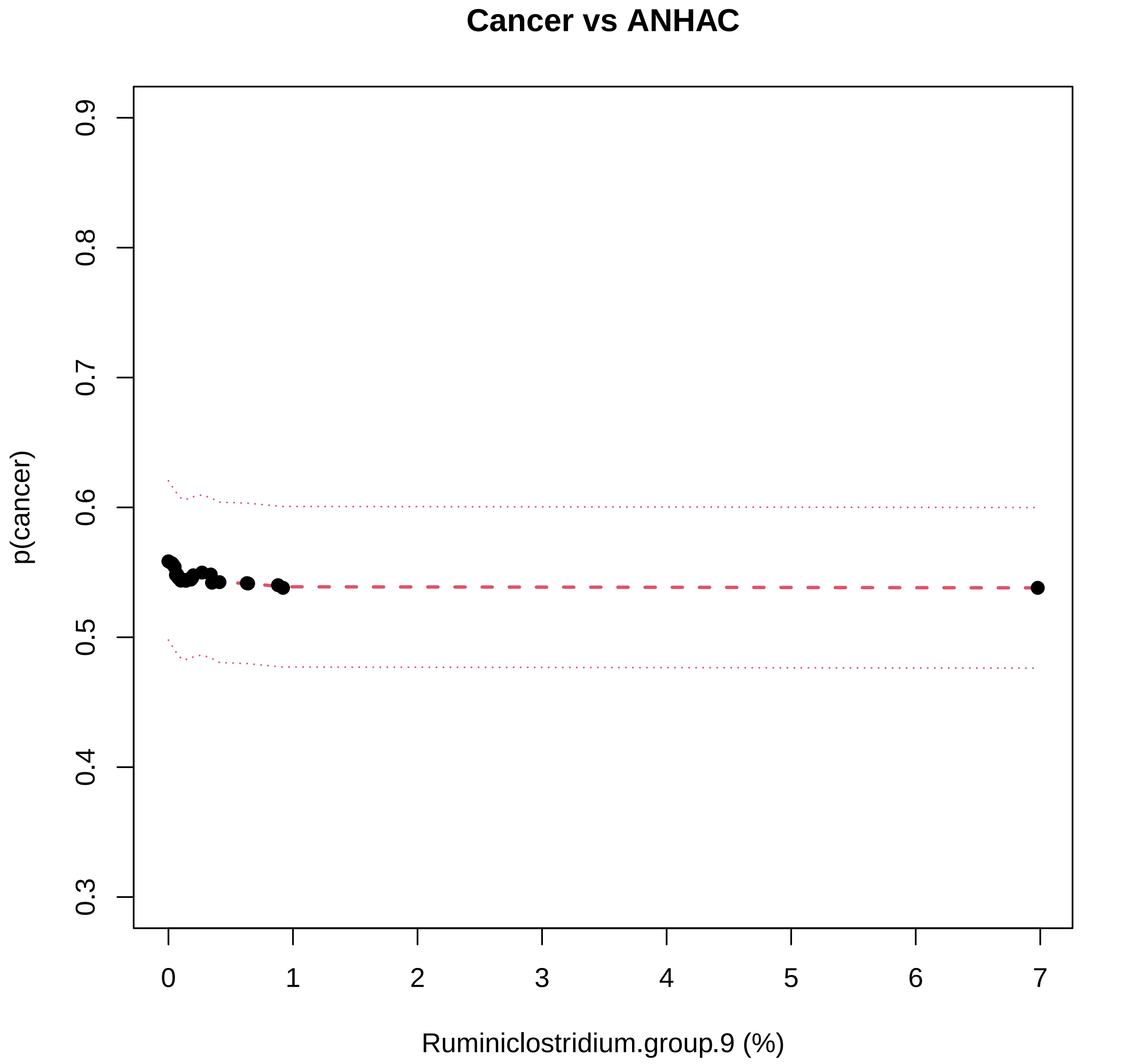

Cancer vs ANHAC
p(cancer)
0.3
0.4
0.5
0.6
0.7
0.8
0.9
0
1
2
3
4
5
6
7
Ruminiclostridium.group.9 (%)

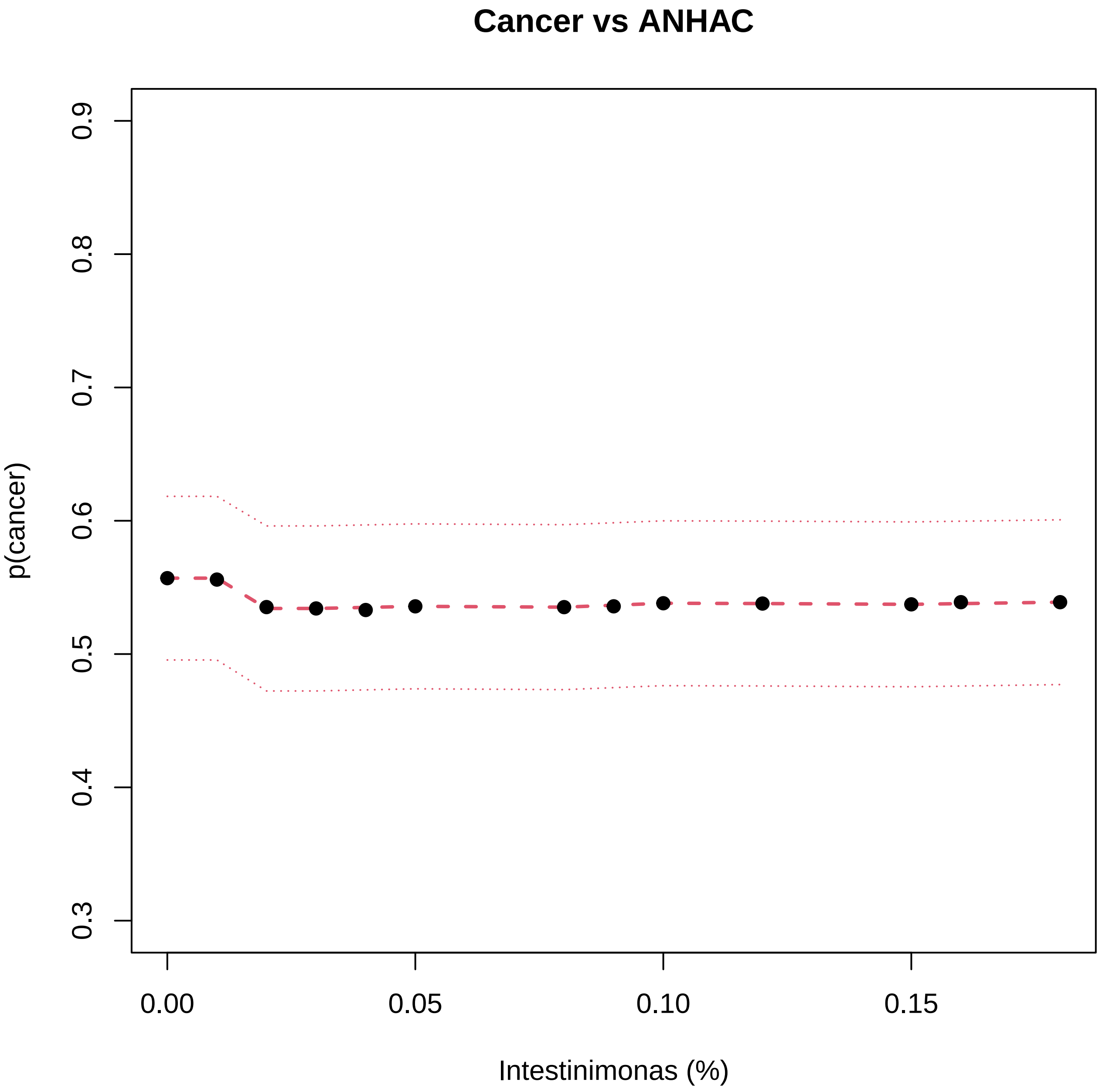

Cancer vs ANHAC
p(cancer)
0.3
0.4
0.5
0.6
0.7
0.8
0.9
0.00
0.05
0.10
0.15
Intestinimonas (%)

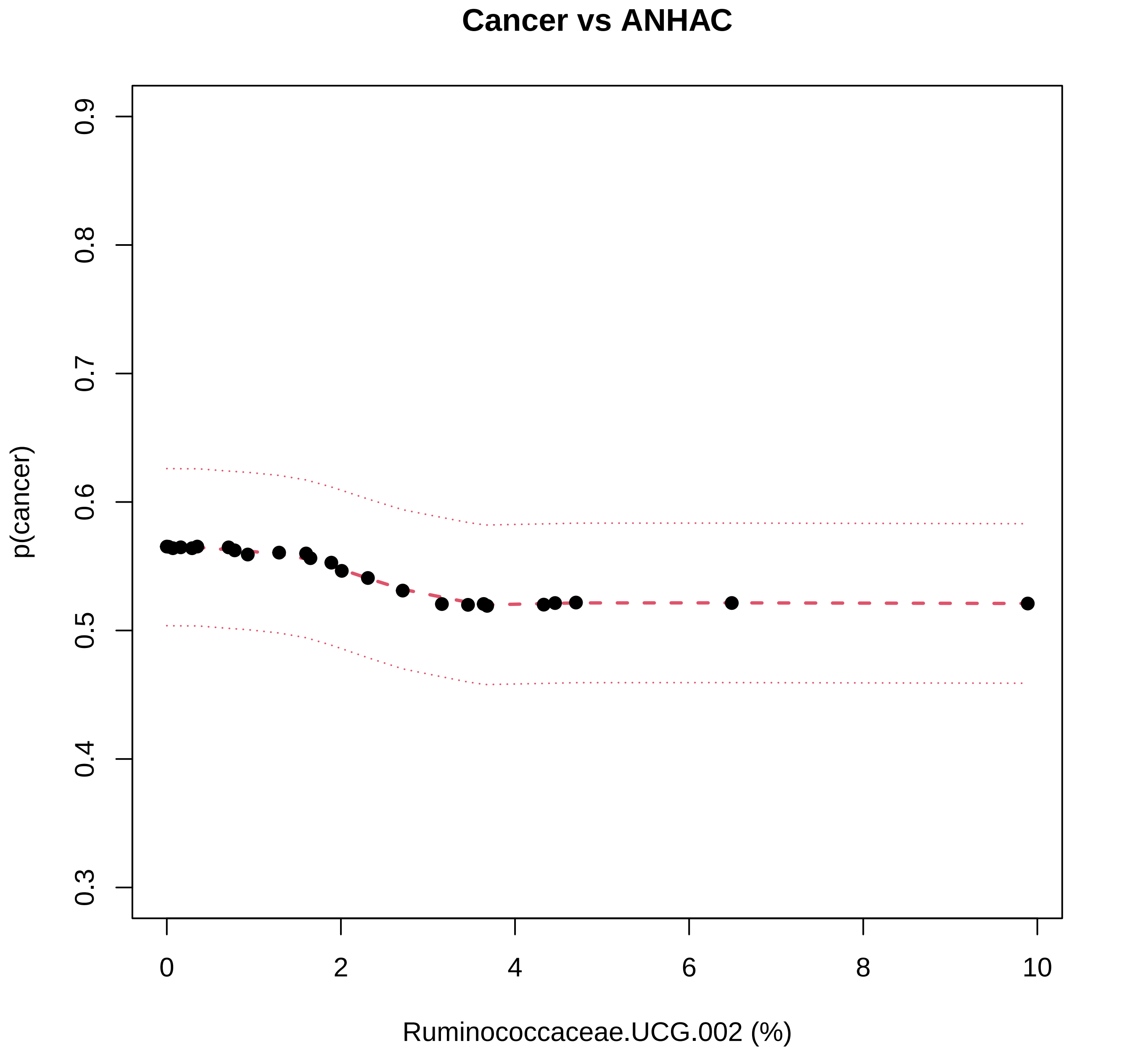

Cancer vs ANHAC
p(cancer)
0.3
0.4
0.5
0.6
0.7
0.8
0.9
0
2
4
6
8
10
Ruminococcaceae.UCG.002 (%)

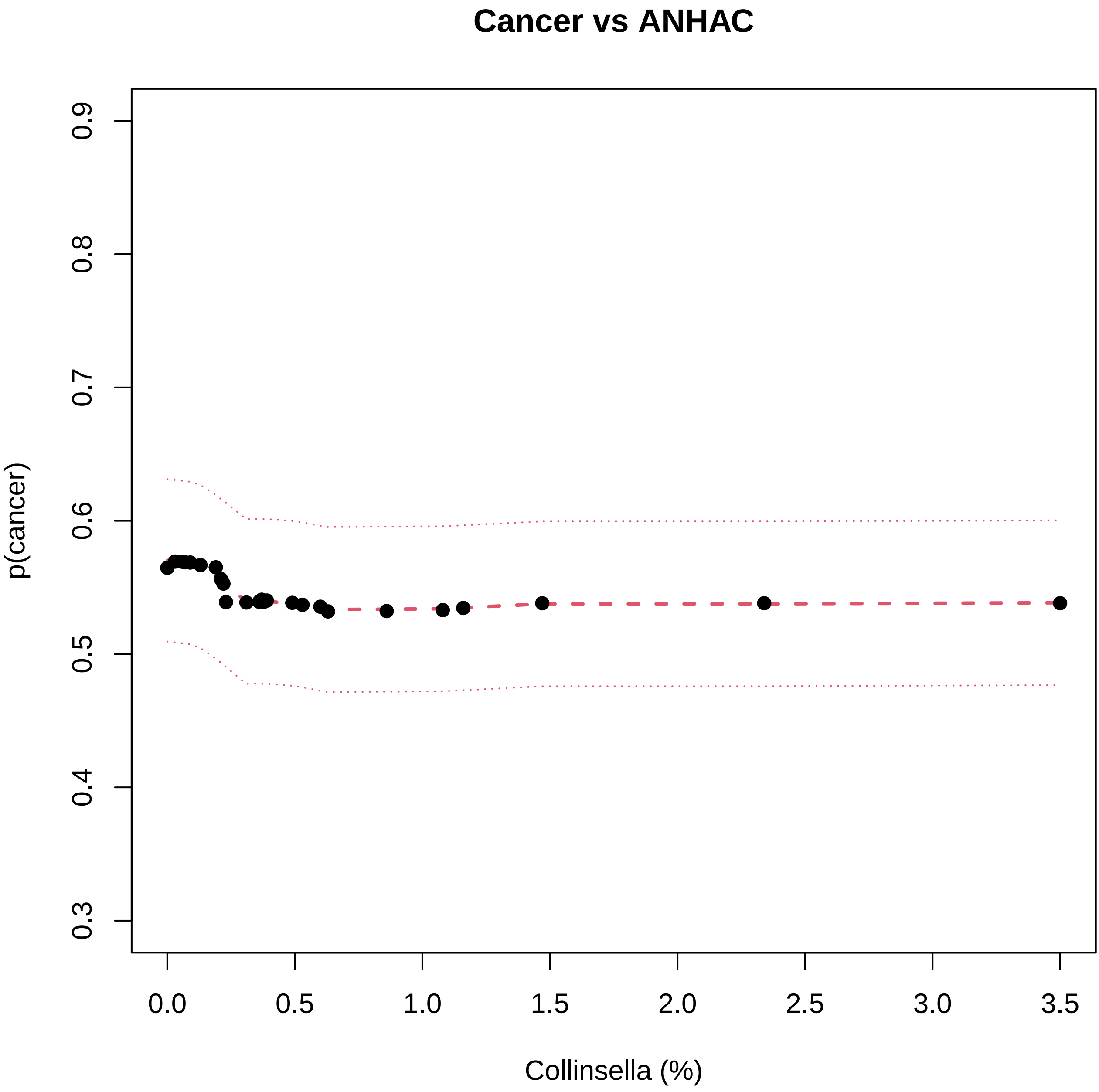
Cancer vs ANHAC
p(cancer)
0.3
0.4
0.5
0.6
0.7
0.8
0.9
0.0
0.5
1.0
1.5
2.0
2.5
3.0
3.5
Collinsella (%)

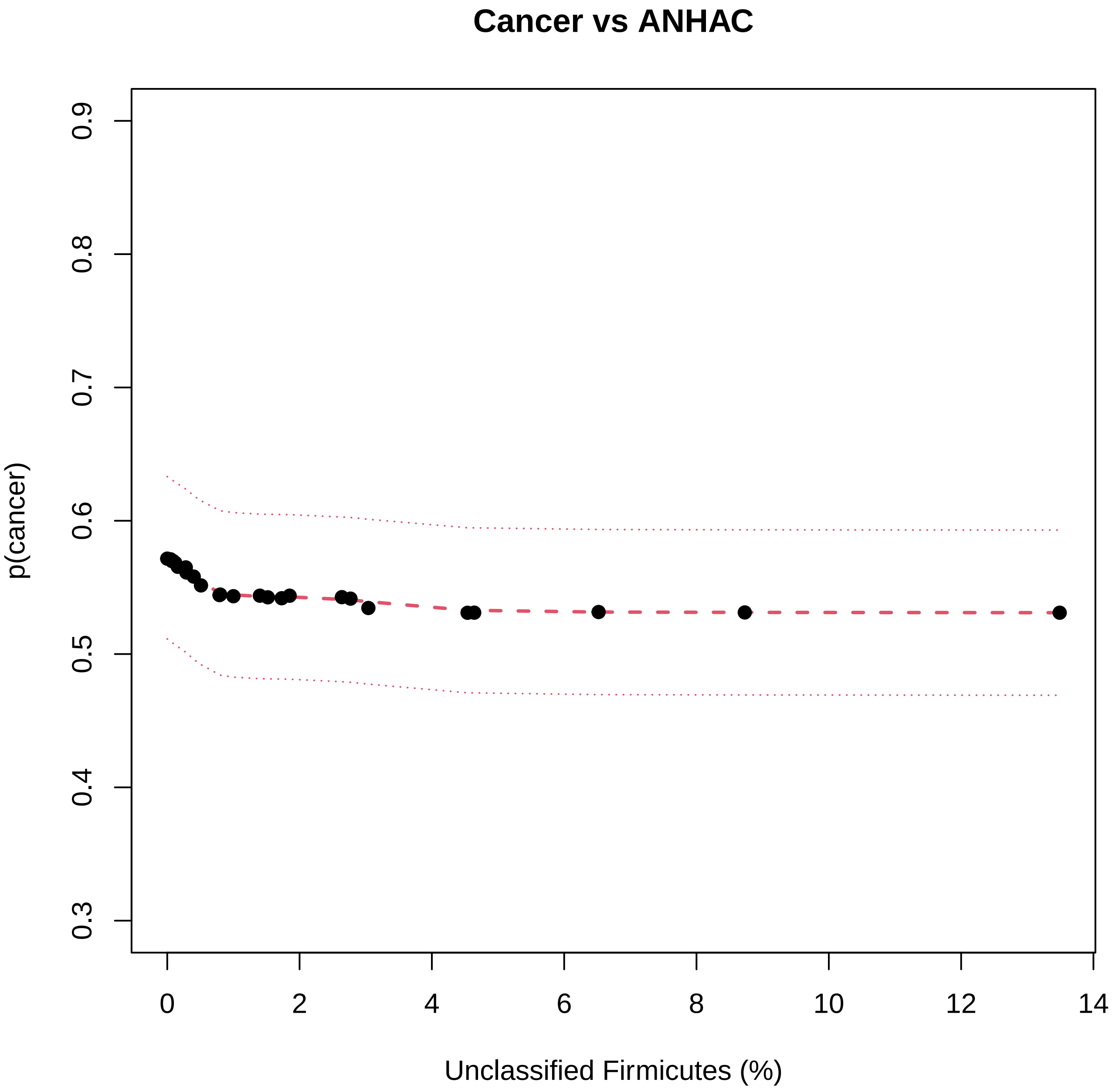
Cancer vs ANHAC
p(cancer)
0.3
0.4
0.5
0.6
0.7
0.8
0.9
0
2
4
6
8
10
12
14
Unclassified Firmicutes (%)

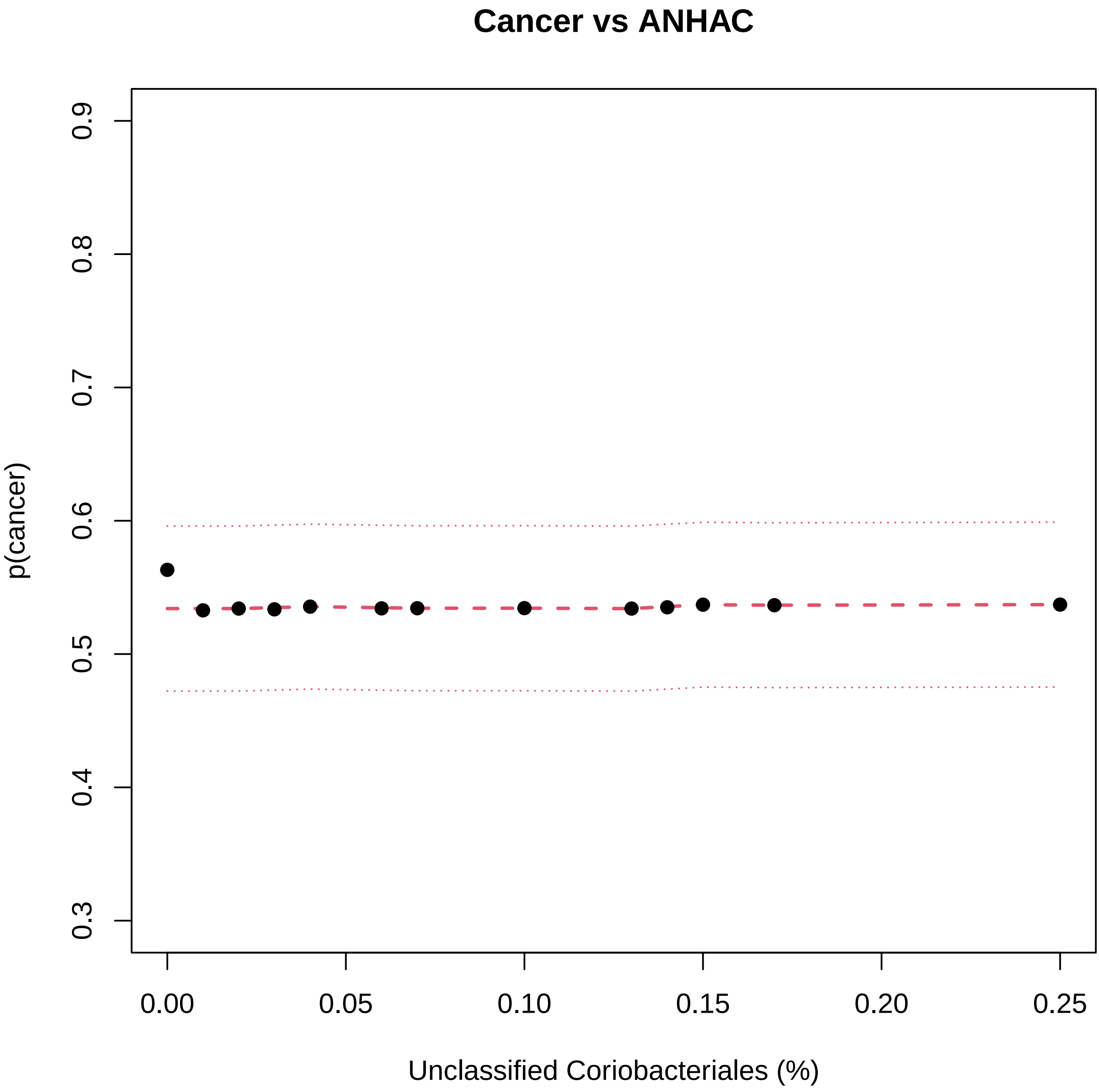

Cancer vs ANHAC
p(cancer)
0.3
0.4
0.5
0.6
0.7
0.8
0.9
0.00
0.05
0.10
0.15
0.20
0.25
Unclassified Coriobacteriales (%)

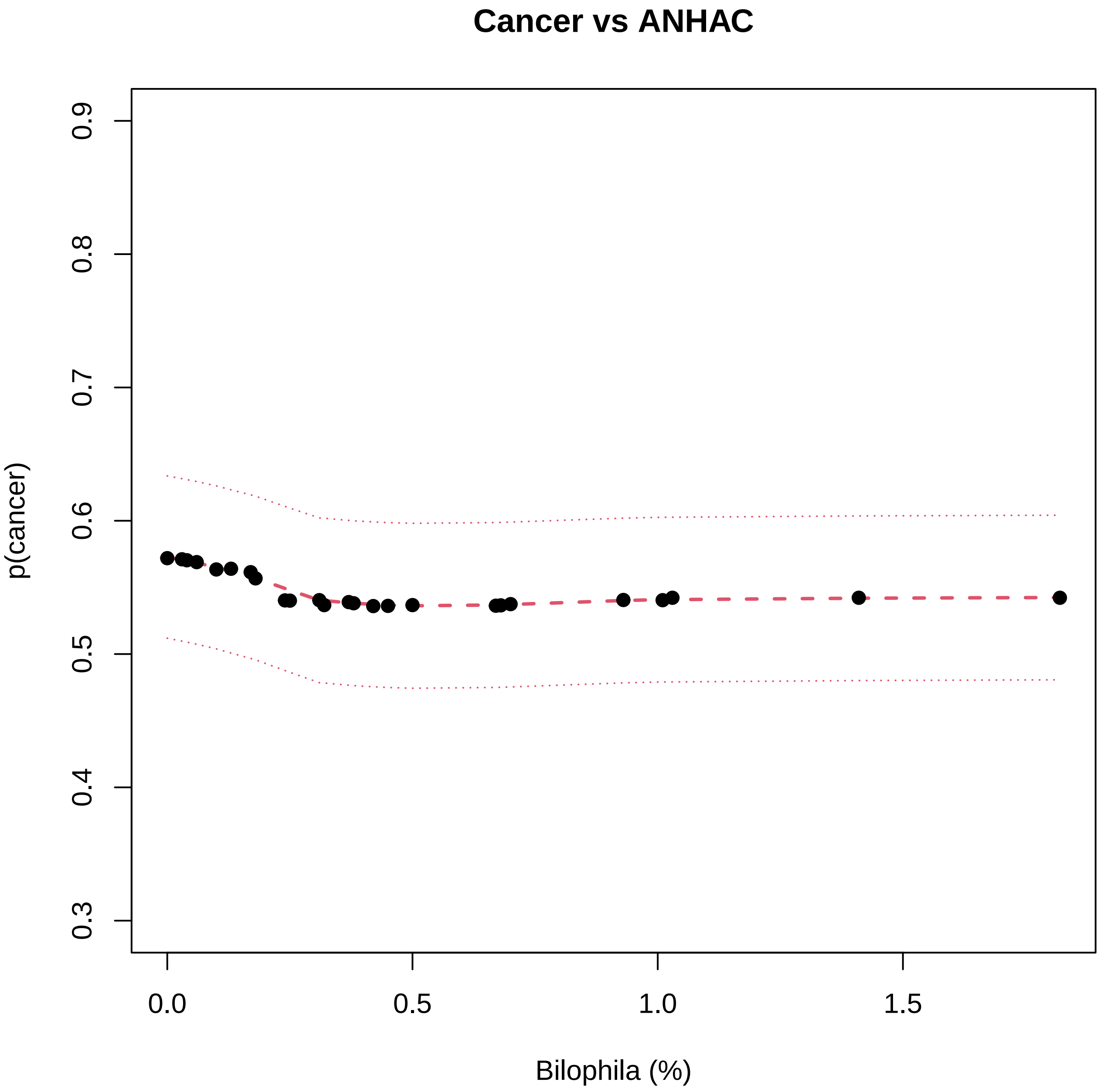

Cancer vs ANHAC
p(cancer)
0.3
0.4
0.5
0.6
0.7
0.8
0.9
0.0
0.5
1.0
1.5
Bilophila (%)

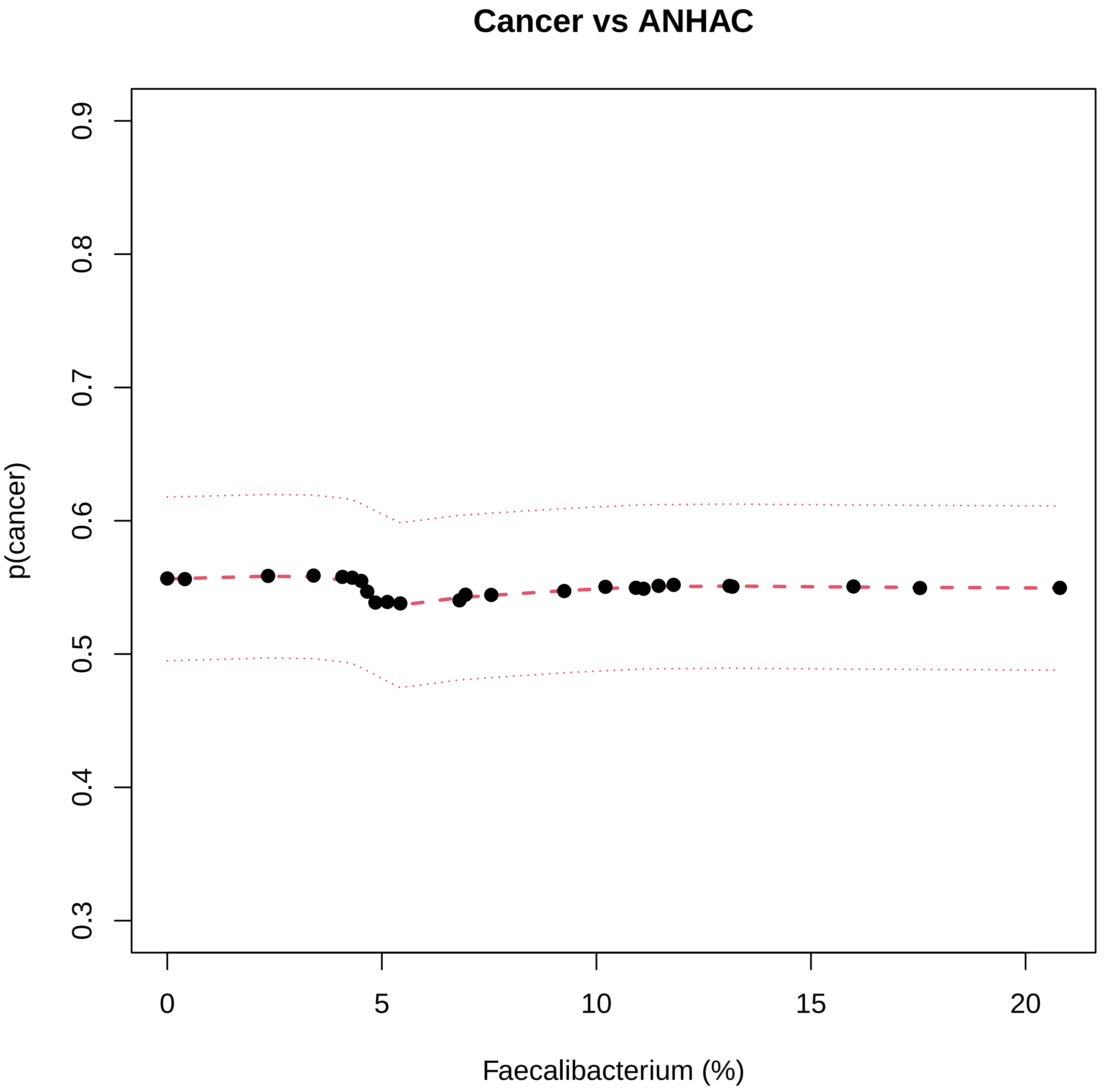
Cancer vs ANHAC
p(cancer)
0.3
0.4
0.5
0.6
0.7
0.8
0.9
0
5
10
15
20
Faecalibacterium (%)

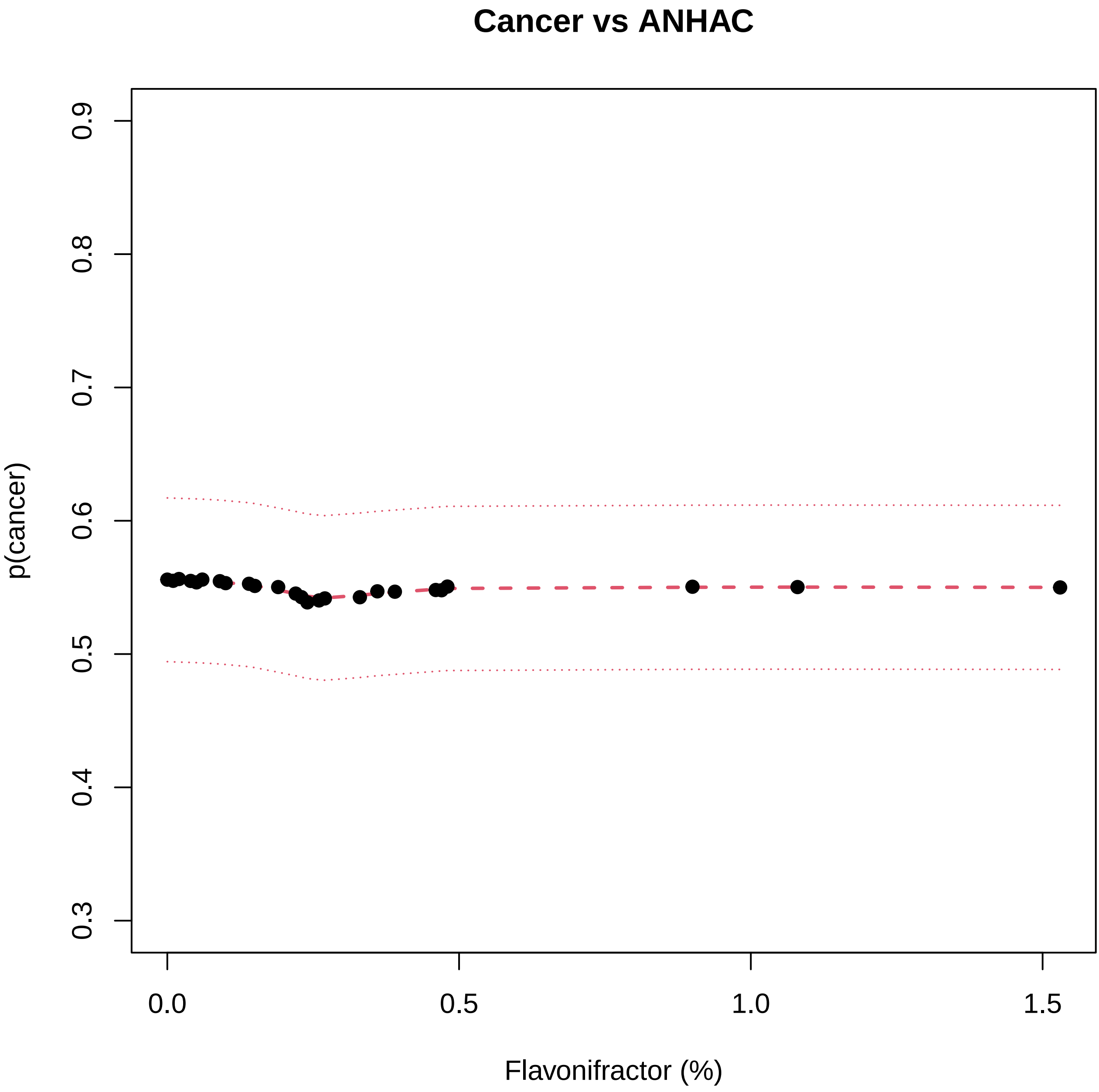

Cancer vs ANHAC
p(cancer)
0.3
0.4
0.5
0.6
0.7
0.8
0.9
0.0
0.5
1.0
1.5
Flavonifractor (%)

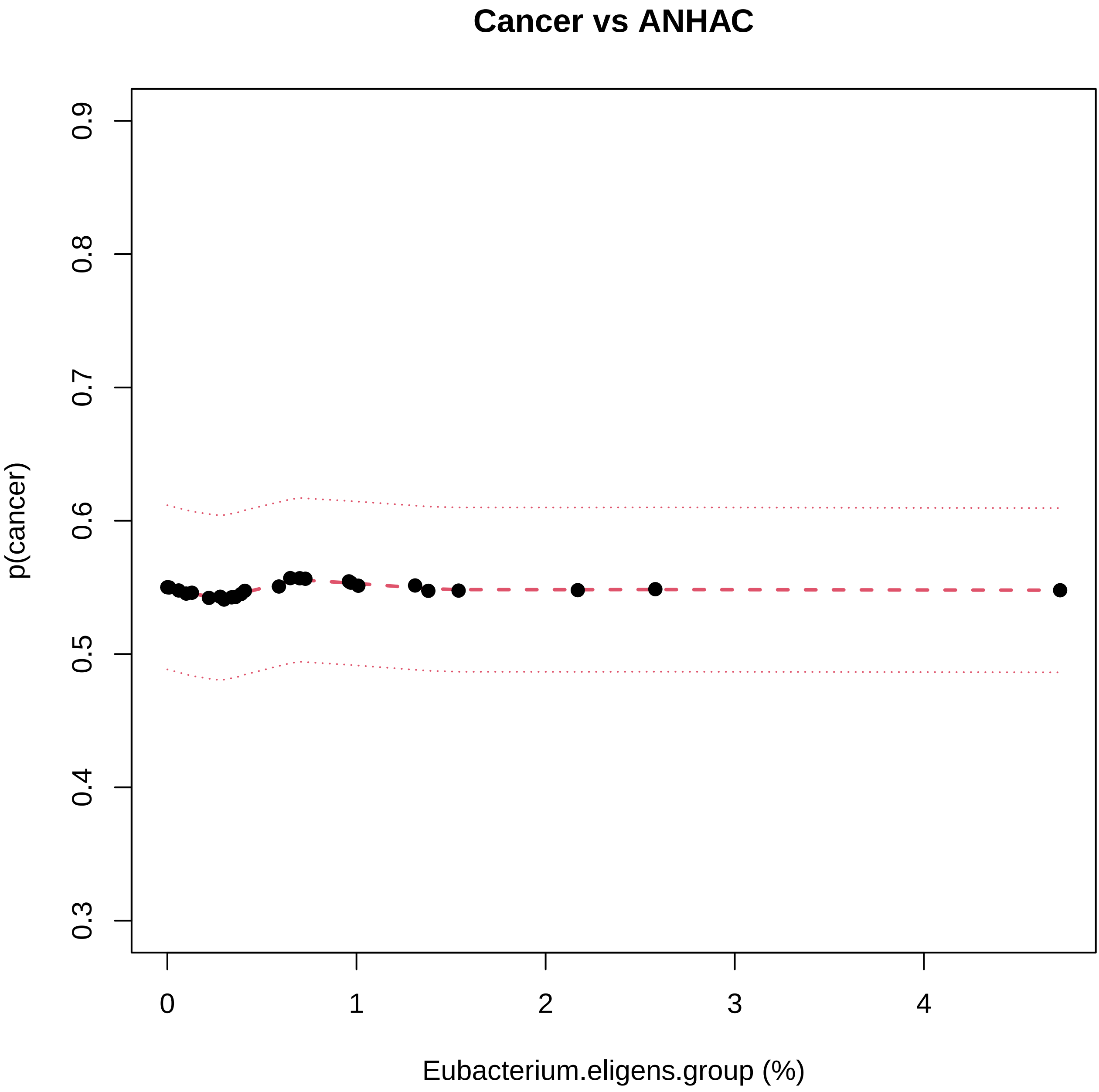

Cancer vs ANHAC
p(cancer)
0.3
0.4
0.5
0.6
0.7
0.8
0.9
0
1
2
3
4
Eubacterium.eligens.group (%)

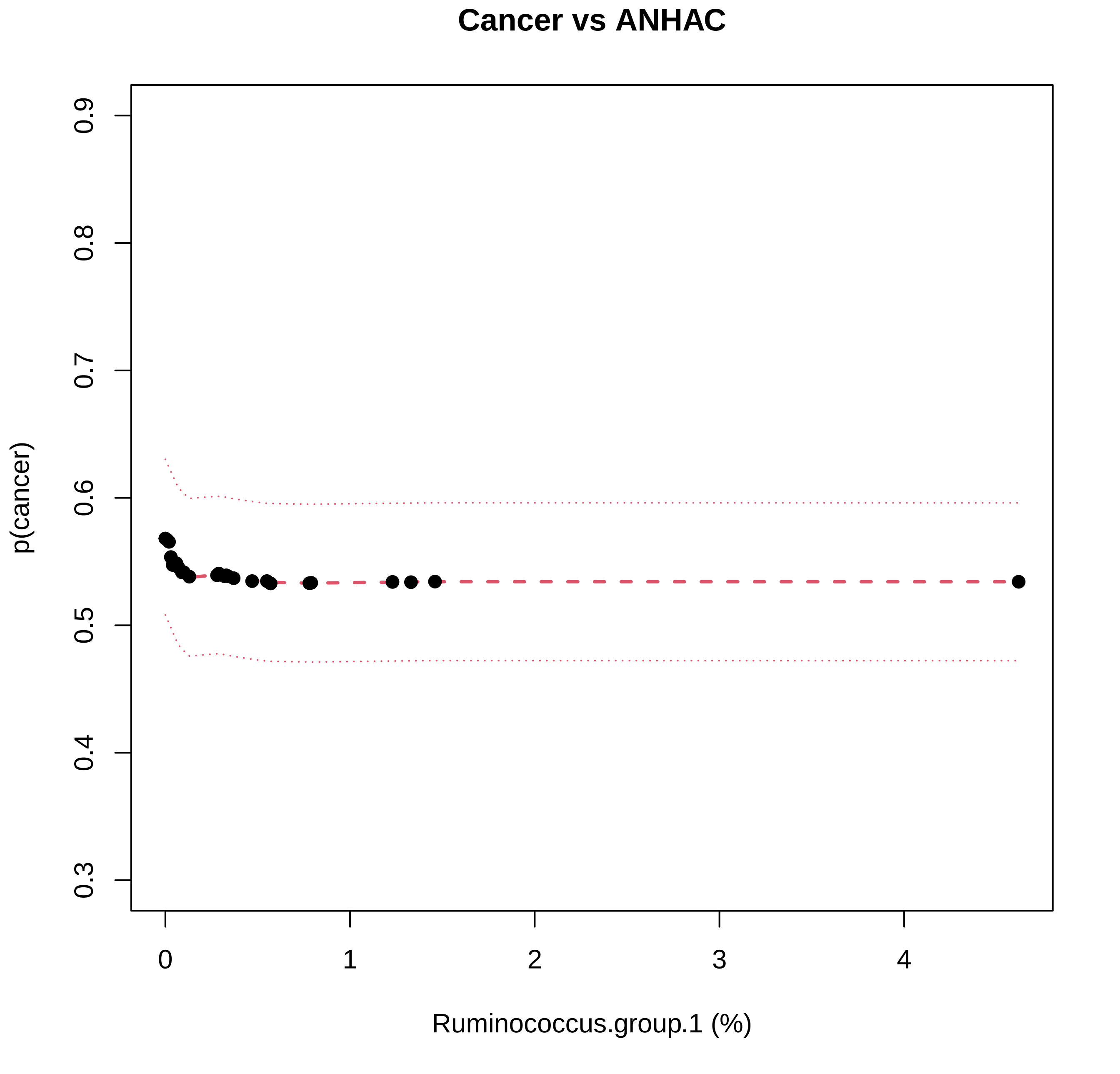
Cancer vs ANHAC
p(cancer)
0.3
0.4
0.5
0.6
0.7
0.8
0.9
0
1
2
3
4
Ruminococcus.group.1 (%)

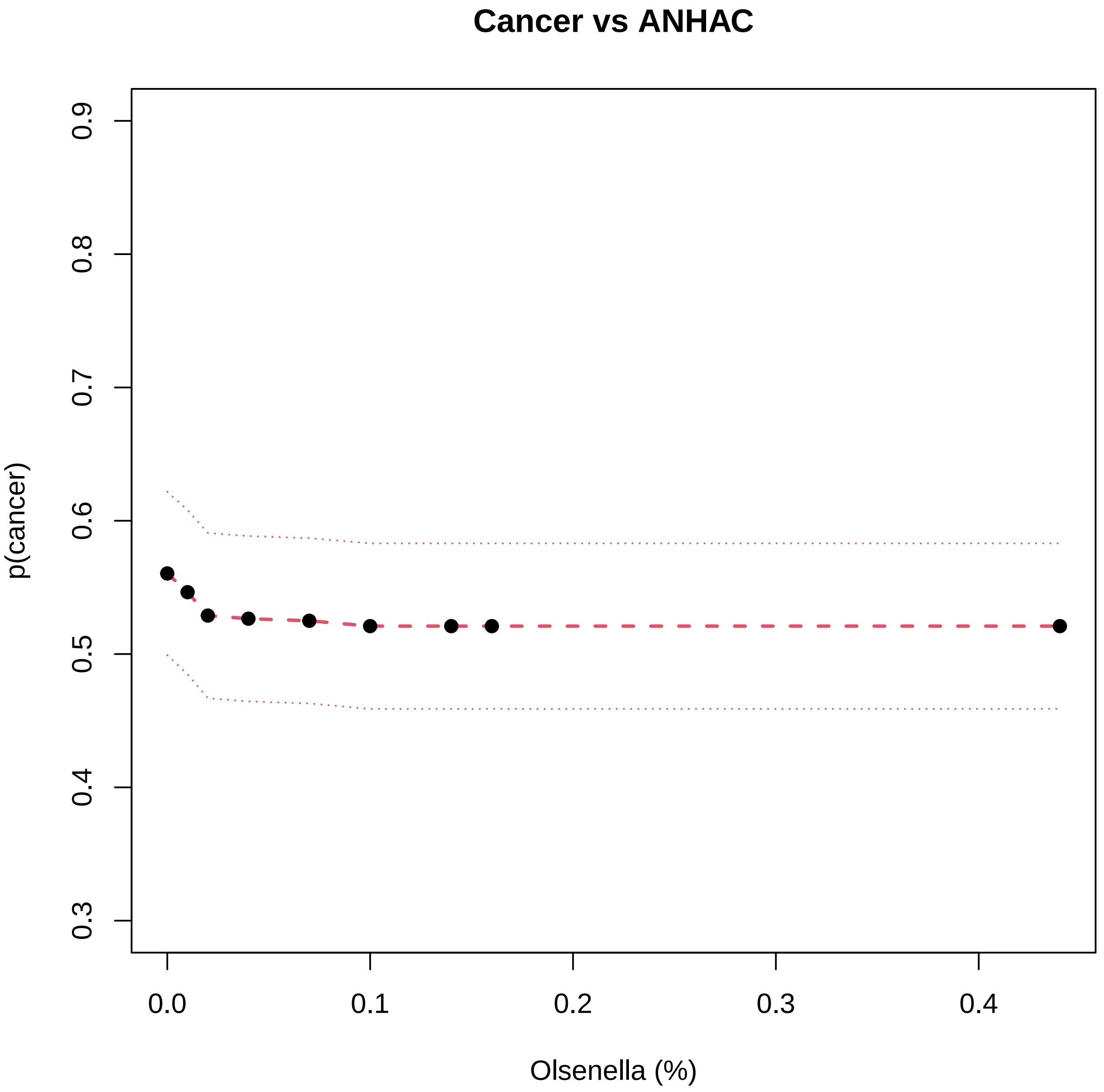

Cancer vs ANHAC
p(cancer)
0.3
0.4
0.5
0.6
0.7
0.8
0.9
0.0
0.1
0.2
0.3
0.4
Olsenella (%)

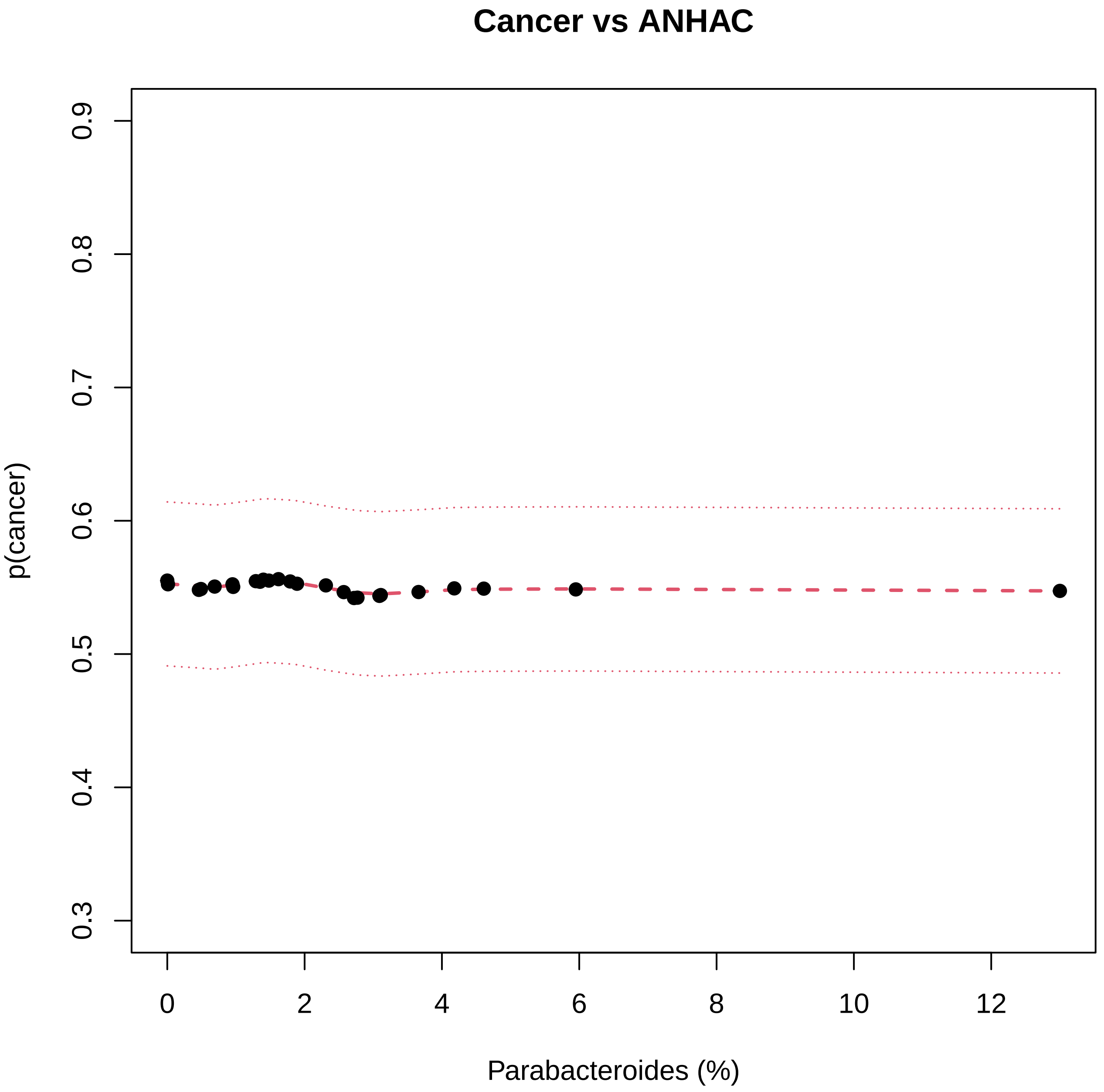

Cancer vs ANHAC
p(cancer)
0.3
0.4
0.5
0.6
0.7
0.8
0.9
0
2
4
6
8
10
12
Parabacteroides (%)

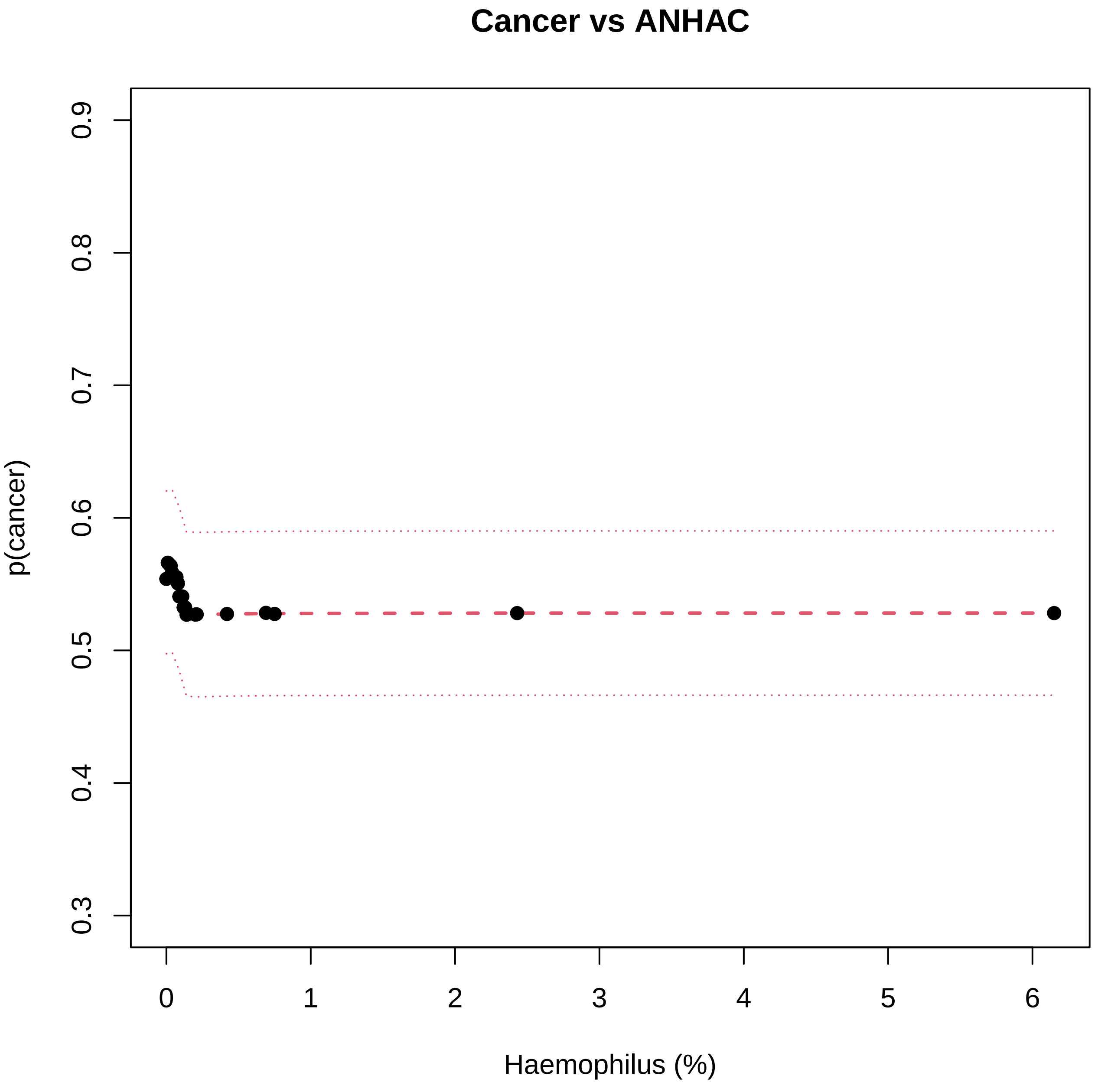
Cancer vs ANHAC
p(cancer)
0.3
0.4
0.5
0.6
0.7
0.8
0.9
0
1
2
3
4
5
6
Haemophilus (%)

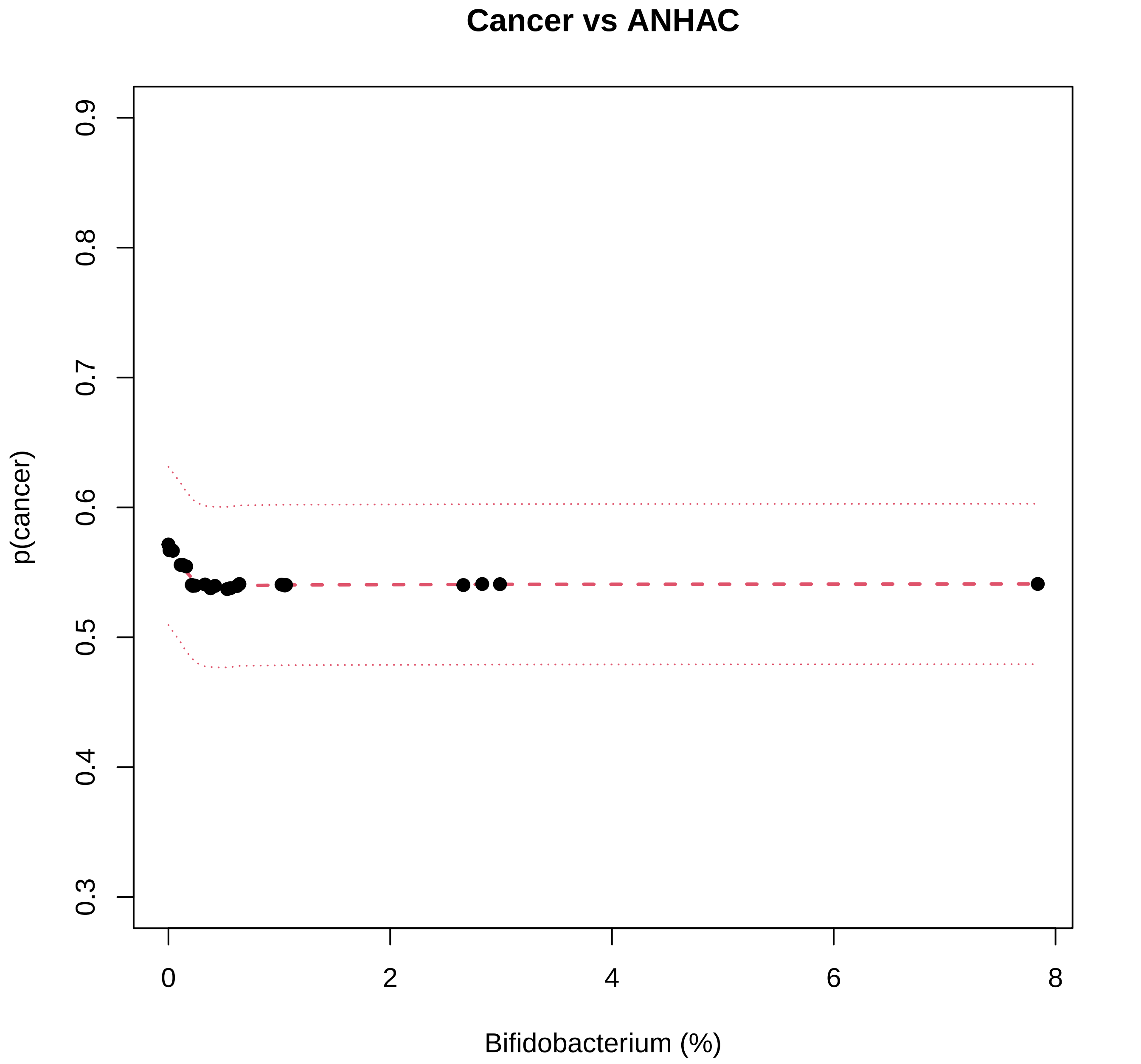

Cancer vs ANHAC
p(cancer)
0.3
0.4
0.5
0.6
0.7
0.8
0.9
0
2
4
6
8
Bifidobacterium (%)

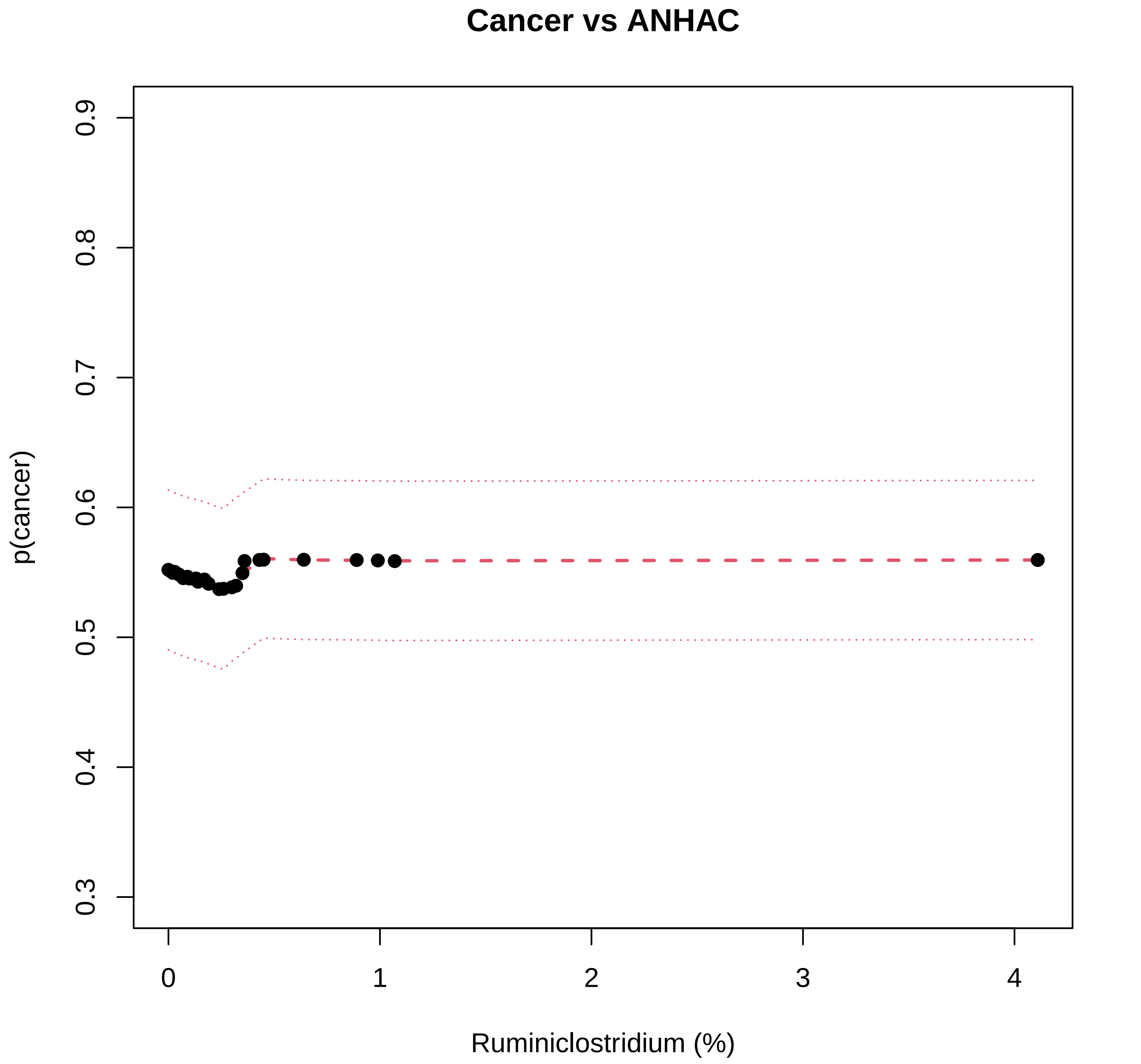
Cancer vs ANHAC
p(cancer)
0.3
0.4
0.5
0.6
0.7
0.8
0.9
0
1
2
3
4
Ruminiclostridium (%)

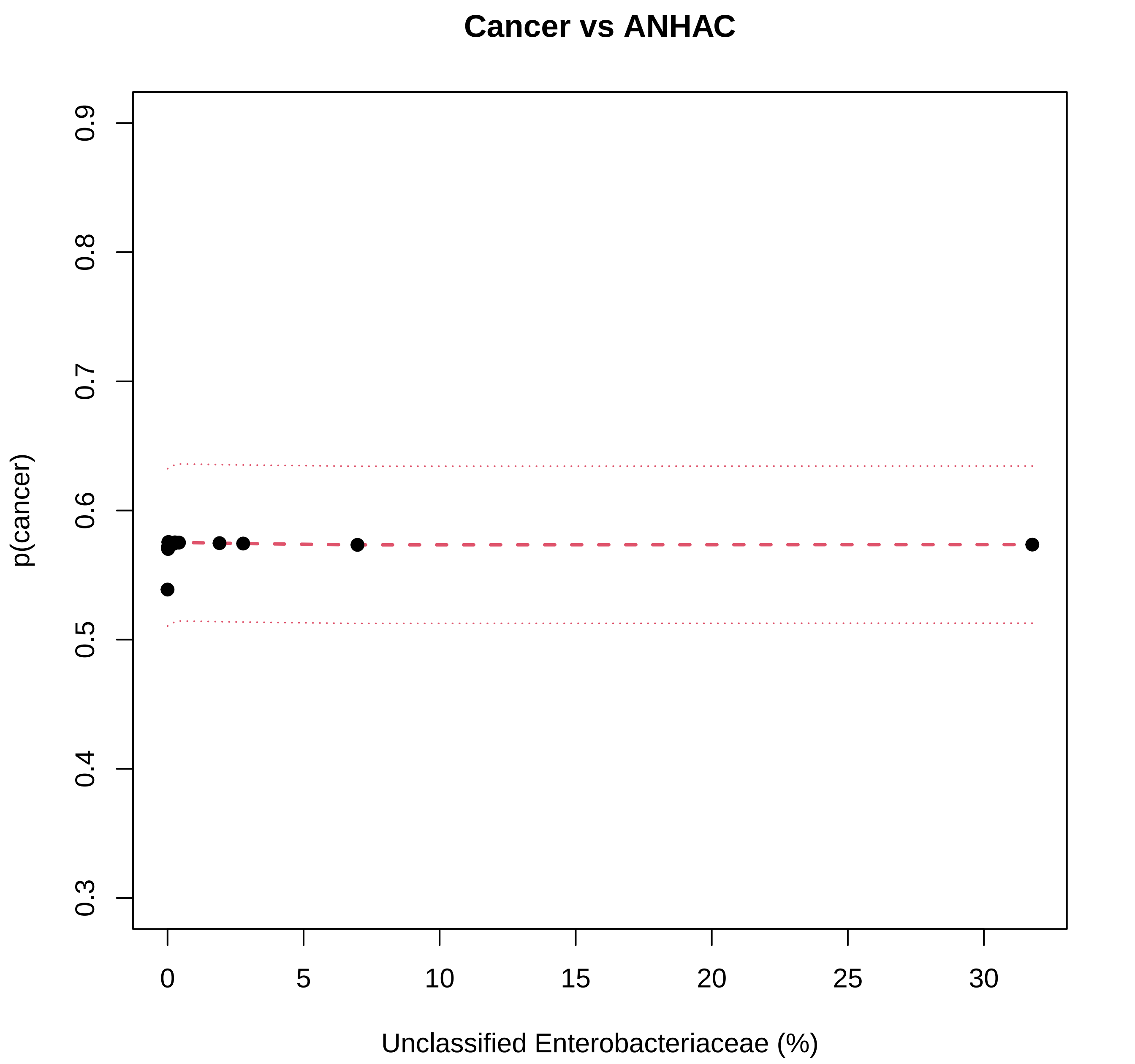
Cancer vs ANHAC
p(cancer)
0.3
0.4
0.5
0.6
0.7
0.8
0.9
0
5
10
15
20
25
30
Unclassified Enterobacteriaceae (%)

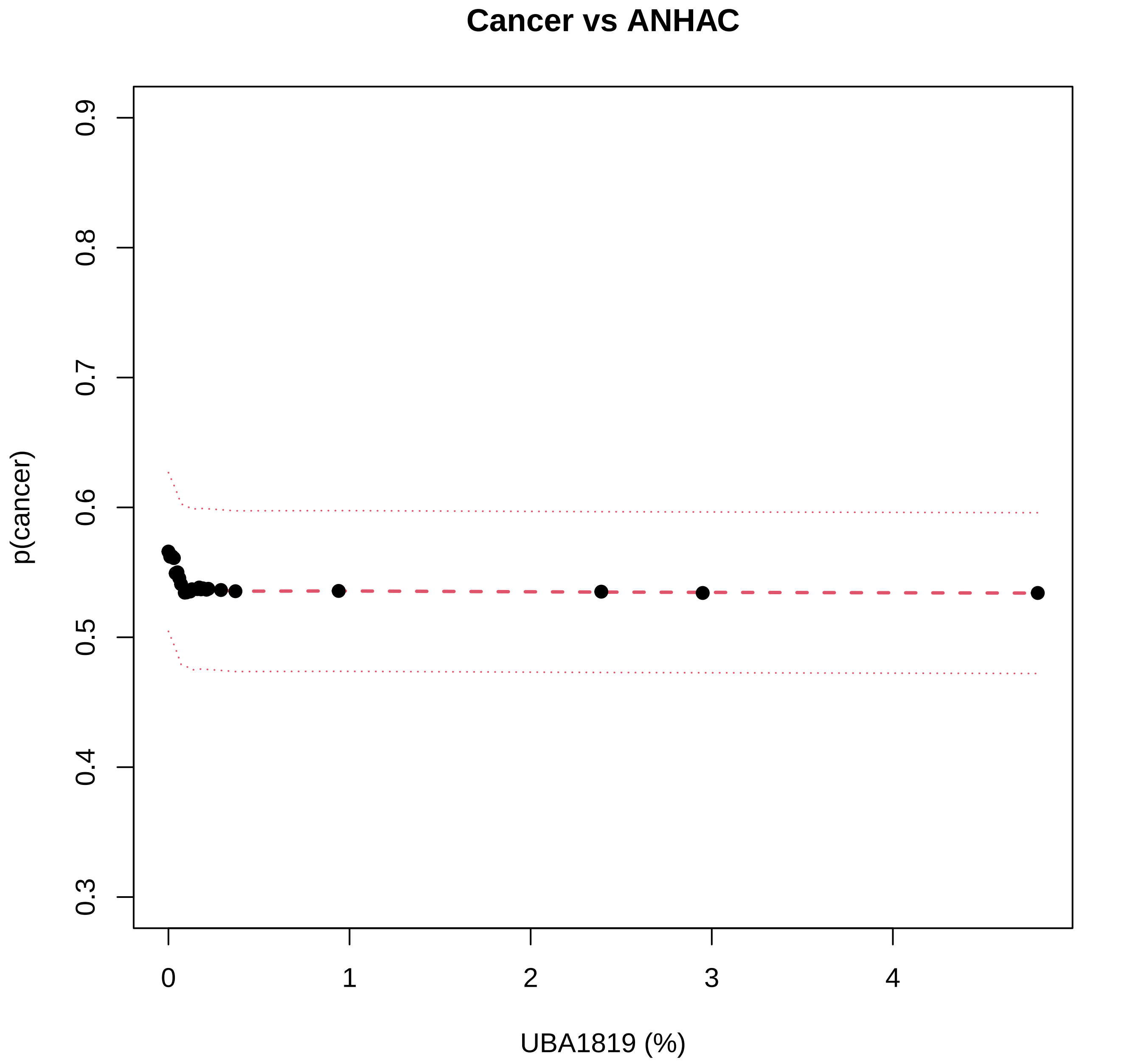

Cancer vs ANHAC
p(cancer)
0.3
0.4
0.5
0.6
0.7
0.8
0.9
0
1
2
3
4
UBA1819 (%)

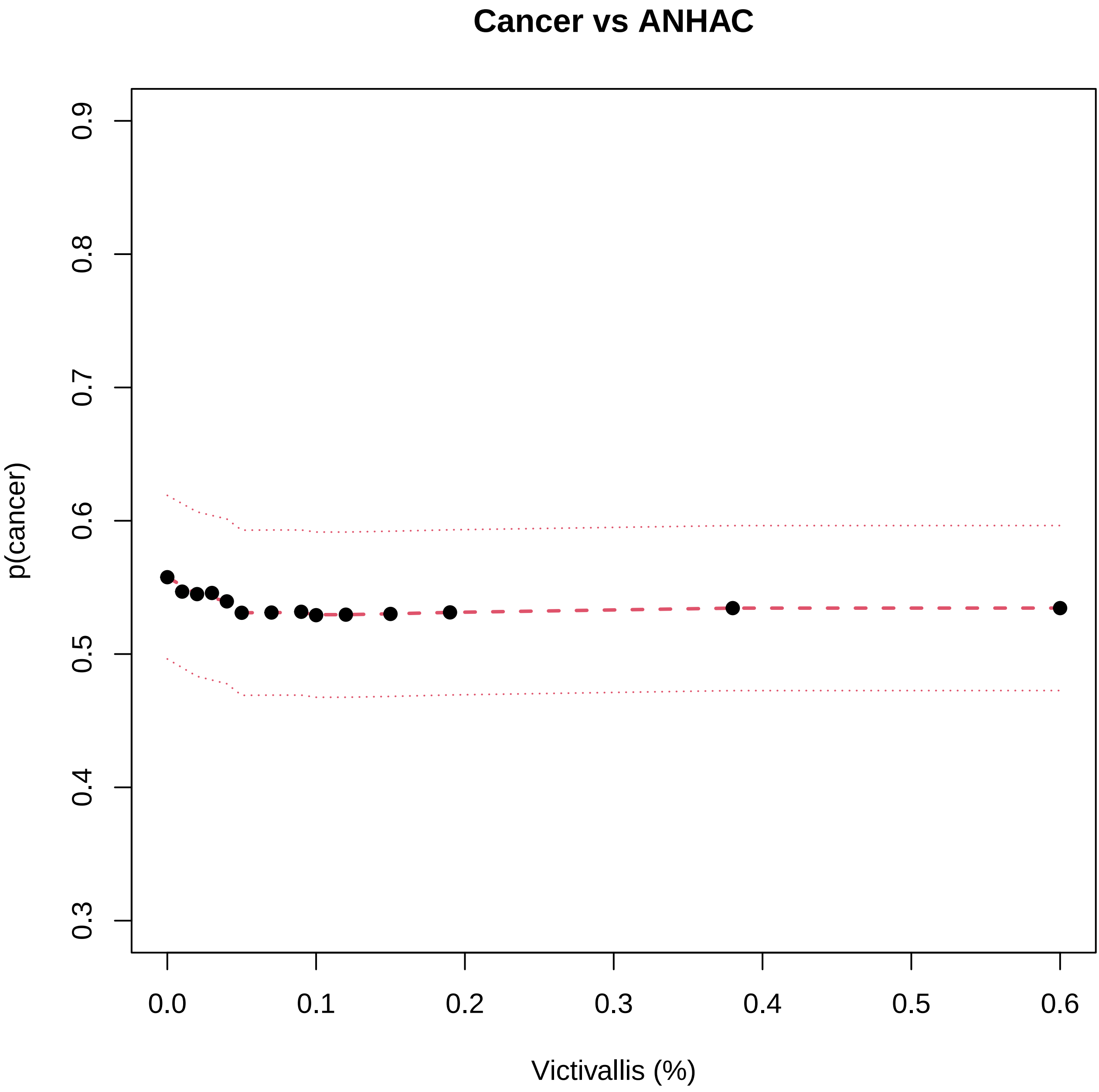
Cancer vs ANHAC
p(cancer)
0.3
0.4
0.5
0.6
0.7
0.8
0.9
0.0
0.1
0.2
0.3
0.4
0.5
0.6
Victivallis (%)

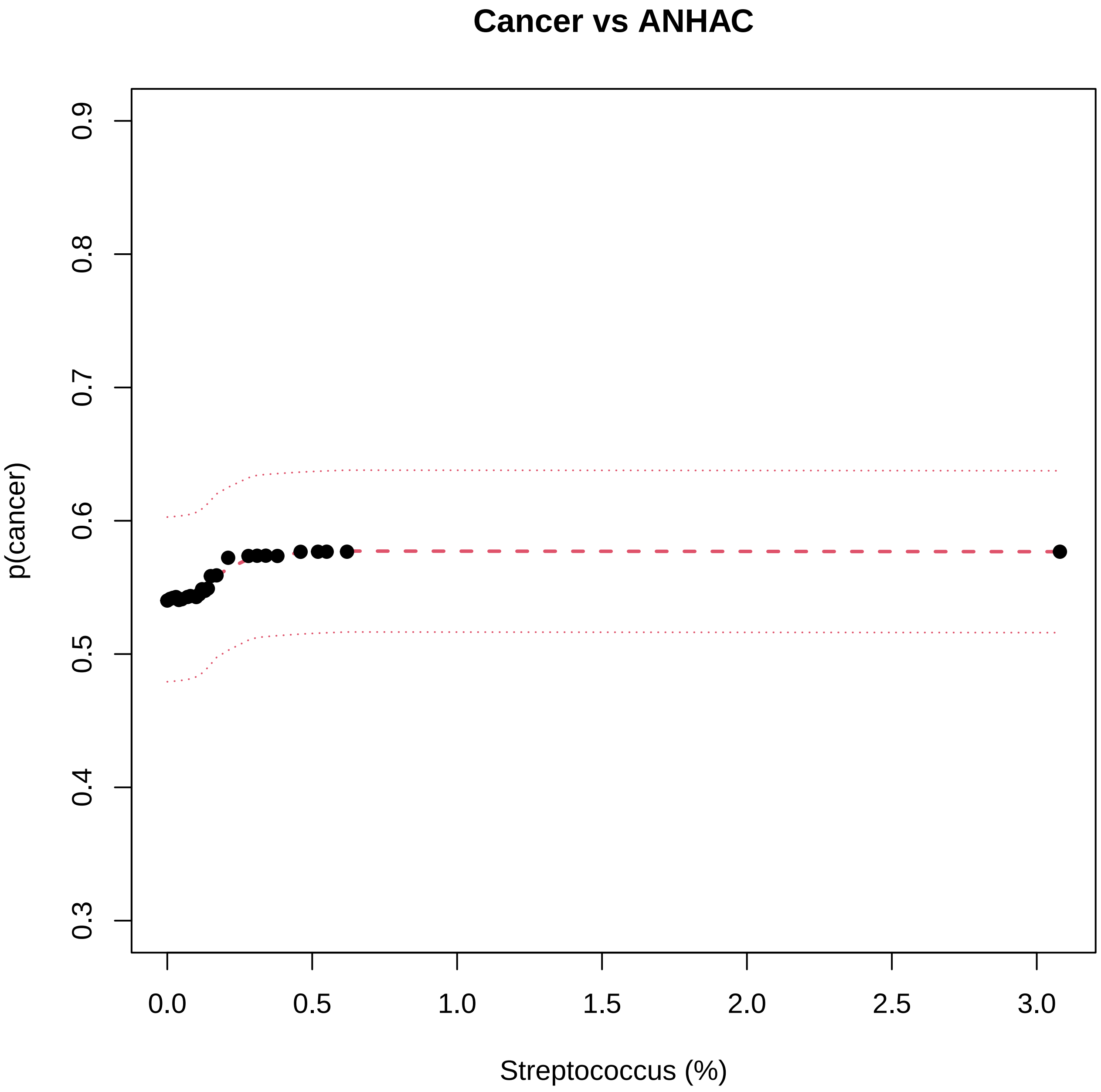
Cancer vs ANHAC
p(cancer)
0.3
0.4
0.5
0.6
0.7
0.8
0.9
0.0
0.5
1.0
1.5
2.0
2.5
3.0
Streptococcus (%)

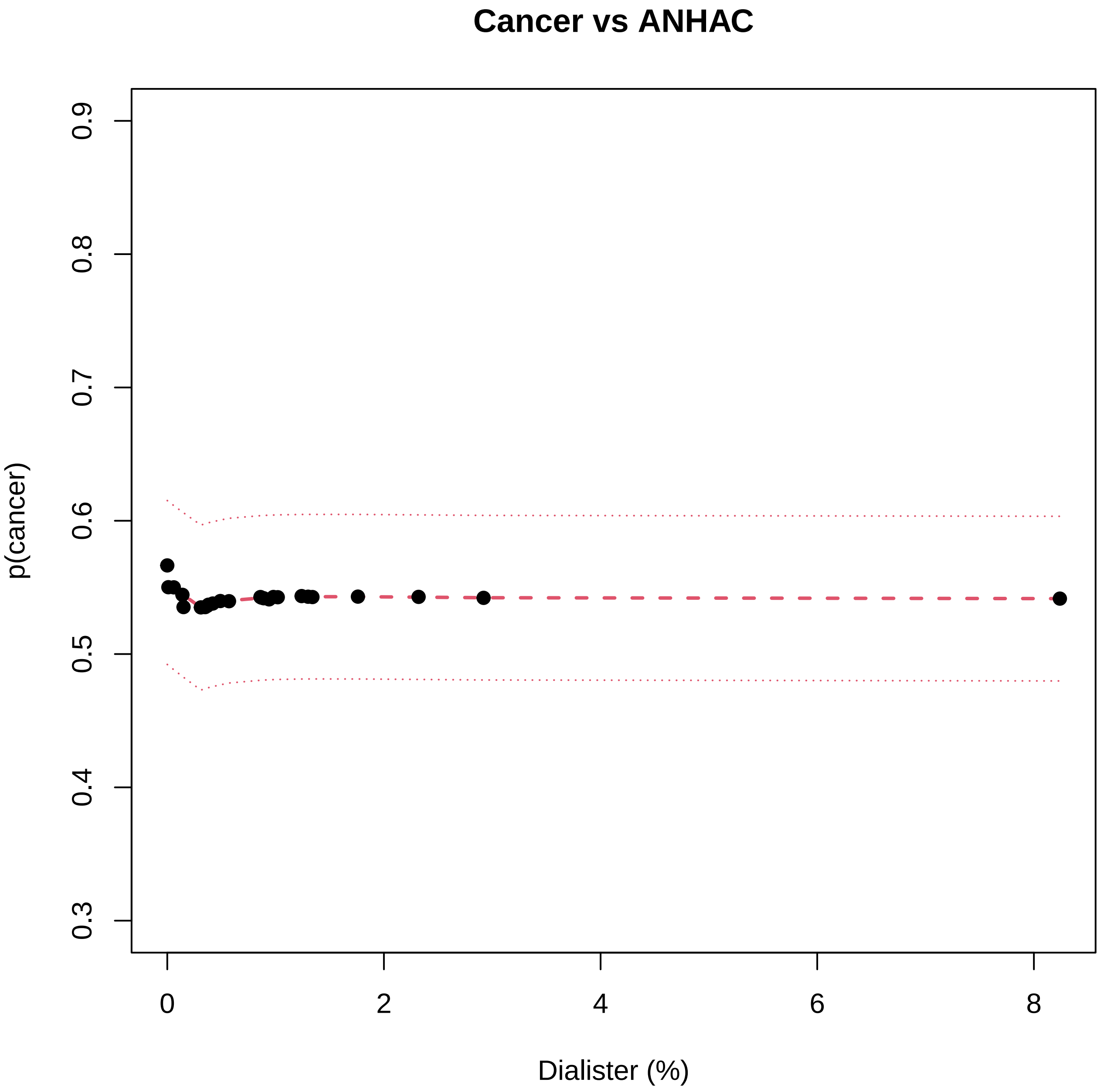

Cancer vs ANHAC
p(cancer)
0.3
0.4
0.5
0.6
0.7
0.8
0.9
0
2
4
6
8
Dialister (%)

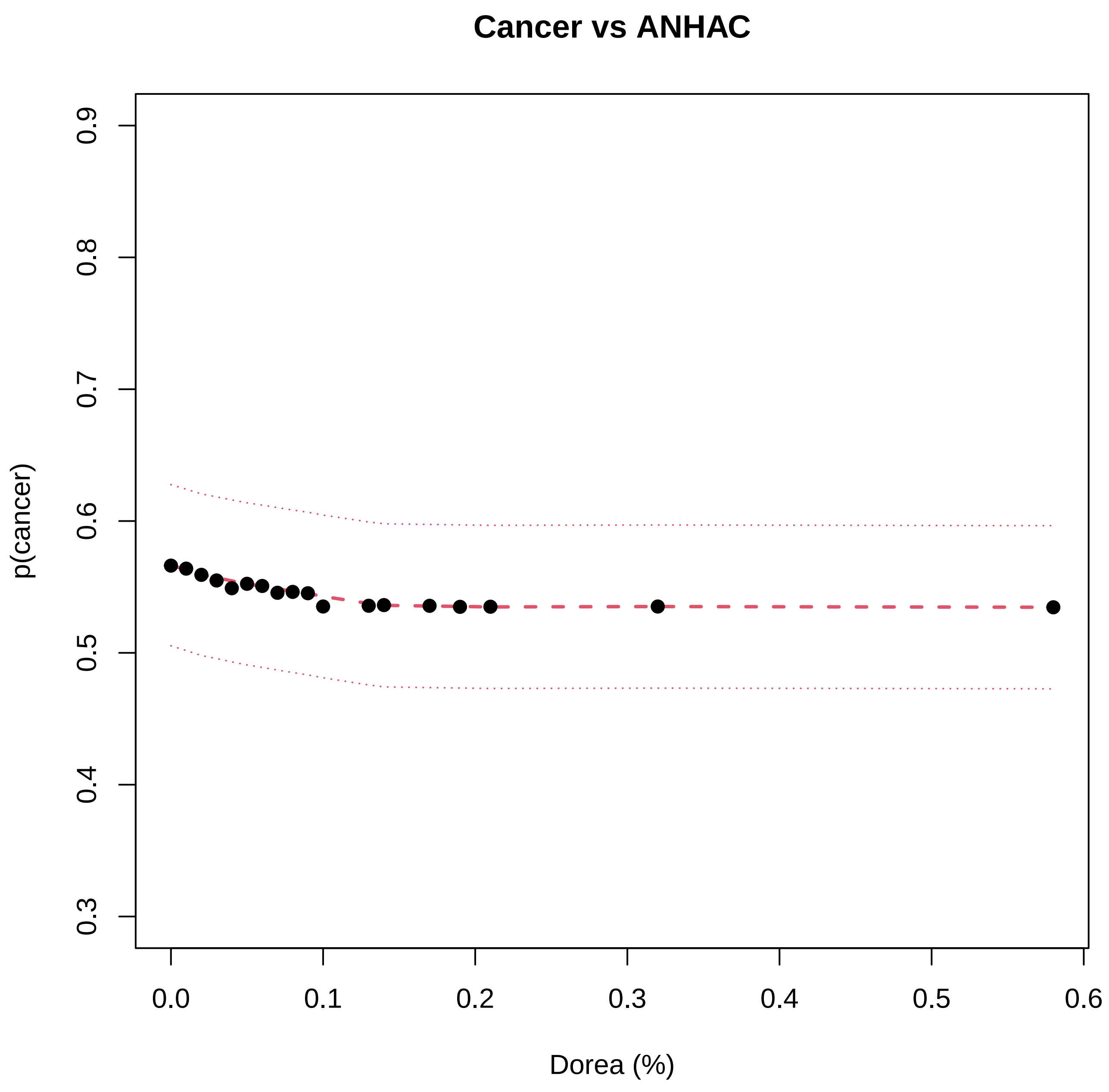

Cancer vs ANHAC
p(cancer)
0.3
0.4
0.5
0.6
0.7
0.8
0.9
0.0
0.1
0.2
0.3
0.4
0.5
0.6
Dorea (%)

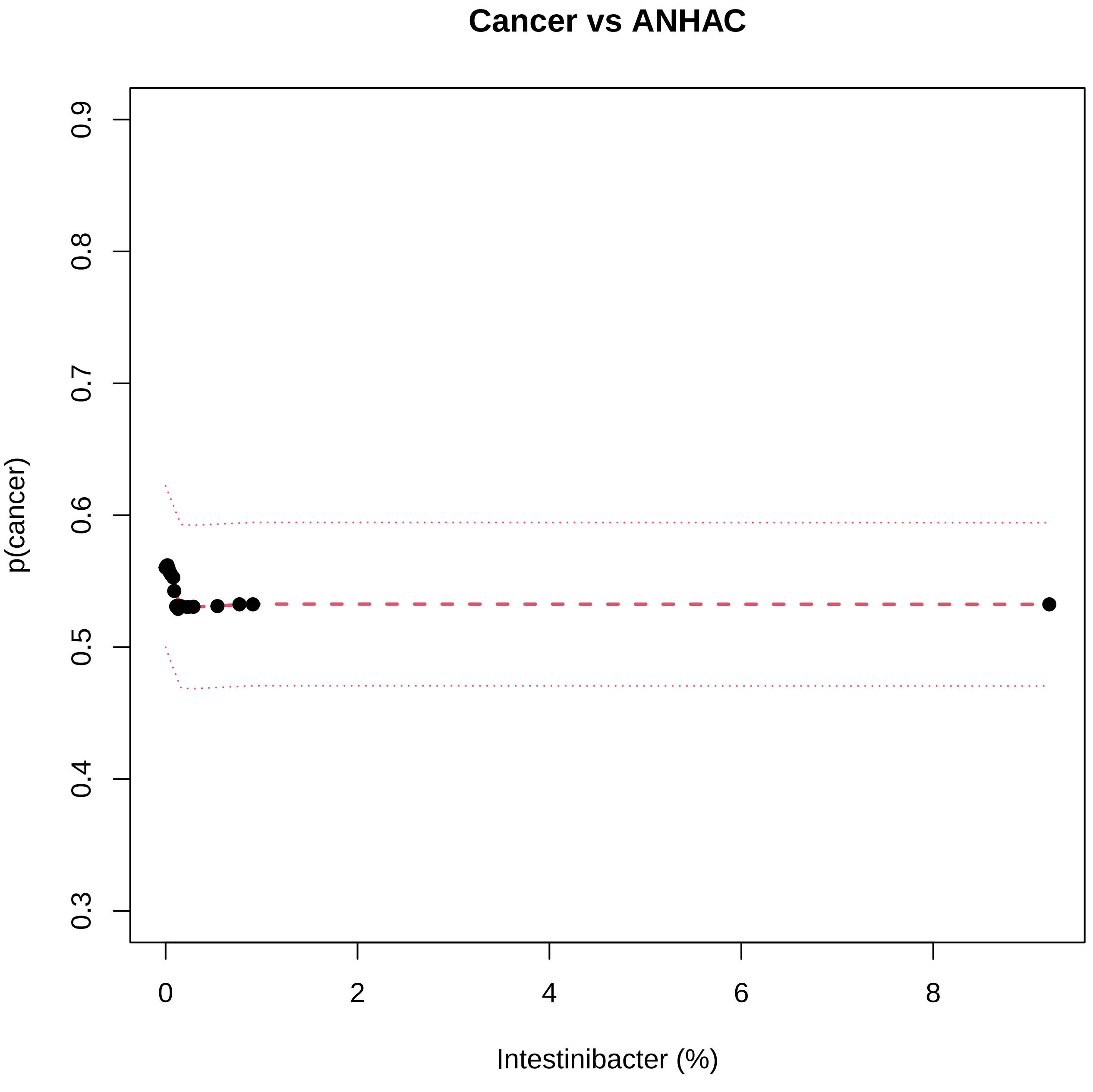
Cancer vs ANHAC
p(cancer)
0.3
0.4
0.5
0.6
0.7
0.8
0.9
0
2
4
6
8
Intestinibacter (%)

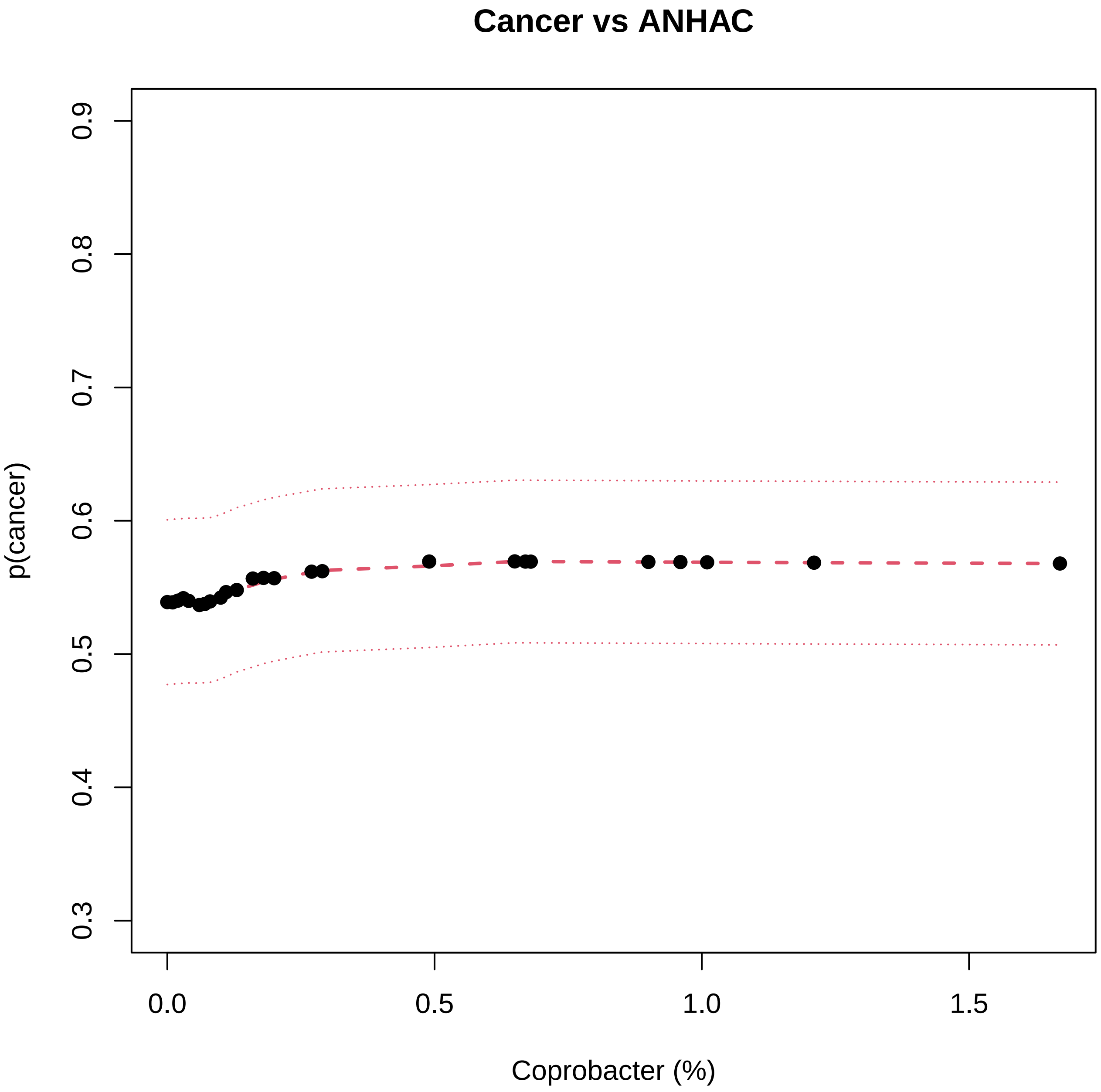
Cancer vs ANHAC
p(cancer)
0.3
0.4
0.5
0.6
0.7
0.8
0.9
0.0
0.5
1.0
1.5
Coprobacter (%)

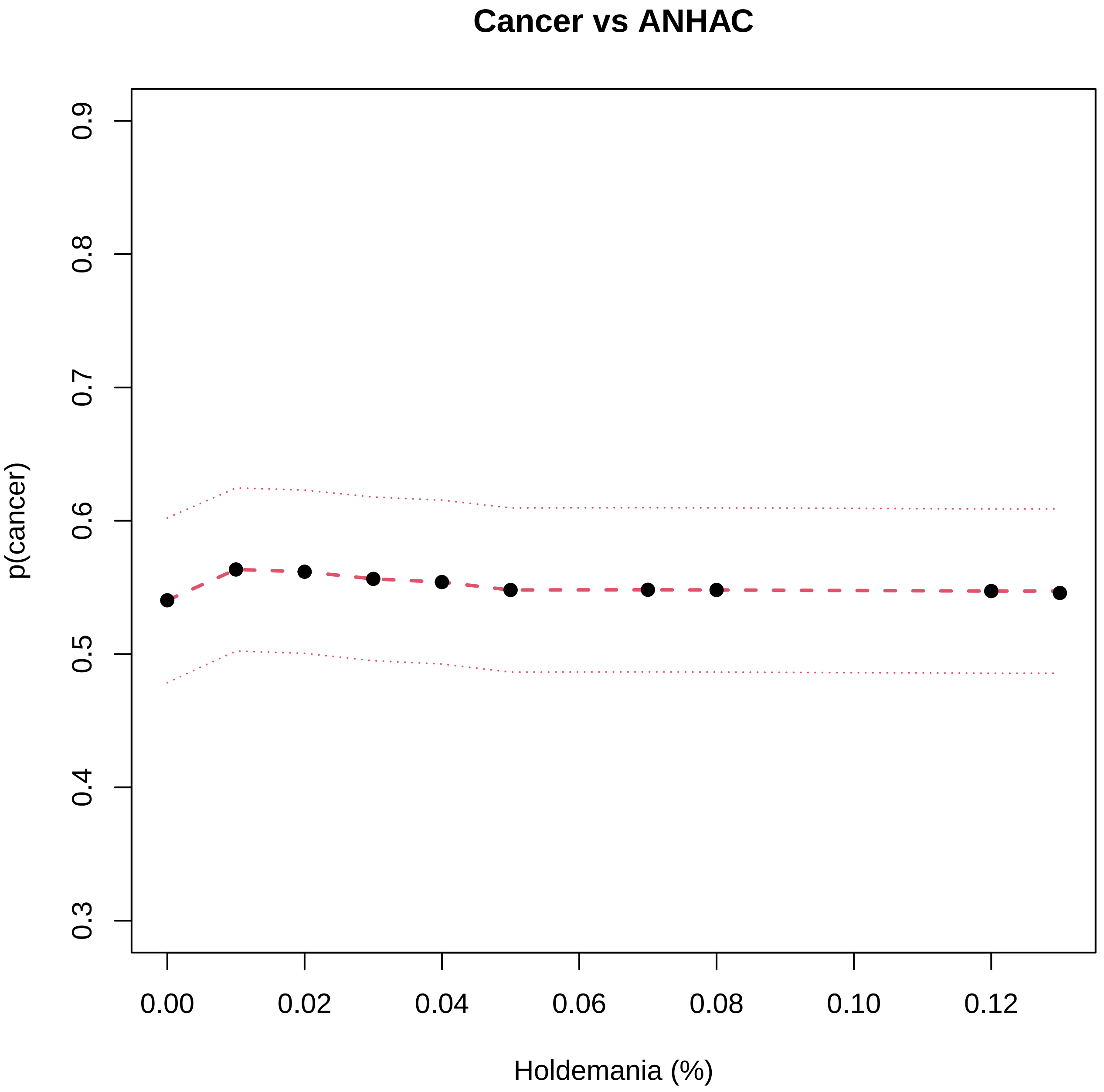

Cancer vs ANHAC
p(cancer)
0.3
0.4
0.5
0.6
0.7
0.8
0.9
0.00
0.02
0.04
0.06
0.08
0.10
0.12
Holdemania (%)

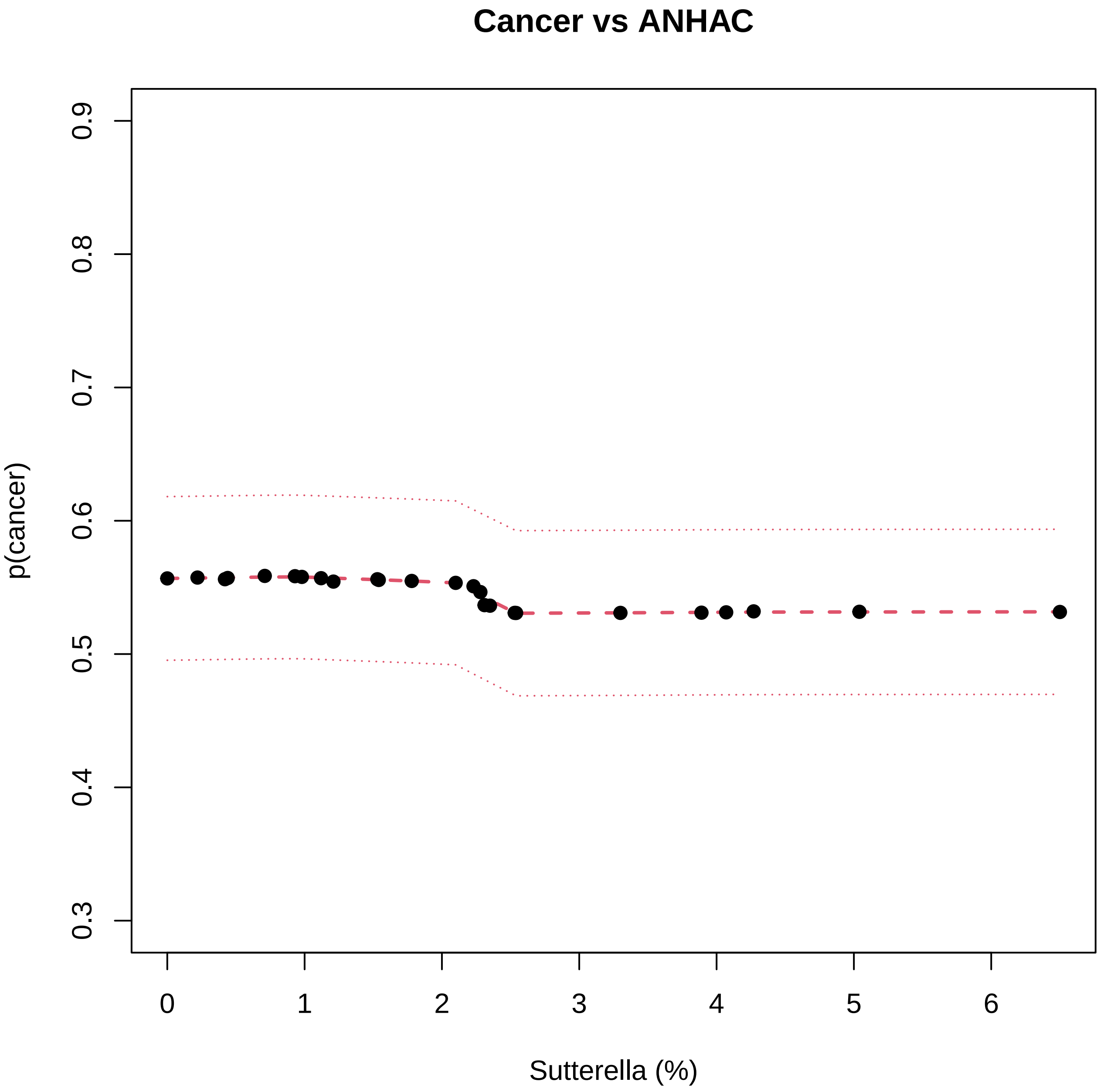

Cancer vs ANHAC
p(cancer)
0.3
0.4
0.5
0.6
0.7
0.8
0.9
0
1
2
3
4
5
6
Sutterella (%)

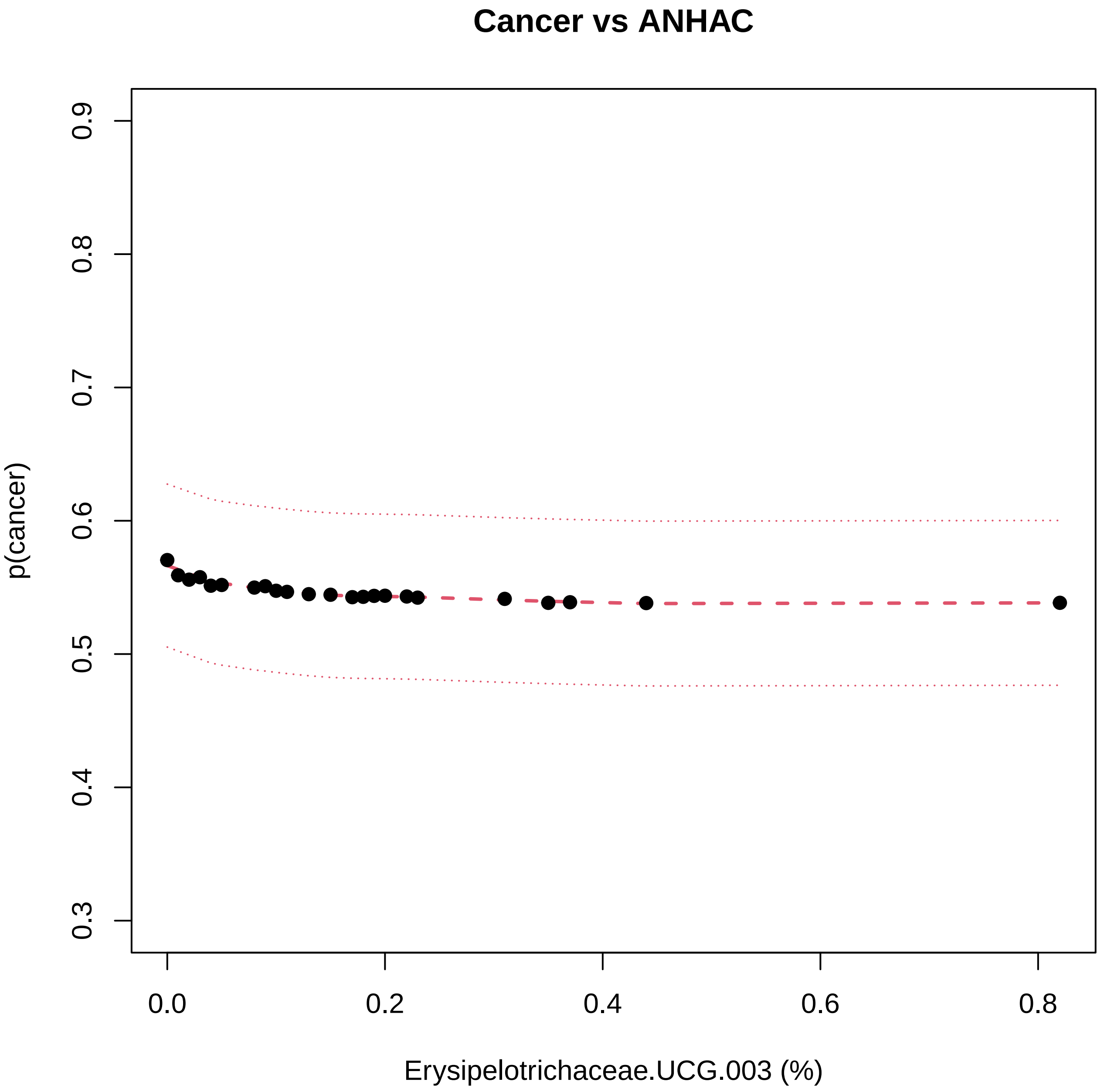

Cancer vs ANHAC
p(cancer)
0.3
0.4
0.5
0.6
0.7
0.8
0.9
0.0
0.2
0.4
0.6
0.8
Erysipelotrichaceae.UCG.003 (%)

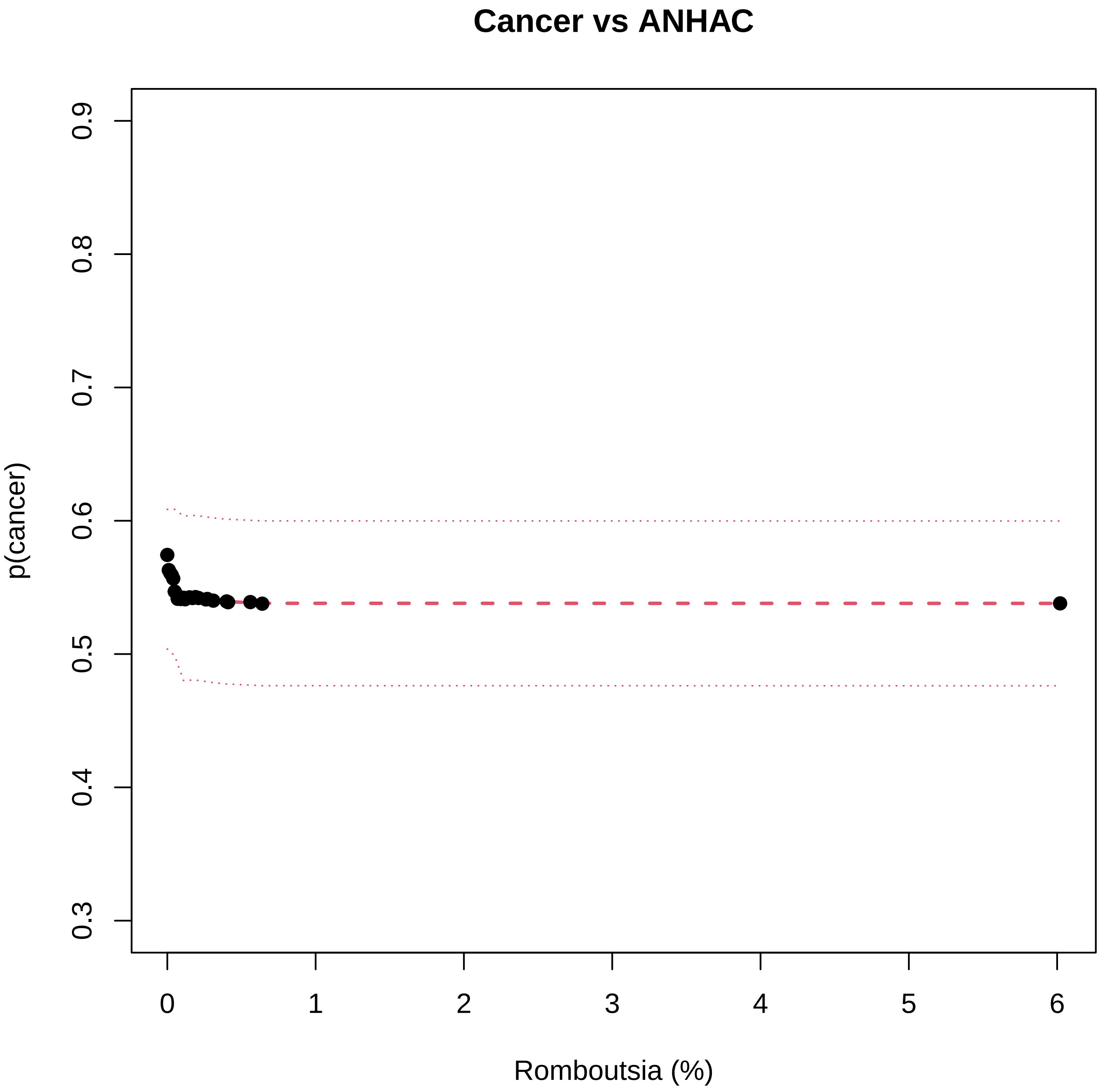

Cancer vs ANHAC
p(cancer)
0.3
0.4
0.5
0.6
0.7
0.8
0.9
0
1
2
3
4
5
6
Romboutsia (%)

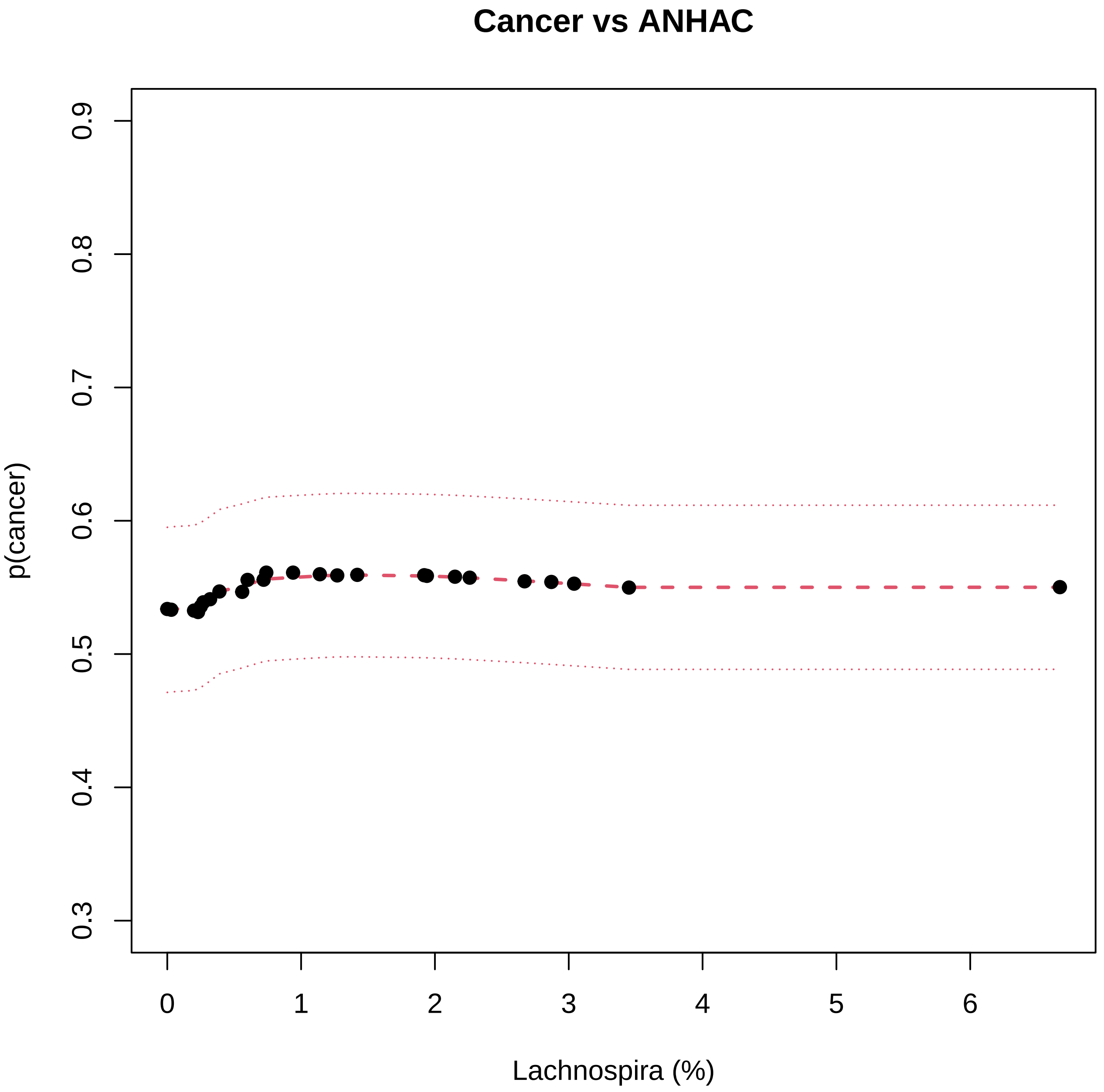
Cancer vs ANHAC
p(cancer)
0.3
0.4
0.5
0.6
0.7
0.8
0.9
0
1
2
3
4
5
6
Lachnospira (%)

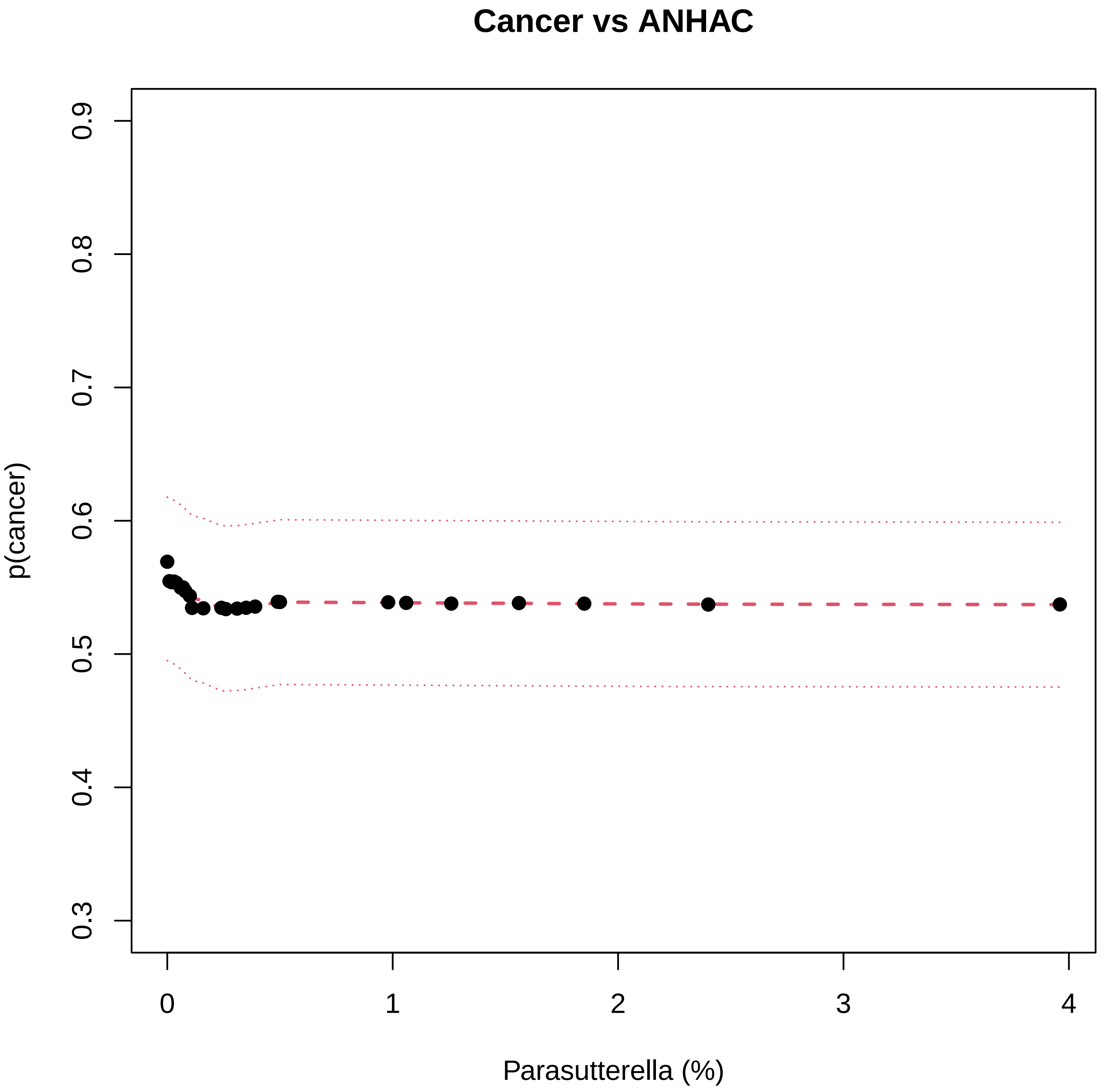

Cancer vs ANHAC
p(cancer)
0.3
0.4
0.5
0.6
0.7
0.8
0.9
0
1
2
3
4
Parasutterella (%)

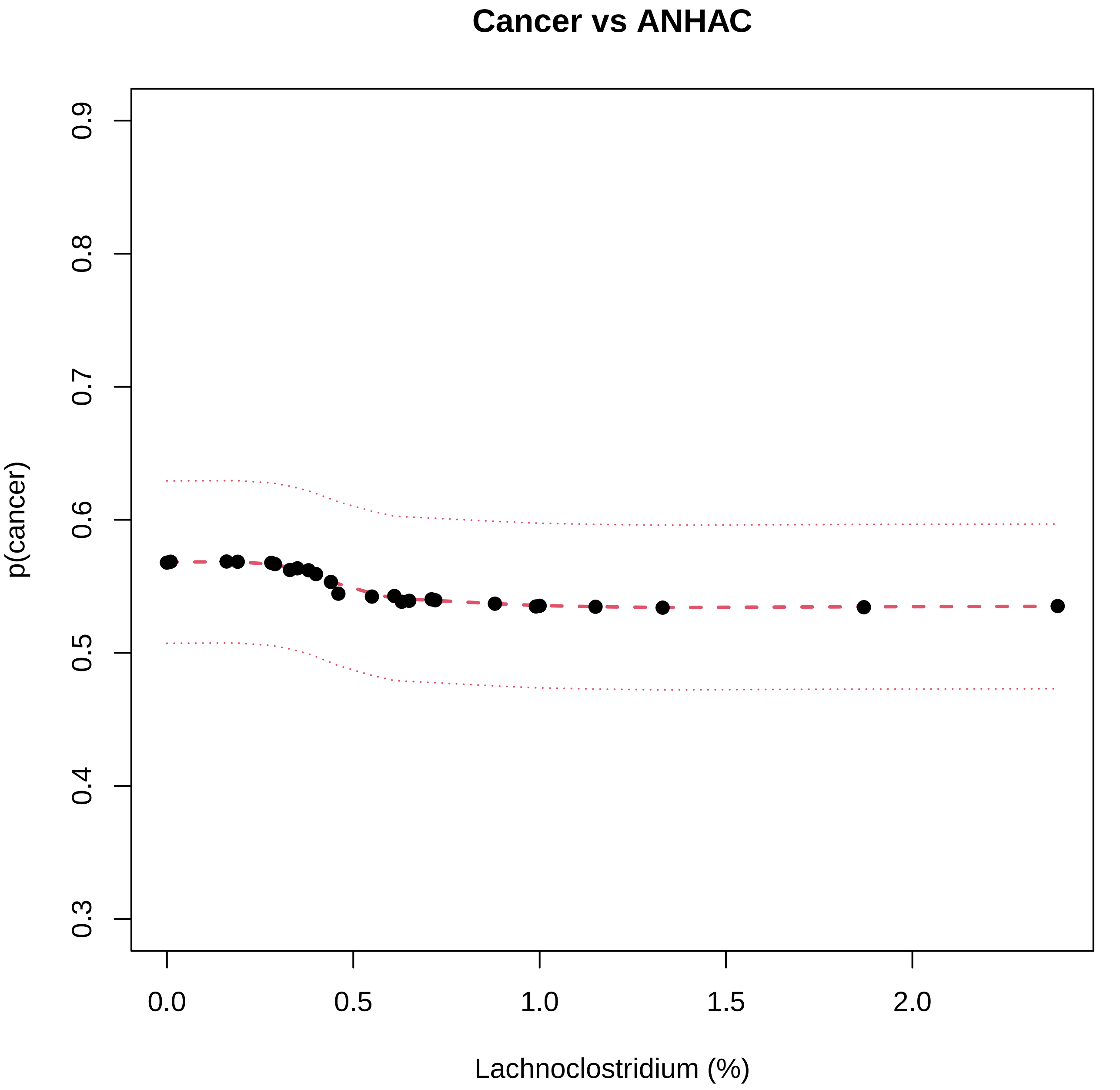

Cancer vs ANHAC
p(cancer)
0.3
0.4
0.5
0.6
0.7
0.8
0.9
0.0
0.5
1.0
1.5
2.0
Lachnoclostridium (%)

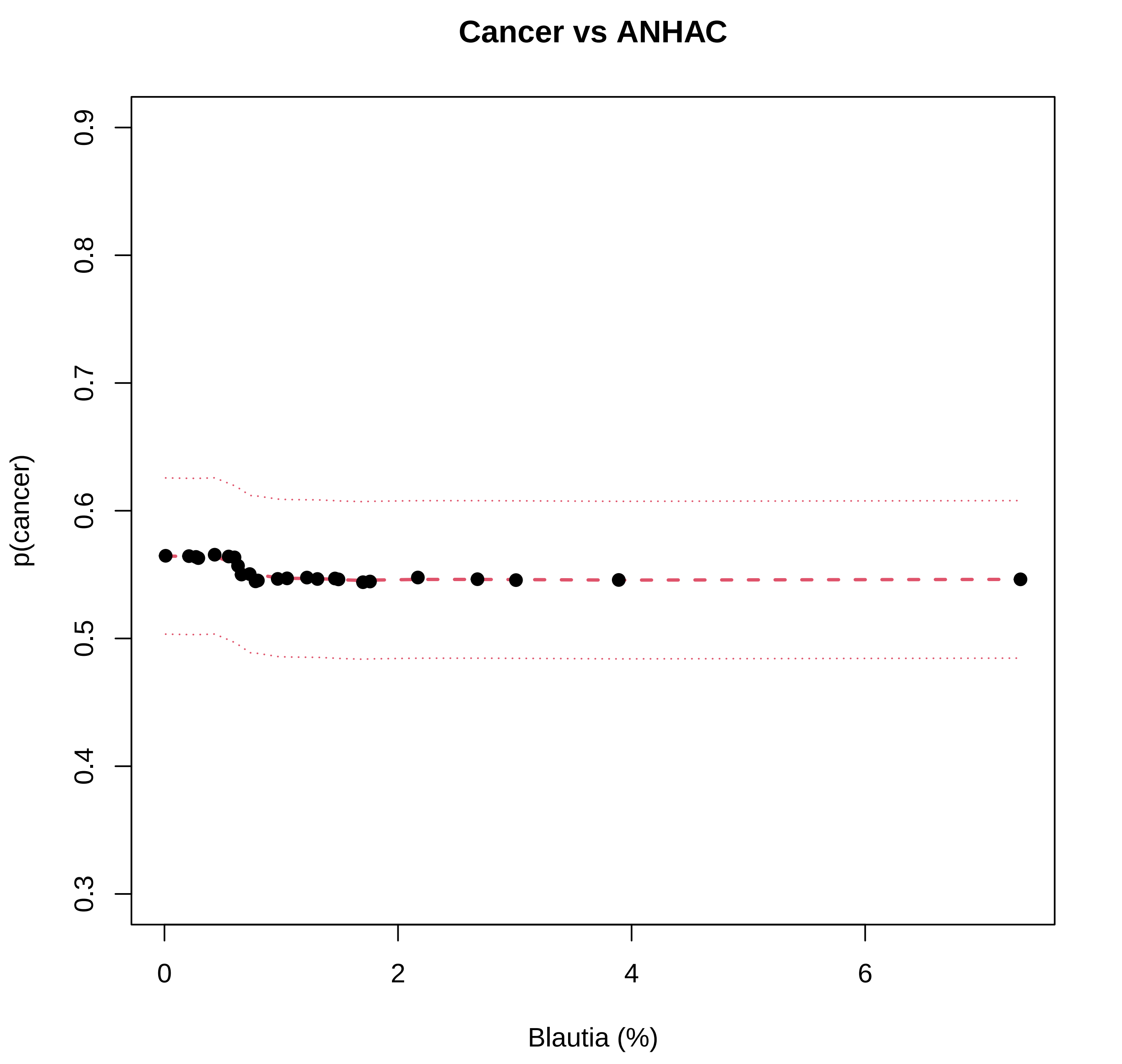
Cancer vs ANHAC
p(cancer)
0.9
0.8
0.7
0.6
0.5
0.4
0.3
0
2
4
6
Blautia (%)

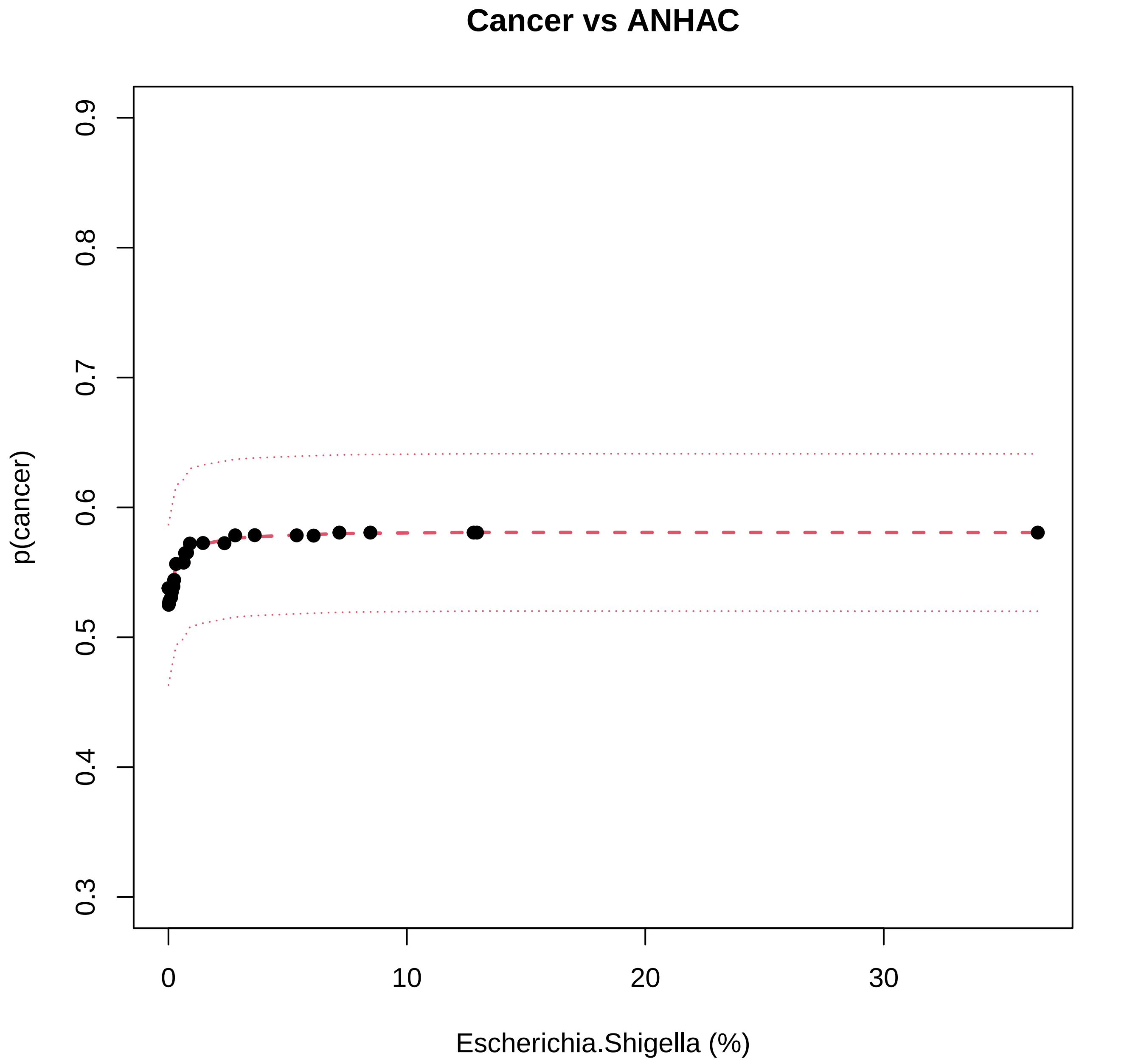
Cancer vs ANHAC
p(cancer)
0.3
0.4
0.5
0.6
0.7
0.8
0.9
0
10
20
30
Escherichia.Shigella (%)
